# Associations of Baseline Clinical Phenotypes with White Matter Hyperintensity Volume Change – A Study of 4329 UK Biobank Participants

**DOI:** 10.1101/2025.11.18.25340385

**Authors:** Angelina Kancheva, Louise Millard, Donald Lyall, Joanna Wardlaw, Terence Quinn

## Abstract

**Background and Objectives:** White matter hyperintensities (WMHs) relate to cognitive and physical impairment. Although WMHs typically increase/progress with time, regression/decreasing has also been observed. We tested which baseline clinical phenotypes predict subsequent WMH volume change in UK Biobank (UKB).

**Methods:** We included participants with total volume of WMHs at first brain magnetic resonance imaging (MRI) and follow-up brain MRI. We assessed whether 107 pre-selected clinical phenotypes at baseline related to follow-up WMH volume change. We derived a measure of WMH change as the residual from a linear regression of follow-up WMH volume on baseline WMH volume. We pre-processed clinical phenotypes using PHESANT. We ran (i) linear regressions for continuous WMH change as the outcome and (ii) multinomial logistic regressions comparing progression, regression, and stable groups after categorising WMH volume change using a quintile-based approach. Models were unadjusted, partially adjusted (age, sex, total brain tissue volume, follow-up time), or fully adjusted (further adding blood pressure; BP). We corrected all *P*-values for multiple testing using false discovery rate (FDR) correction.

**Results:** We included 4329 participants (median age=52; IQR=12), 54.6% female. Median follow-up was 2.3 years (range=1-7 years). On average, total brain volume decreased across follow-up (mean decrease=16,058 mm^3^) and WMH volume increased (median increase=293 mm^3^; both *p*<0.001). WMHs progressed in 53.9% of participants, regressed in 26.01%, and remained stable in 20%. Fully-adjusted associations with WMH increase included an increased risk of diabetes-related eye disease and higher diastolic BP (FDR-adjusted *p*<0.05). In group comparisons, progressors were older than regressors and stable individuals, on average. Regressors and progressors had higher BMI than the stable group. Regressors had the highest baseline WMH burden. In unadjusted models, progressors versus stable were more likely to have cataract and higher BP, while regressors had higher weight and systolic BP. These associations did not survive covariate adjustment.

**Discussion:** Well-established vascular risk factors related to subsequent WMH volume change. Distinct clinical and demographic profiles characterized WMH progression, regression, and stable groups. Results suggest that vascular factors relate to WMH change but are sensitive to covariate control. Further studies should establish factors differentially related to WMH progression, regression, and stability.

## Introduction

White matter hyperintensities (WMHs) are the most common neuroimaging feature of cerebral small vessel disease (cSVD)^1^. On T2-weighted and fluid-attenuated inversion recovery (FLAIR) magnetic resonance imaging (MRI), WMHs appear as bilateral, mostly symmetrical hyperintense lesions, typically located in the periventricular and deep white matter^1^.

The prevalence of WMHs increases with age^2^ but WMHs are also observed in symptomatic and asymptomatic individuals below the age of 50^3^. Cardiovascular risk factors, most notably hypertension, diabetes mellitus (DM) and smoking, are strongly associated with WMHs^4^. Presence of WMHs has also been linked to other risk factors and conditions, such as higher sedentary time and worse kidney function^5^. Classically, WMHs have been associated with increased risk of stroke^4^ and cognitive syndromes^6^. Other dose-dependent relationships between number of WMHs and clinical outcomes have also been described^7^; these include, but are not limited to, depression and apathy^8^, falls^9^, and respiratory and sleep problems^10,11^. In a cross-sectional study of 45,013 UK Biobank (UKB) participants, we confirmed well-established concurrent associations between higher burden of WMHs and a diverse set of clinical phenotypes, including several novel ones, i.e., self-reported eye, hearing, and mouth/dental problems^12^.

WMH volume progression, which is thought to reflect white matter damage due to fluid shifts leading to demyelination and axonal loss, has been widely investigated^13^. While decrease in WMH volume has previously been considered a potential measurement error, recent studies with serial imaging suggest that WMH volume regression is a genuine finding, reflecting the dynamic and at least partially potentially reversible nature of WMHs^14–16^. To illustrate, a systematic review and meta-analysis of longitudinal changes of WMHs including 12,284 participants, and subsequent individual cohort studies, reported WMH volume regression in up to one-third of participants^14^. The mechanisms of WMH volume regression remain insufficiently understood.

While WMH volume progression is well-described, baseline clinical phenotypes associated with subsequent WMH change have not been assessed in large-scale well-phenotyped cohorts. A better understanding of which phenotypes relate to WMH volume change over time is important, as it may help inform potential intervention strategies aimed at reducing WMH burden.

We aimed to identify baseline clinical phenotypes associated with WMH volume change at follow-up in a large multisite UK cohort, and examine which baseline phenotypes help classify individuals into three WMH trajectory groups, namely WMH progressors (participants in whom WMHs increased), WMH regressors (participants in whom WMHs decreased), and WMH stable (participants in whom WMHs remained stable) according to a previously described WMH change categorisation approach^17^, in the specified time period.

## Methods

### Study Population

UKB is a large-scale, population-based, prospective cohort containing detailed genetic, health and lifestyle information on over half a million participants from across the United Kingdom aged 40-69 at baseline when assessed between 2006 and 2010^18^. We included participants who underwent brain imaging at the first brain MRI visit (≥2014), completed a first repeat brain MRI assessment (2019+), and had available information on total volume of WMHs. Details of the brain MRI assessments are available elsewhere^19,20^.

### Ethical Approval

We performed this secondary data analysis study under the generic approval from the NHS National Research Ethics Service, UKB Application No. 17689. All participants provided written informed consent.

### Brain MRI Data

Details of the UKB brain imaging protocol have been described elsewhere^20,21^ and are available at http://www.fmrib.ox.ac.uk/ukbiobank/protocol/V4_23092014.pdf. Briefly, MRI was conducted using a Siemens Skyra 3T scanner with а 32-channel head coil^20^. The protocol includes three structural MRI scans; T1, T2 fluid-attenuation inversion recovery (FLAIR), and susceptibility-weighted MRI^20^. The UKB MRI pipeline (http://www.ukbiobank.ac.uk/expert-working-groups)^20^ processes the brain MRI, performs quality control, and automatically generates imaging-derived phenotypes. Participants started having MRI in 2014 and imaging is planned to continue until 100,000 individuals have been scanned at 1 of 4 assessment centres (Manchester, Newcastle, Reading, and Bristol).

#### White Matter Hyperintensity Volumes

For the purposes of this study, we used total volume of WMHs (in mm^3^), available as an imaging-derived phenotype (UKB data field 25781). Total WMH volumes were calculated based on T1 and T2 FLAIR, derived by UKB using the Brain Intensity Abnormality Classification Algorithm (BIANCA)^22^. For correction purposes in the statistical analysis, we calculated total brain tissue volume as the sum of volume of white matter, volume of grey matter and volume of ventricular cerebrospinal fluid (CSF), as a better measure of intracranial volume including subarachnoid CSF was not available (i.e., our variable thus does not represent the entire space enclosed within the skull).

Prior to running the statistical analysis, we removed outliers using an outlier detection method based on the interquartile range (IQR), rule value>quartile 3+3×IQR (243 outliers; based on WMH volume difference between the two time points; 136 progressors and 107 regressors). We used a stringent outlier removal method as we wanted to ensure inclusion of biologically plausible values^23^.

### Clinical Phenotypes

We assessed 107 previously identified clinical phenotypes based on our cross-sectional work^12^, defined as observable signs and symptoms of individuals with any extent of WMH burden, and broadly classified them into demographic and lifestyle, general health, cognitive, mood and neuropsychiatric, respiratory and sleep, locomotor and physical activity, cardiac and cardiovascular, neurological, renal-urinary, gastrointestinal and digestive health, sensory system, and dental phenotypes, adhering to the International Classification of Functioning, Disability, and Health of the World Health Organization (https://icd.wh.int/dev11/l-icf/en). We used the names of the clinical phenotypes exactly as they appear in the UKB showcase (https://biobank.ndph.ox.ac.uk/showcase/).

We used age at recruitment and self-reported sex from initial assessment visit. For most phenotypes, we used data from first imaging visit (2014+). A detailed list of all clinical phenotypes is presented in Supplementary Material, eAppendix1.

### Statistical Analysis

#### Data Preprocessing using PHESANT

We selected phenotypes of interest and explored the data initially in STATA (v. 18). We summarised and visualised data, and performed all other analyses, using R (v. 4.3.1). We pre-processed variables using the open-source PHESANT analysis tool (https://github.com/MRCIEU/PHESANT) by running PHESANT with the ‘save’ option. A detailed description of the tool is available in Millard et al. (2018)^24^. In short, the decision rules start with the variable field type whereby each variable is categorized into one of four data types: binary, ordered categorical, unordered categorical, or continuous. Variables with the continuous and integer field type are usually assigned to the continuous data type, however, some are assigned to ordered categorical if, for example, there are only a few distinct values. Variables of the categorical (single) field type are assigned to either the binary, ordered categorical or unordered categorical, depending on whether the field has two distinct values, or has been specified as ordered or unordered in PHESANT. Variables of the categorical (multiple) field type are converted to a set of binary variables, one for each value in the categorical (multiple) fields. An inverse normal rank transform (IRNT) is applied to variables of the continuous data type to ensure normal distribution. The derived processed clinical features were stored as CSV files and used in the next stage of the analysis. For more details on PHESANT’s automated rule-based method, see eAppendix 2 in Supplementary Material.

#### Deriving a Measure of WMH Volume Change

To derive a measure of WMH volume change, we regressed WMH volumes at follow-up on baseline WMH volumes, to account for baseline WMH burden, and extracted the residuals as a measure of individual deviation from expected follow-up WMH volumes. These residuals capture WMH volume change beyond what would be predicted from baseline levels. We refer to these residuals as the WMH volume change outcome in the rest of this paper.

#### Analysis of Baseline Clinical Phenotypes Associated with WMH Volume Change

The pre-processed baseline PHESANT phenotypes were grouped into four categories: binary (n=62), ordered categorical (n=29), unordered categorical (n=3), and continuous (n=13). For all phenotypes, we fitted linear regression models with each phenotype as the predictor and standardised WMH volume change as the outcome. For binary predictors, estimates represent the standardized difference in WMH change between individuals with a condition and those without. For ordered categorical predictors, estimates reflect the standardized difference in WMH change per one-step increase in category (for example, data field 20458 (‘General happiness’) ranges from 1=Extremely happy to 6=Extremely unhappy; thus, a positive association indicates greater WMH change with increasing unhappiness). For unordered categorical predictors (i.e., those with more than two categories but no natural ordering), estimates reflect the standardized difference in WMH change compared to the reference category. For continuous predictors, estimates represent the standardized difference in WMH volume change per 1 SD increase in the IRNT predictor^24^.

To examine baseline phenotypes related to subsequent WMH volume change, we conducted (1) unadjusted (predictor only), (2) partially adjusted (age, sex, total brain tissue volume, follow-up time), and (3) fully adjusted (age, sex, total brain tissue volume, follow-up time, and blood pressure) analyses. For each predictor, we calculated regression coefficients, 95% confidence intervals (CI), raw *p-*values, and false discovery rate (FDR)-corrected *p-*values. The FDR method controls the proportion of false positives amongst identified associations (unlike family-wise error rate approaches, which control for the probability that at least one identified association is a false positive, e.g., Bonferroni correction). FDR correction was performed separately for the unadjusted, partially adjusted, and fully adjusted results sets.

#### Comparison between WMH Progression, WMH Regression and WMH Stability

We tested between-group differences in baseline demographic, clinical, and imaging features after categorising participants into three groups – regression, stable, and progression, similar to the approach described by Jochems et al. (2025)^14^. We first determined the percentile corresponding to no WMH change within the sample distribution. This corresponded to the 36^th^ percentile. Stable WMH was then defined as ±10 percentiles around this point (26^th^-46^th^ percentiles), corresponding to a change range of approximately –283 to +207 mm^3^. Participants with WMH decreases below –283 mm^3^ were classified as regressors, and those with WMH increases above +207 mm^3^ as progressors. For approximately normally distributed variables (systolic (SBP) and diastolic blood pressure (DBP), we applied one-way analysis of variance (ANOVA) with Tukey’s honestly significant difference (HSD) test for post hoc pairwise comparisons. For non-normally distributed variables (age, BMI, baseline WMH volume, and follow-up WMH volume), we applied the Kruskal-Wallis test, followed by Dunn’s test with Holm correction for post hoc comparisons. For categorical variables (sex and smoking status), we used Pearson’s χ² tests to assess overall group differences, applying Holm’s correction for multiple testing.

In subsequent analyses, multinomial logistic regression was used to examine the association between clinical phenotypes and the WMH change categories (i.e., with WMH change category as the dependent variable). The stable group was used as the reference. Separate models were fitted for binary, continuous, ordered categorical, and unordered categorical predictors. The same three levels of adjustment were specified. Regression results were expressed as odds ratios (ORs) with 95% CI. *P*-values were FDR-corrected for multiple testing.

## Results

### Clinical Descriptives

A flowchart of participant inclusion is presented in Figure 1. A description of participants with available imaging dates and information on WMH volumes at the two timepoints (N=4329) is presented in Table 1. Median follow-up was 2.3 years (range=1-7 years). Median age was 52 years (IQR=12) and 2362 (54.6%) participants were female. Average (SD) SBP was 138.3 mmHg (18.9) and average (SD) DBP was 78.1 mmHg (10.6).

**Figure 1.**
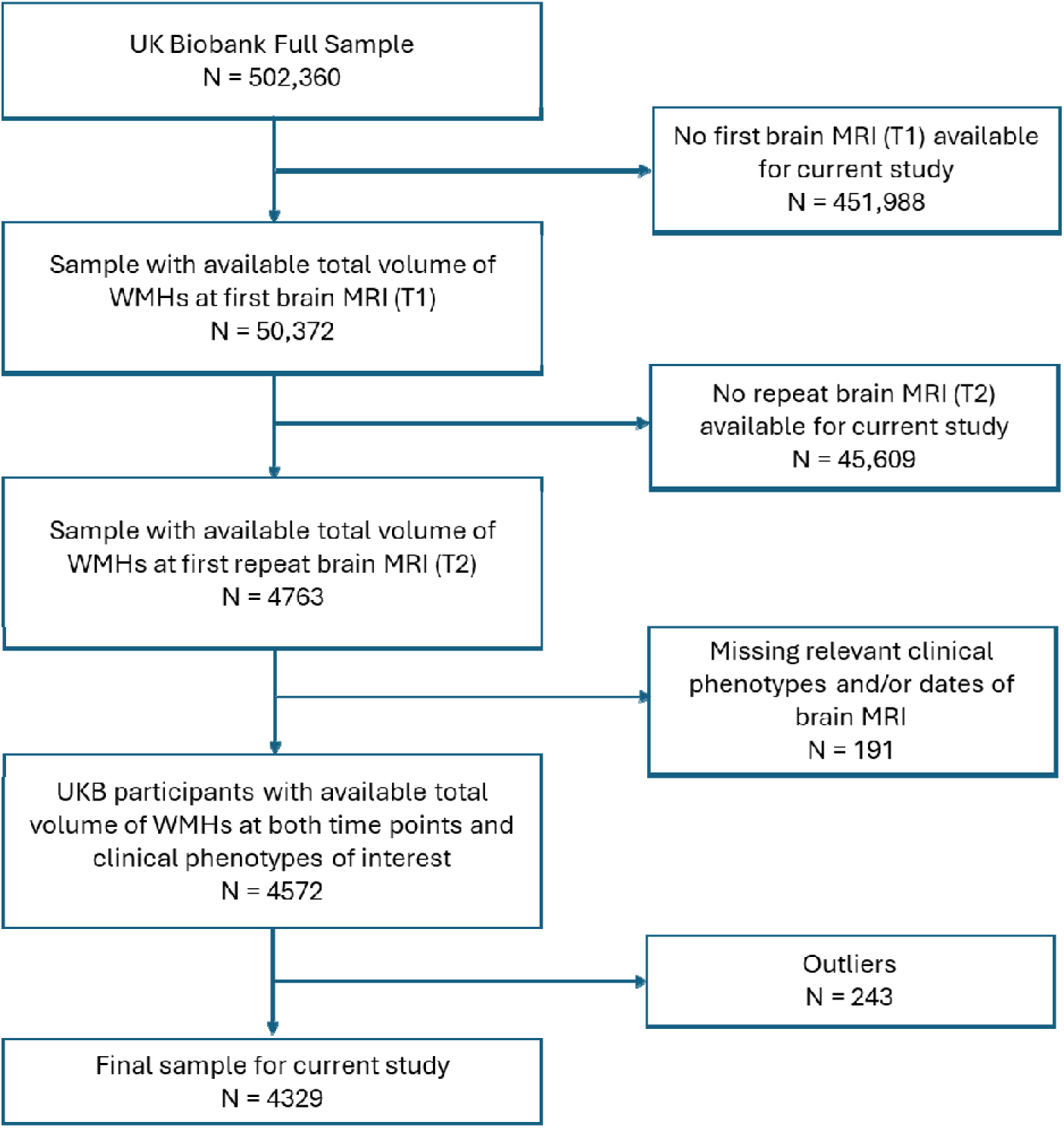
Flowchart of UKB participant inclusion.

**Table 1.**
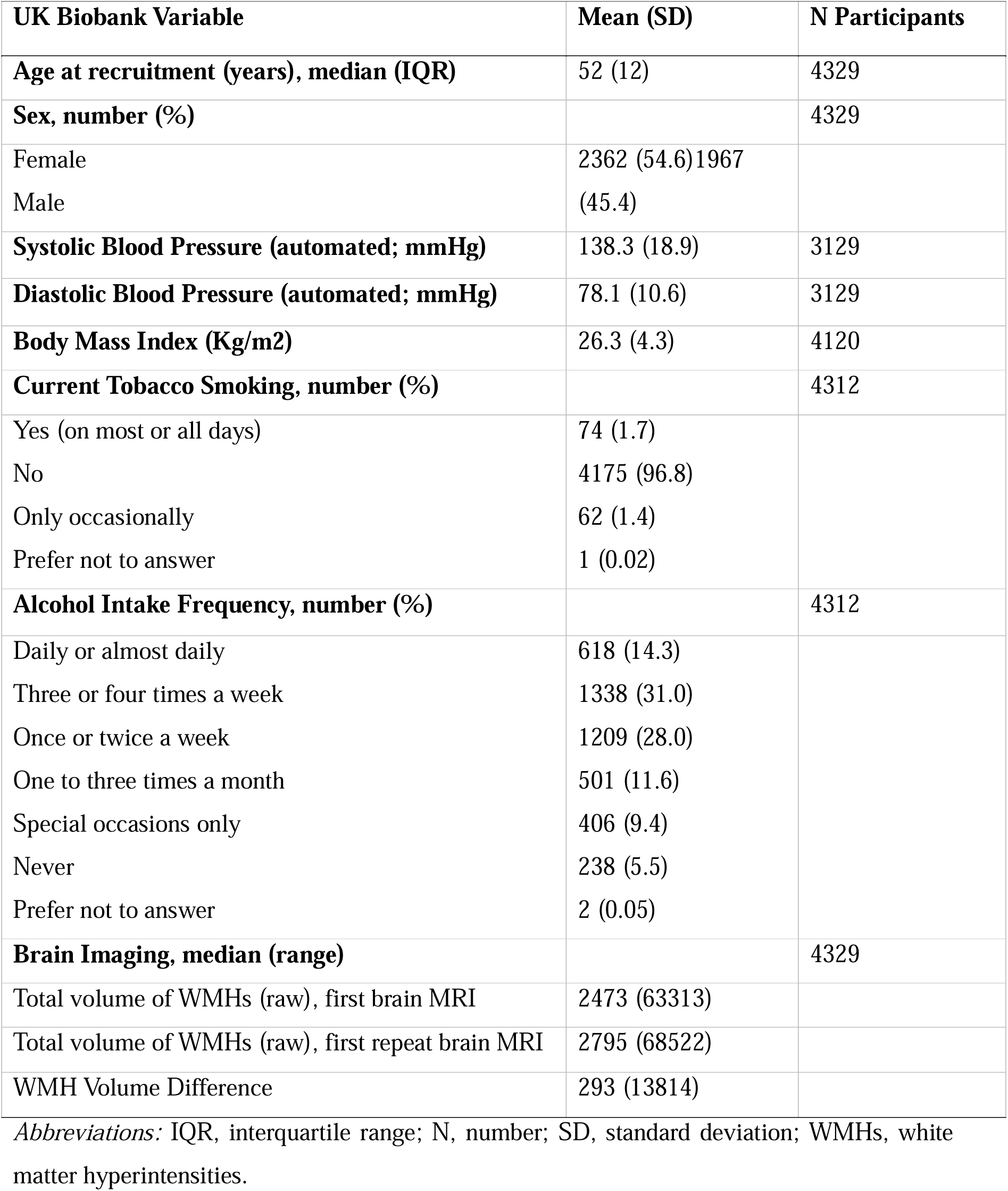
Description of the repeat imaging UKB cohort.

We compared the imaging UKB subsample with first brain MRI but no repeat imaging (N=43,355) to the sample used here (N=4329) using Welch’s *t*-tests or Chi-square tests, as appropriate. Those who underwent repeat imaging were on average slightly younger, had lower SBP, DBP and body mass index (BMI), were more often females, and had lower WMH volumes at baseline (see eAppendix 3 for details).

### Brain Changes Across Follow-up

Mean (SD) total brain volume (grey matter plus white matter) at baseline was higher than at follow-up (1,169,594 mm^3^ [SD=111,645.4] versus 1,153,536 mm^3^ [SD=110,937.1] paired t-test *p*<0.0001). Median (IQR) total volume of WMHs at baseline was 2473mm^3^ (SD=3187) and 2795 mm^3^ (SD=3882) at follow-up.

Across the cohort, WMH volumes increased from baseline to follow-up (median change=+293 mm^3^, IQR: −323 to +1258, Wilcoxon signed-rank test *p*<0.0001). Categorical classification showed that 53.9% of participants exhibited WMH progression, 26.01% regression, and 20.0% remained stable (χ² test *p*<0.0001). Pairwise comparisons confirmed that all groups differed from one another after FDR correction (*p*<0.001 for all comparisons).

Figure 2 displays WMH volume change (in mm^3^) across regression, stable, and progression groups based on the percentile approach. To further visualize the distribution of WMH burden across the spectrum of change, we expressed WMH volume change as quintiles of the overall sample distribution (Q1-Q5). Q1 represented participants with the greatest WMH regression, and Q5 those with the greatest WMH progression. Each quintile contained approximately 20% of the sample (Q1-Q5: N=867, 867, 863, 867, 865; total N=4,329). Baseline WMH volumes (expressed as % total brain tissue volume) were plotted across these quintiles to visualize starting burden differences across the WMH change continuum (see eAppendix 4 for the visualisation).

**Figure 2.**
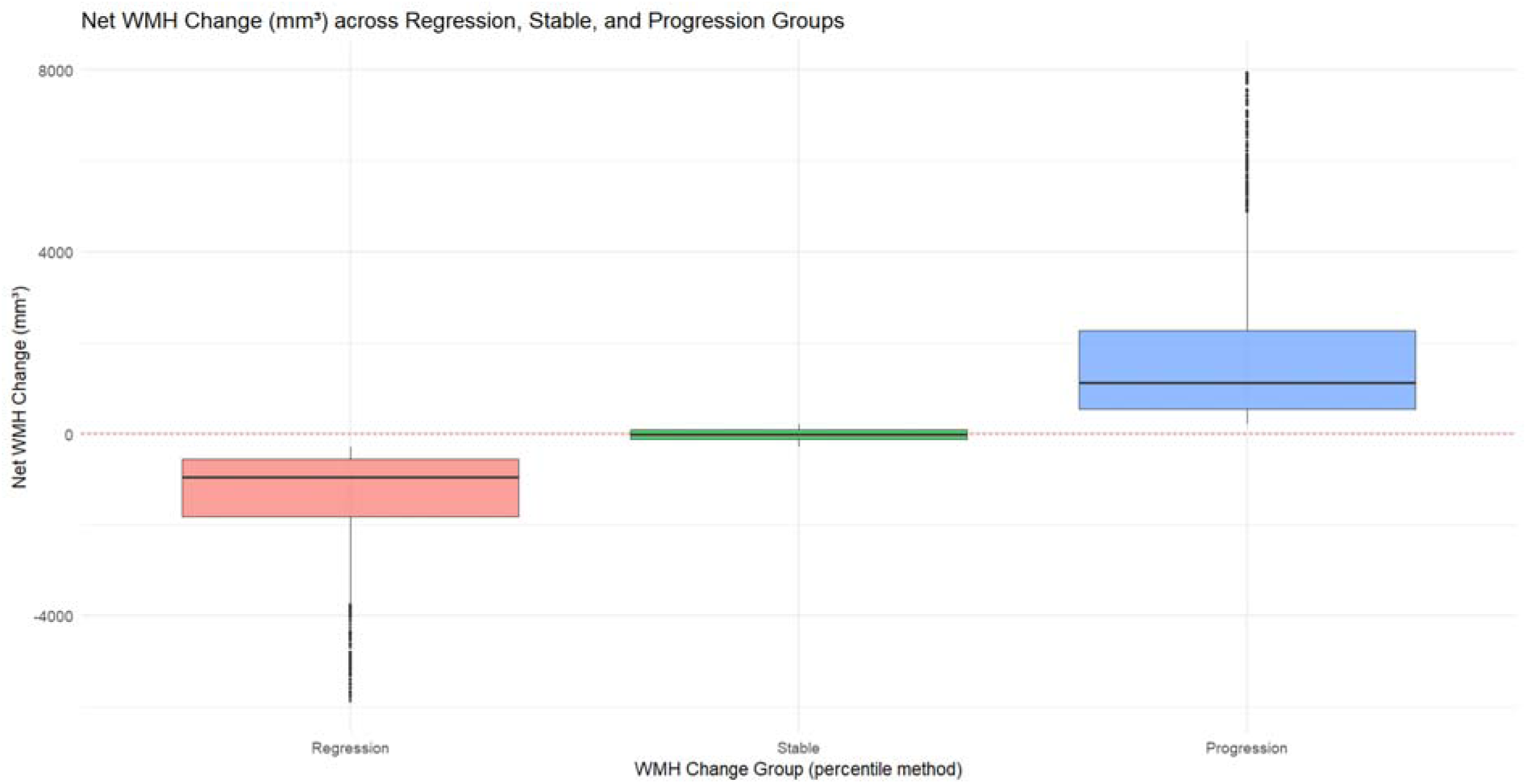
WMH volume change (in mm^3^) across regression, stable, and progression groups based on percentile approach. *Notes:* Boxplots illustrate the distribution of WMH volume change (mm^3^) between baseline and follow-up. Groups were defined using percentile thresholds of the sample distribution of WMH change (regression: WMH decreases below –283 mm^3^; stable: ±10 percentiles around 26th-46th percentiles corresponding to a change range of approximately –283 to +207 mm^3^; progression: WMH increases above +207 mm^3^). Negative values indicate WMH volume regression, and positive values indicate progression. The red dashed line marks zero change.

Longer follow-up duration was also associated with greater WMH progression. Each additional year corresponded to an average increase of ∼204 mm^3^ in WMH volume (as per linear regression model; estimate=203.7, 95%CI [143.5, 264.0], *p*<0.0001). However, the model explained only about 1% of the variance in WMH change (R²=0.01).

#### Baseline Clinical Phenotypes Associated with WMH Volume Change

Of the 107 clinical phenotypes selected for inclusion, we tested associations for 52 of these (some being excluded by PHESANT due to low sample sizes). Table 2 presents all results with *p*<0.05 after FDR correction for associations with any WMH volume change. In unadjusted analyses, several phenotypes were associated with WMH change; to name a few, individuals who reported cataract (0.25SD higher WMH change; 95%CI [0.14, 0.35]), higher SBP (0.08SD higher WMH change per 1SD higher IRNT SBP; 95%CI [0.05, 0.12]), higher DBP (0.07SD higher WMH change per 1SD higher IRNT DBP; 95%CI [0.04, 0.11]), having dentures (0.20SD higher WMH change; 95%CI [0.09, 0.30]), and having longer mean reaction times (0.06SD higher WMH change per 1SD higher IRNT reaction time; 95%CI [0.03, 0.09]) had, on average, greater WMH accumulation at follow-up. In contrast, hip pain lasting over three months (0.35SD lower WMH change; 95%CI [-0.57, -0.13]) and higher frequency of unenthusiasm in last two weeks were associated with lower WMH volume change (0.09SD lower WMH change; 95%CI [-0.15, -0.03]).

**Table 2.**
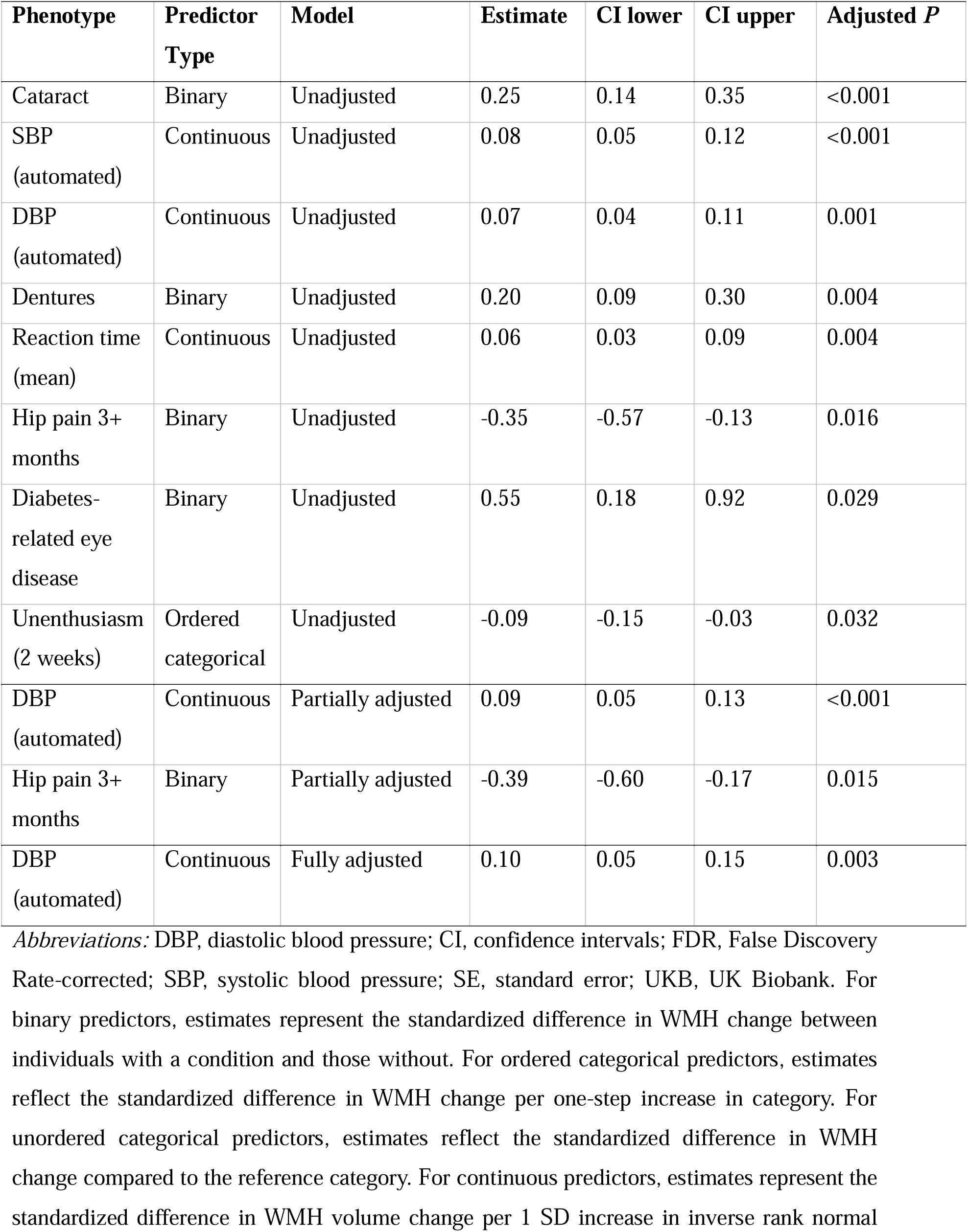
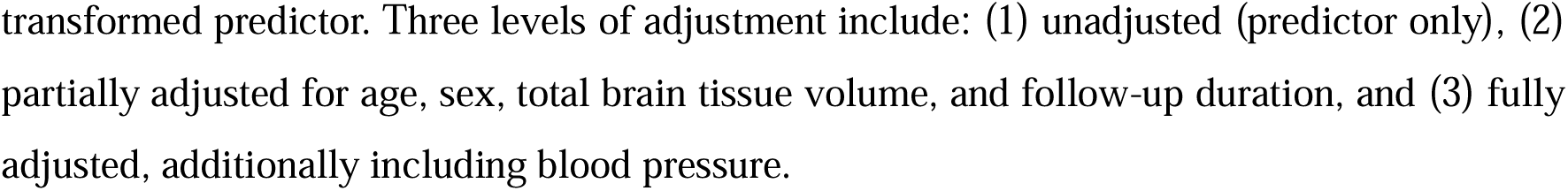
Linear regression results for baseline clinical phenotypes associated with subsequent WMH volume change, with FDR corrected *p*<0.05.

| Phenotype | Predictor Type | Model | Estimate | CI lower | CI upper | Adjusted $P$ |
| --- | --- | --- | --- | --- | --- | --- |
| Cataract | Binary | Unadjusted | 0.25 | 0.14 | 0.35 | <0.001 |
| SBP<br>(automated) | Continuous | Unadjusted | 0.08 | 0.05 | 0.12 | <0.001 |
| DBP<br>(automated) | Continuous | Unadjusted | 0.07 | 0.04 | 0.11 | 0.001 |
| Dentures | Binary | Unadjusted | 0.20 | 0.09 | 0.30 | 0.004 |
| Reaction time<br>(mean) | Continuous | Unadjusted | 0.06 | 0.03 | 0.09 | 0.004 |
| Hip pain 3+<br>months | Binary | Unadjusted | -0.35 | -0.57 | -0.13 | 0.016 |
| Diabetes-related eye<br>disease | Binary | Unadjusted | 0.55 | 0.18 | 0.92 | 0.029 |
| Unenthusiasm<br>(2 weeks) | Ordered<br>categorical | Unadjusted | -0.09 | -0.15 | -0.03 | 0.032 |
| DBP<br>(automated) | Continuous | Partially adjusted | 0.09 | 0.05 | 0.13 | <0.001 |
| Hip pain 3+<br>months | Binary | Partially adjusted | -0.39 | -0.60 | -0.17 | 0.015 |
| DBP<br>(automated) | Continuous | Fully adjusted | 0.10 | 0.05 | 0.15 | 0.003 |

*Abbreviations:* DBP, diastolic blood pressure; CI, confidence intervals; FDR, False Discovery Rate-corrected; SBP, systolic blood pressure; SE, standard error; UKB, UK Biobank. For binary predictors, estimates represent the standardized difference in WMH change between individuals with a condition and those without. For ordered categorical predictors, estimates reflect the standardized difference in WMH change per one-step increase in category. For unordered categorical predictors, estimates reflect the standardized difference in WMH change compared to the reference category. For continuous predictors, estimates represent the standardized difference in WMH volume change per 1 SD increase in inverse rank normal
transformed predictor. Three levels of adjustment include: (1) unadjusted (predictor only), (2) partially adjusted for age, sex, total brain tissue volume, and follow-up duration, and (3) fully adjusted, additionally including blood pressure.

In FDR-corrected (*p*<0.05), partially adjusted analyses, the direction of associations remained consistent. In the fully adjusted models, only the association with DBP survived FDR correction at *p*<0.05.

Figure 3 presents all associations of WMH change for binary, continuous, and ordered categorical predictors separately (see eAppendix 5 for all linear regression results).

**Figure 3.**
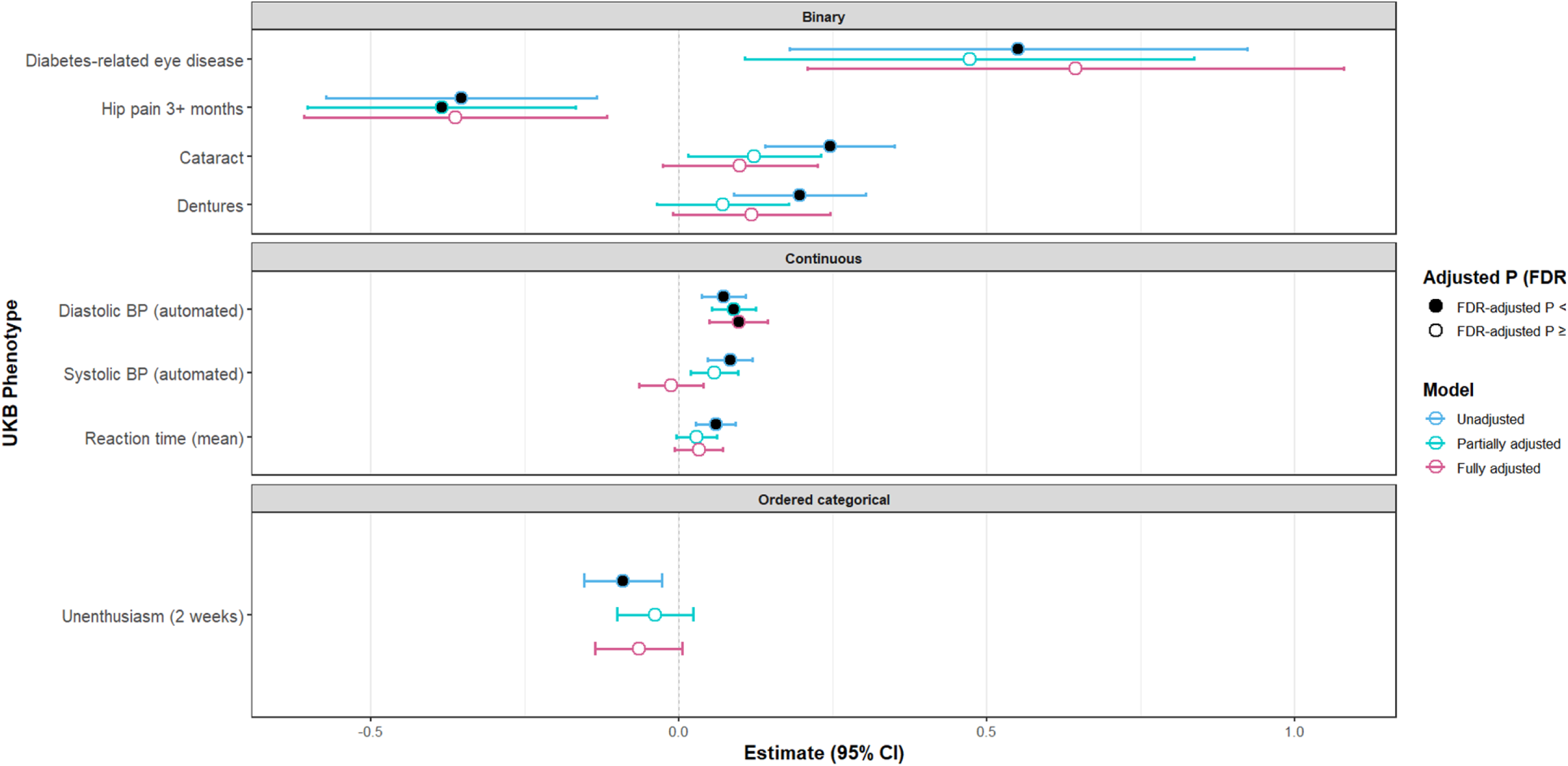
Associations with WMH change for all predictor types across unadjusted, partially adjusted and fully adjusted analyses. *Notes:* Forest plot showing phenotypes associated with WMH volume change across unadjusted, minimally adjusted, and fully adjusted linear regression models. Estimates for binary predictors reflect standardized differences in WMH change per phenotype (±95% CI). Estimates for continuous predictors reflect standardized differences in WMH change per 1SD increase in the predictor (±95% CI). Estimates for ordered categorical predictors reflect standardized differences in WMH change per one-step increase in category (±95% CI). Results are shown for all three model specifications; unadjusted (blue outline), partially adjusted (cyan outline), and fully adjusted (pink outline). Estimates are shown separately by predictor type (binary, continuous, or ordered categorical; unordered categorical predictors did not survive FDR adjustment and are thus not visualised). For each estimate, circle points are used, with the outline colour indicating the adjustment model and the interior fill reflecting FDR correction. Specifically, a solid black fill denotes FDR-adjusted *p*<0.05, while a hollow fill indicates FDR-adjusted *p*≥0.05.

#### Comparison between WMH Progression, WMH Regression and WMH Stability

Differences in age (χ² test *p*<0.001) and BMI (χ² test *p*=0.02) were observed across WMH change groups. Progressors were older than both regressors (Kruskal-Wallis test *p*=0.01) and stable subjects (Kruskal-Wallis test *p*<0.001), while regressors were also older than stable (Kruskal-Wallis test *p*<0.001). Both regressors (Kruskal-Wallis test *p*=0.02) and progressors (Kruskal-Wallis test *p*=0.04) had higher BMI than the stable group. SBP and DBP also differed by group (ANOVA test for SBP *p*<0.0001; ANOVA test for DBP *p*=0.03) whereby stable participants had lower SBP compared to both regressors and progressors at *p*<0.0001, with no difference between regressors and progressors. Progressors had higher DBP than stable (ANOVA test *p*=0.024), whereas little evidence of an association was observed for other DBP contrasts; all *p* FDR-corrected.

Baseline WMH volumes differed markedly between the three groups (χ² test *p*<0.0001). Both progressors and regressors had higher WMH volumes than the stable group (median WMH volume for progressors=2474 mm^3^ (IQR=3291) versus median WMH volume for regressors=3566.5 mm^3^ (IQR=3928.8) versus median WMH volume for stable 1582.5 mm^3^ (IQR=1592.8), *p*<0.0001). Notably, regressors had higher baseline WMH burden compared to progressors (Kruskal-Wallis test *p*<0.0001). At follow-up, WMH volumes also differed across groups (χ² test *p*<0.0001). WMH progressors had markedly higher WMH volumes (median=3998 mm^3^, IQR=4949) than both regressors (median=2213, IQR=2747.8) and stable participants (median=1538.5, IQR=1597); *p*<0.0001. Regressors also had higher WMH volumes than the stable group at follow-up (Kruskal-Wallis test *p*<0.0001). All presented *p* values are FDR-corrected. For full details on the comparison between WMH progressors, WMH regressors, and WMH stable, see eAppendix 6.

Results from the multinomial regression analyses after FDR correction are presented in Table 3 and Figure 4 for unadjusted, partially and fully adjusted models (see eAppendices 7 and 8 for all associations before and after FDR correction). In unadjusted analyses, individuals in the progression group were more likely to report cataract compared to stable participants (OR=1.91, 95%CI [1.39, 2.62]), headaches lasting more than three months (OR=1.77, 95%CI [1.14, 2.75]), as well as higher manual SBP (OR=1.32 per 1SD higher IRNT SBP, 95%CI [1.08, 1.60]), higher automated SBP (OR=1.31 per 1SD higher IRNT SBP, 95%CI [1.18, 1.44]), and higher automated DBP (OR=1.13 per 1SD higher IRNT DBP, 95%CI [1.03, 1.25]). In contrast, better puzzle performance was inversely related to WMH progression versus stability (OR=0.95 per 1 category higher puzzle performance, 95%CI [0.91, 0.99]). Regarding regression, higher body weight (OR=1.20 per 1SD higher IRNT body weight, 95%CI [1.10, 1.32]) and higher BMI (OR=1.14 per 1SD higher IRNT BMI, 95%CI [1.03, 1.25]) were both linked to higher probability of regression compared to stability. Higher manual SBP was also related to higher odds of WMH regression versus stability (OR=1.29 per 1SD higher IRNT SBP, 95%CI [1.04, 1.60]); all *p*<0.05 FDR-corrected.

**Figure 4.**
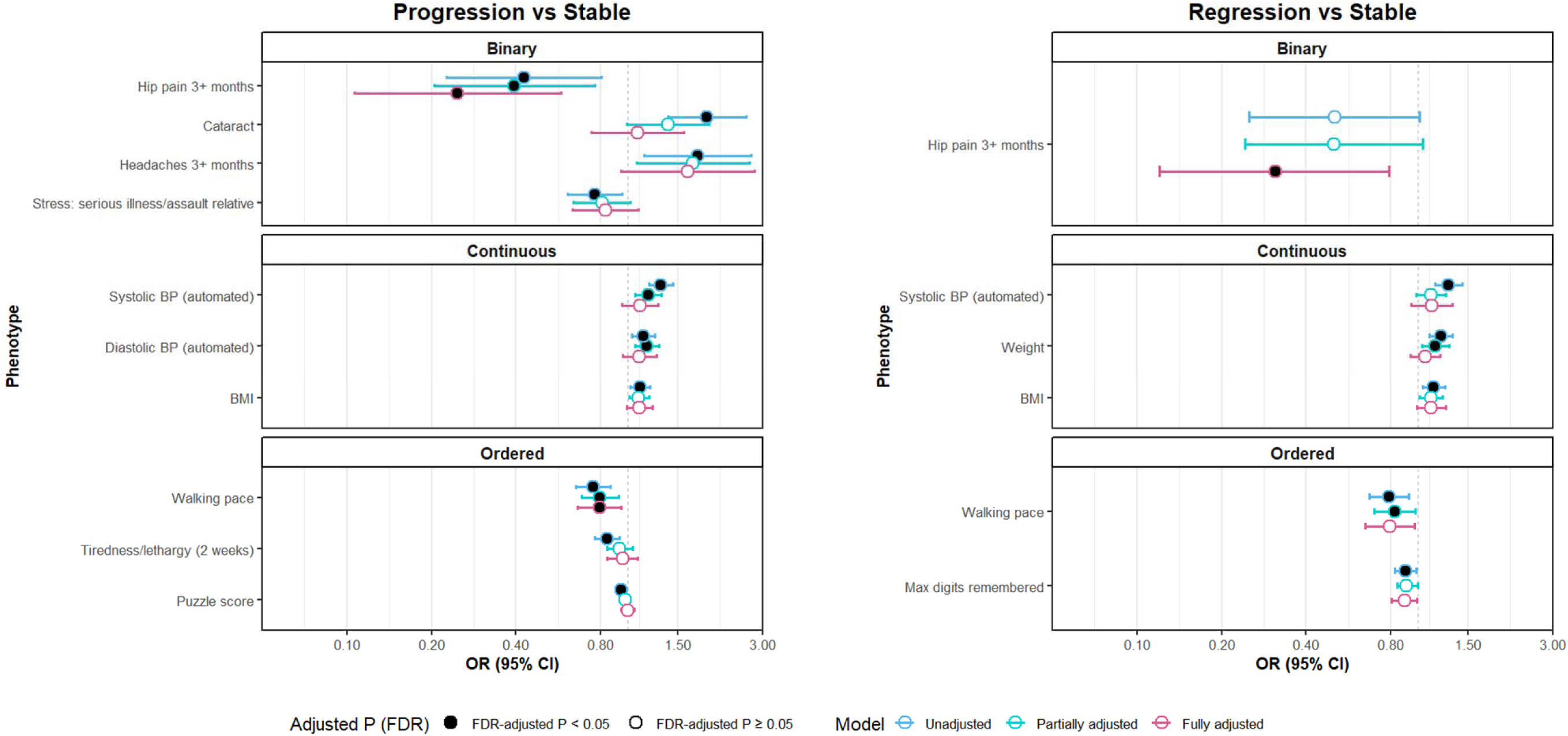
Clinical phenotypes associated with subsequent WMH volume change according to group membership (progression, regression, or stable). *Notes:* Forest plot showing phenotypes associated with WMH volume change across unadjusted, minimally adjusted, and fully adjusted multinomial logistic regression models. For binary predictors, ORs represent the odds of group membership for participants with the exposure (value=1) versus without (value=0). The dashed vertical line marks the value OR=1. For continuous predictors, ORs reflect the change in odds per 1SD increase (inverse-normal transformed). For ordered categorical predictors, ORs represent the change in odds per category increase. Unordered categorical predictors did not survive FDR adjustment and are thus not visualised. ORs greater than 1 indicate higher odds of belonging to the respective WMH change group compared with the stable group, and ORs less than 1 indicate lower odds. Results are shown for all three model specifications; unadjusted (blue outline), partially adjusted (cyan outline), and fully adjusted (pink outline). Estimates are shown separately by predictor type. For each estimate, circle points are used, with the outline colour indicating the adjustment model and the interior fill reflecting FDR correction. Specifically, a solid black fill denotes FDR-adjusted *p*<0.05, while a hollow fill indicates FDR-adjusted *p*≥0.05.

**Table 3.**
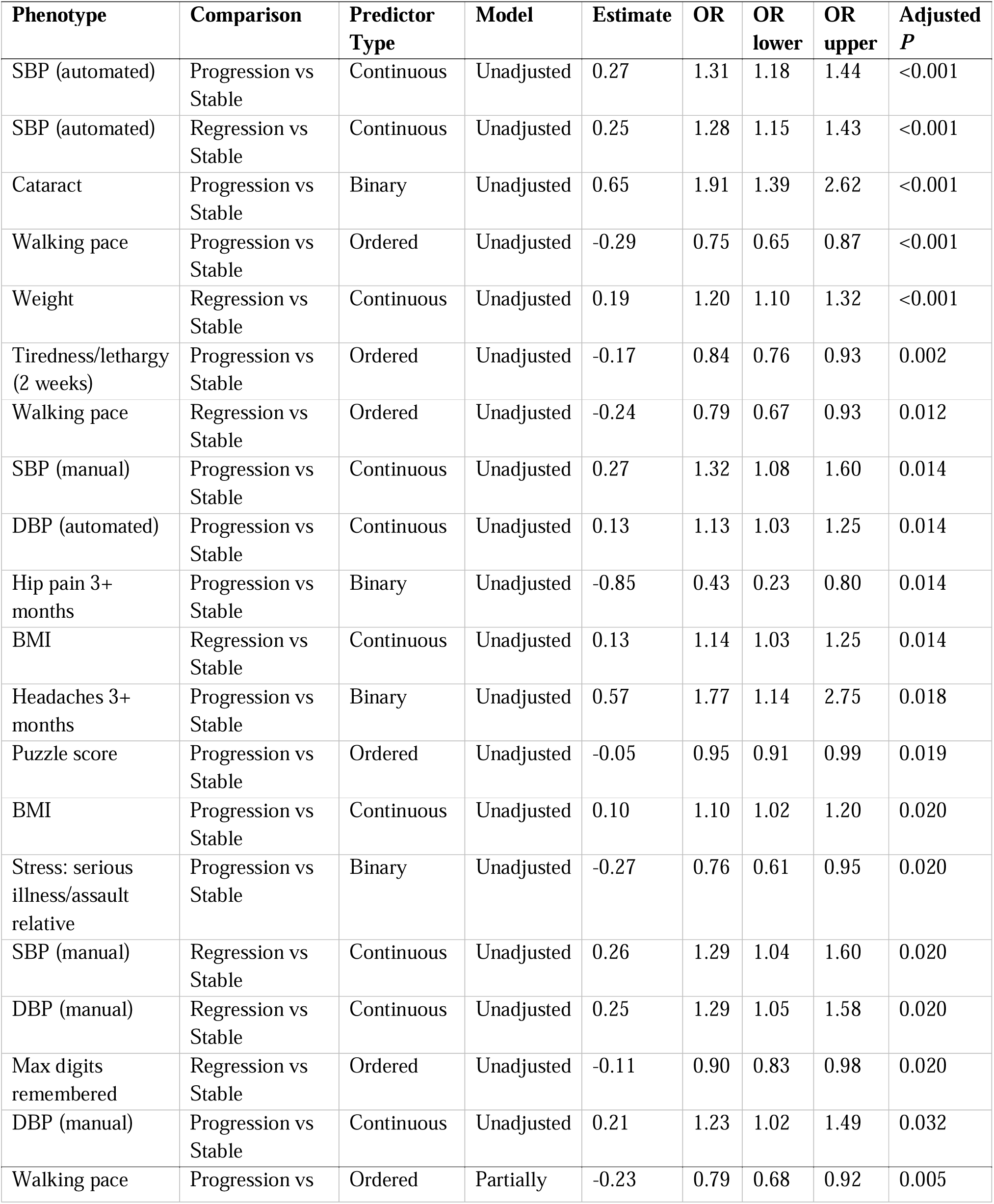
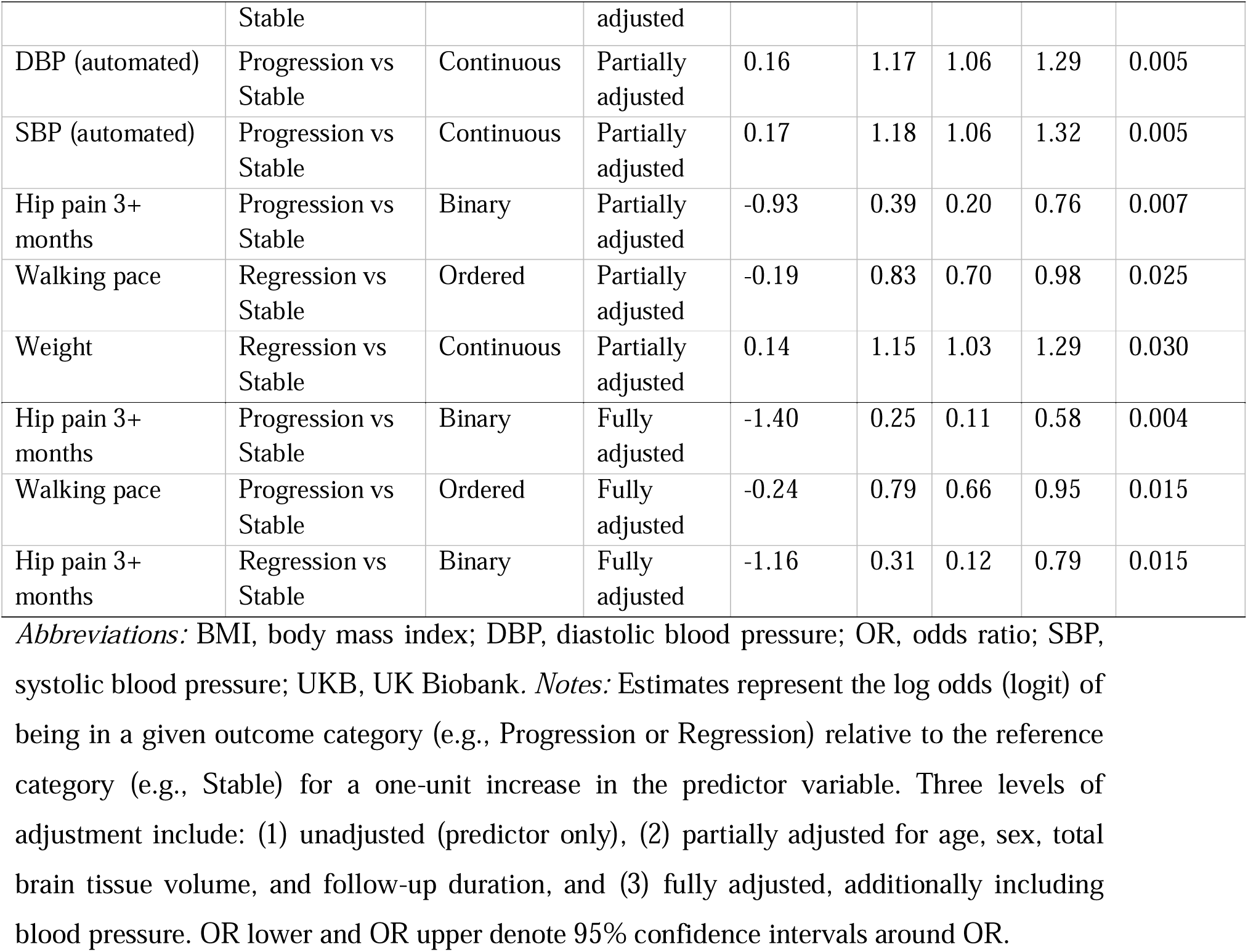
Multinomial logistic regression results for associations between clinical phenotypes and WMH change categories (progression, regression, stable; with stable as the reference), with FDR corrected *p*<0.05.

| Phenotype | Comparison | Predictor Type | Model | Estimate | OR | OR lower | OR upper | Adjusted *P* |
| --- | --- | --- | --- | --- | --- | --- | --- | --- |
| SBP (automated) | Progression vs Stable | Continuous | Unadjusted | 0.27 | 1.31 | 1.18 | 1.44 | <0.001 |
| SBP (automated) | Regression vs Stable | Continuous | Unadjusted | 0.25 | 1.28 | 1.15 | 1.43 | <0.001 |
| Cataract | Progression vs Stable | Binary | Unadjusted | 0.65 | 1.91 | 1.39 | 2.62 | <0.001 |
| Walking pace | Progression vs Stable | Ordered | Unadjusted | -0.29 | 0.75 | 0.65 | 0.87 | <0.001 |
| Weight | Regression vs Stable | Continuous | Unadjusted | 0.19 | 1.20 | 1.10 | 1.32 | <0.001 |
| Tiredness/lethargy (2 weeks) | Progression vs Stable | Ordered | Unadjusted | -0.17 | 0.84 | 0.76 | 0.93 | 0.002 |
| Walking pace | Regression vs Stable | Ordered | Unadjusted | -0.24 | 0.79 | 0.67 | 0.93 | 0.012 |
| SBP (manual) | Progression vs Stable | Continuous | Unadjusted | 0.27 | 1.32 | 1.08 | 1.60 | 0.014 |
| DBP (automated) | Progression vs Stable | Continuous | Unadjusted | 0.13 | 1.13 | 1.03 | 1.25 | 0.014 |
| Hip pain 3+ months | Progression vs Stable | Binary | Unadjusted | -0.85 | 0.43 | 0.23 | 0.80 | 0.014 |
| BMI | Regression vs Stable | Continuous | Unadjusted | 0.13 | 1.14 | 1.03 | 1.25 | 0.014 |
| Headaches 3+ months | Progression vs Stable | Binary | Unadjusted | 0.57 | 1.77 | 1.14 | 2.75 | 0.018 |
| Puzzle score | Progression vs Stable | Ordered | Unadjusted | -0.05 | 0.95 | 0.91 | 0.99 | 0.019 |
| BMI | Progression vs Stable | Continuous | Unadjusted | 0.10 | 1.10 | 1.02 | 1.20 | 0.020 |
| Stress: serious illness/assault relative | Progression vs Stable | Binary | Unadjusted | -0.27 | 0.76 | 0.61 | 0.95 | 0.020 |
| SBP (manual) | Regression vs Stable | Continuous | Unadjusted | 0.26 | 1.29 | 1.04 | 1.60 | 0.020 |
| DBP (manual) | Regression vs Stable | Continuous | Unadjusted | 0.25 | 1.29 | 1.05 | 1.58 | 0.020 |
| Max digits remembered | Regression vs Stable | Ordered | Unadjusted | -0.11 | 0.90 | 0.83 | 0.98 | 0.020 |
| DBP (manual) | Progression vs Stable | Continuous | Unadjusted | 0.21 | 1.23 | 1.02 | 1.49 | 0.032 |
| Walking pace | Progression vs | Ordered | Partially | -0.23 | 0.79 | 0.68 | 0.92 | 0.005 |
|  | Stable |  | adjusted |  |  |  |  |  |
| DBP (automated) | Progression vs Stable | Continuous | Partially adjusted | 0.16 | 1.17 | 1.06 | 1.29 | 0.005 |
| SBP (automated) | Progression vs Stable | Continuous | Partially adjusted | 0.17 | 1.18 | 1.06 | 1.32 | 0.005 |
| Hip pain 3+ months | Progression vs Stable | Binary | Partially adjusted | -0.93 | 0.39 | 0.20 | 0.76 | 0.007 |
| Walking pace | Regression vs Stable | Ordered | Partially adjusted | -0.19 | 0.83 | 0.70 | 0.98 | 0.025 |
| Weight | Regression vs Stable | Continuous | Partially adjusted | 0.14 | 1.15 | 1.03 | 1.29 | 0.030 |
| Hip pain 3+ months | Progression vs Stable | Binary | Fully adjusted | -1.40 | 0.25 | 0.11 | 0.58 | 0.004 |
| Walking pace | Progression vs Stable | Ordered | Fully adjusted | -0.24 | 0.79 | 0.66 | 0.95 | 0.015 |
| Hip pain 3+ months | Regression vs Stable | Binary | Fully adjusted | -1.16 | 0.31 | 0.12 | 0.79 | 0.015 |

*Abbreviations:* BMI, body mass index; DBP, diastolic blood pressure; OR, odds ratio; SBP, systolic blood pressure; UKB, UK Biobank. *Notes:* Estimates represent the log odds (logit) of being in a given outcome category (e.g., Progression or Regression) relative to the reference category (e.g., Stable) for a one-unit increase in the predictor variable. Three levels of adjustment include: (1) unadjusted (predictor only), (2) partially adjusted for age, sex, total brain tissue volume, and follow-up duration, and (3) fully adjusted, additionally including blood pressure. OR lower and OR upper denote 95% confidence intervals around OR.

In partially adjusted analyses, (n=4201 participants with complete data), a small number of phenotypes were associated with WMH progression versus stability with FDR-corrected *p*<0.05. Walking pace was inversely associated with both WMH progression (OR=0.79 per 1 category higher walking pace, 95%CI [0.68, 0.92]) and regression (OR=0.83, 95%CI [0.70, 0.98]). Higher automated DBP (OR=1.17 per 1SD higher IRNT DBP, 95%CI [1.06, 1.29]) and higher automated SBP were also associated with greater odds of WMH progression (OR=1.18 per 1SD higher IRNT SBP, 95%CI [1.06, 1.32]). Hip pain lasting more than three months was inversely related to WMH progression (OR=0.39, 95%CI [0.20, 0.76]); all *p*<0.05 FDR-corrected.

Results after full adjustment (n=3038 participants with complete data) were similar to the partially adjusted results. Hip pain lasting more than three months (OR=0.25, 95%CI [0.11, 0.58]) and faster walking pace (OR=0.79, 95%CI [0.66, 0.95]) were inversely related to WMH progression; hip pain lasting more than three months was also inversely associated with WMH regression versus stability (OR=0.31, 95%CI [0.12, 0.79]); all *p*<0.05 FDR-corrected

## Discussion

We investigated baseline phenotypes associated with subsequent WMH volume change in 4329 UKB participants, and examined which phenotypes relate to WMH progression, regression, or stability. This study builds on our previous work systematically reviewing clinical phenotypes associated with cSVD, and exploring concurrent associations between WMH volume and clinical phenotypes in UKB^12,25^. We demonstrate that established vascular risk factors relate to subsequent WMH volume change. Distinct clinical and demographic profiles seemed to characterize WMH progression, regression, and stable groups, but results require further corroboration.

WMH volume change (overall positive across follow-up) was associated with well-established lifestyle and cardiovascular (CV) factors, including SBP and DBP, in line with previous work^12,14,26^. Longitudinal evidence is less consistent, but associations with increasing age, hypertension, and diabetes have been widely corroborated^27,28^. WMH volume progression was also related to slower reaction times, and better performance on the puzzle solving task was inversely related to WMH progression versus stability, extending established associations between higher WMH burden and poorer executive function and processing speed^29^. Additionally, the finding that individuals in the progression group were more likely to report persistent headaches compared to stable participants supports previous reports of a link between headache prevalence and vascular disorders^30^, where elevated BP can increase pressure within cerebral blood vessels, leading to vascular changes that may manifest as headache. However, this association did not remain significant after covariate adjustment.

The inverse association with unenthusiasm contradicts our previous cross-sectional findings and other reports that higher WMH burden relates to apathy^12,31^. One possible explanation is that baseline unenthusiasm may not directly reflect progression of early cerebrovascular damage but could relate to other neuropsychiatric or systemic factors instead.

Further, we found an association of any subsequent WMH volume change with cataract. In our previous UKB study, diabetes-related eye disorders, including cataract, were concurrently associated with higher WMH burden^12^. In the context of cSVD, this might reflect unaccounted effects of age, or that cataract is a proxy of poor glycaemic control, which is mechanistically related to age-related modification, aggregation, and precipitation of aberrant proteins, alongside oxidative stress^32,33^. Of note, residual confounding by age is likely, as statistical adjustment may only partially account for age effects.

The association between dentures and higher WMH burden also extends our previous work^12^. Poor oral health is common in dementia^34^ and can be due to reduced capacity to maintain personal hygiene^35^, motor and behavioural disturbances^35,36^, but also, is more common with advancing age, smoking, and poor glycaemic control. While our findings require further corroboration and could be due to confounding by age, they underscore the importance of regular oral health screening, particularly given that failure to act on oral health might impact on other aspects of general health, including stress, pain management, and psychosocial wellbeing^37^.

Higher DBP consistently related to worse WMHs in fully adjusted analyses. While large BP variability has been linked to higher WMH burden and worse cognitive outcomes^38^, evidence clarifying the impact of different BP-related metrics separately is still scarce. Importantly, the association between BP and WMHs might differ depending on the location of WMHs^39^. At the base of the brain, short straight arteries transmit BP directly to small resistance vessels; in that vicinity, high BP is thought to directly damage small arterioles. However, these small arterioles are supplied by long arteries with many branches, and so BP in them is typically lower^39^. Related to that, periventricular and subcortical WMHs are probably somewhat differently impacted by BP, and may have different aetiologies^39^. While we were unable to assess different metrics related to BP in this work, going forward, the field would benefit from a more detailed analysis of the individual contribution of SBP, DBP, BP variability, and pulse rate/pressure to vascular brain health.

Regarding group comparisons, higher age, BP and BMI were associated with higher likelihood of WMH progression versus stability and regression, in line with other studies^14^. Interestingly, higher baseline BP, weight and BMI also related to higher odds of regression versus stability in our study, but we did not have information as to whether weight and BMI changed across follow-up. There are a few potential explanations for these findings, for example, that fluctuations in BP (i.e., high variability rather than chronic elevation) may intermittently improve perfusion and clearance, possibly leading to transient reductions in WMH volume in select regions^40^. Importantly, we did not have BP measures at both timepoints and could not assess whether BP changed over time. Thus, our findings cannot be used to draw conclusions about the potential effect of increasing or decreasing BP over time on the likelihood of WMH progression or regression.

There are several potential explanations for the inverse relationships found between hip pain, faster walking pace, and WMH progression and regression, as observed in our results after adjustment for confounders. Pain often leads to reduced mobility, greater sedentary behaviour, and/or less overall physical activity, which are established risk factors for WMH progression^41^. Of note, UKB participants are middle-aged and healthy, and those who underwent follow-up brain MRI are likely to represent a subgroup with better physical health^42^. Thus, the inverse relationship observed for WMH progression versus stability is likely attributable to confounding, selection bias, or other unmeasured factors. The disappearance of the association with WMH regression after covariate adjustment further suggests residual confounding; additionally, regressors had higher baseline WMH burden than stable participants, so slower walking speed in this group is not unexpected.

The finding that faster walking pace was inversely related to both progression and regression is interesting. Faster walking pace is a marker of overall vascular and neurological health^43^. Our results could be taken to suggest that those with better mobility might have lower odds of being in groups with dynamic WMH changes, regardless of the direction. Interestingly, the association only remained significant after full adjustment for confounders when comparing only progressors with stable participants, suggesting that progressors had more established WMH damage, which is less likely to reverse. In this context, poor walking speed could serve as a marker of increased risk for WMH progression, although further work is required to substantiate this. The stable group in our study may also have been enriched for the healthiest participants, although our descriptive comparisons did not strongly indicate this. Overall, the associations observed for prolonged hip pain and faster walking pace may reflect the multifaceted influence of activity, resilience, and neurovascular health on white matter, distinct from traditional vascular risk pathways. Nonetheless, these findings require further corroboration before firmer conclusions can be drawn.

Last but not least, in our study, participants in the WMH regression group were those who had the highest baseline WMH burden. There are various potential explanations for these results. Some studies note that part of the regression seen in WMH volume may be attributable to factors, such as MRI scan variability, thresholding methods, or differences in hydration or inflammation at the time of scan, and that true biological regression is rarer but does occur^44^. Lesions due to transient changes (e.g., perivascular oedema or inflammation) may also resolve over time, appearing as WMH regression on imaging^45^. Importantly, altered interstitial fluid mobility, which may be reversible, is thought to predate demyelination and axonal damage, which are less likely to be reversible and are probably a late-stage phenomenon^46^. Finally, it is worth mentioning that regression is not always benign; factors underlying lesion shrinkage may reflect compensatory changes related to prior damage. Also, preserved or even improved cognitive and executive performance in regressors has not been extensively documented and requires further study^17^. Of note, we did not check the raw brain images, so could not determine factors related to the observed WMH volume regression in this study.

The main strength of our study is that it contributes to a growing body of research assessing longitudinal WMH change in a relatively large sample of participants. Including WMH volume regression in addition to progression is another strength, a phenomenon that remains poorly understood. Elucidating the factors contributing to WMH regression can help pave the way for new approaches to understanding cSVD pathology and potentially designing new intervention strategies, which consider specific risk factors and/or groups of individuals.

Limitations of our study need to be acknowledged. A key limitation of our approach is that the majority of clinical phenotypes were only available at baseline, and we therefore examined their associations with subsequent WMH volume change using these values. A purely longitudinal design would have required repeated measures of both phenotypes and WMHs at baseline and follow-up. The follow-up period in this study was also relatively short, reducing statistical power to identify associations with WMH burden.

This study used a small subsample of the whole UK Biobank cohort. The sample was young (median age=52 years), which is well below the age at which WMHs typically become more prominent^47^. WMH volumes were also very small (median 2.5ml), approximately half the volumes observed in community-dwelling cohorts at age 65 or in participants at the start of the SPRINT MIND study^47,48^. Therefore, the associations observed with WMH change may not generalise to older populations or to individuals with more severe WMH burden^49^.

UKB participants are representative of the UK population in terms of age and sex, but are healthier, more educated, and do not represent socio-economically deprived populations^42,50^. Those who underwent repeat brain imaging are also healthier compared to participants who did not^50^. Importantly, our analysis did not account for WMH location^51^ and we did not perform visual checks of the WMH lesion masks. Further, we could not explore other potentially relevant WMH associations, such as with incontinence or renal function, due to unavailability of those data from UKB. The total brain tissue volume variable that we used did not include subarachnoid CSF and, as such, is not a proper proxy for ICV. Thus, our adjustment will not have corrected for original brain size as effectively as possible, should a better measure of ICV were available. In order to test associations across several phenotypes, we used three analysis versions adjusting for different confounders (unadjusted, partially adjusted and fully adjusted) as it was not practical to determine potential confounders for each phenotype separately. This means that we may not have included the most appropriate set of potential confounders for each phenotype. We cannot exclude reverse causation and it is also possible that some of the identified associations are due to collider bias^42^. It remains possible that unknown or unmeasured variables could have contributed residual confounding.

Previous studies have shown that baseline level of WMHs predicts WMH progression^27^. In our sample, WMH regressors had a higher baseline volume of WMHs, contrary to other work. Importantly, WMH regression should not unequivocally be considered a marker of improved brain health, as some studies have observed WMH regression in the context of increased total brain volume loss^52^. Future research should focus efforts on improving understanding of the clinical significance of WMH regression in the context of systemic health. In this regard, more longitudinal studies in diverse, more representative populations and using longer follow-up times would be paramount.

Our results have implications for both research and clinical practice. If WMHs are not simply markers of irreversible damage, interventions aimed at reducing or stabilizing WMH burden can be designed going forward. These may include lifestyle modifications, vascular risk management, and pharmacological interventions. Future work should aim to replicate associations that require further study and collect information on understudied clinical variables where possible. Potential differences between groups with progressing, regressing, or stable WMHs should also be further examined in future work.

## Conclusions

We investigated a wide range of baseline clinical phenotypes associated with subsequent WMH volume change in a sub-sample of UKB imaging study, and examined which baseline phenotypes differed between individuals with progressing, regressing, or stable WMHs. Well-established vascular risk factors related to subsequent WMH volume change. Distinct clinical and demographic profiles characterized WMH progression, regression, and stable groups, but further work is needed to establish factors differentially related to WMH change trajectories.

## Data Availability

All data produced in the present study are available upon reasonable request to the authors.

