## Supplements for "Associations of Baseline Clinical Phenotypes with White Matter Hyperintensity Volume Change – A Study of 4329 UK Biobank Participants"

**SUPPLEMENTARY MATERIAL – eAppendices**

**CONTENTS**

**eAppendix 1.** An overview of UK Biobank (UKB) variables with corresponding data fields and data types as categorised by PHESANT.

**eAppendix 2.** PHESANT Phenome Scan Tool.

**eAppendix 3.** Comparison between UKB imaging cohort with first brain MRI but no repeat imaging and current sample.

**eAppendix 4.** Baseline WMH (%total brain tissue volume) across quintiles of WMH volume change.

**eAppendix 5.** All linear regression results for baseline clinical phenotypes associated with subsequent WMH volume change.

**eAppendix 6.** A comparison between WMH progressors, WMH regressors, and WMH stable (N=4329).

**eAppendix 7.** All unadjusted multinomial regression results for baseline clinical phenotypes associated with subsequent WMH volume change not surviving FDR-correction at *p*<0.05.

**eAppendix 8.** All partially and fully adjusted multinomial regression results for baseline clinical phenotypes associated with subsequent WMH volume not surviving FDR-correction at *p*<0.05.

**eAppendix 1.** An overview of UK Biobank (UKB) variables with corresponding data fields and data types as categorised by PHESANT.

| **UKB Data Field and PHESANT type** | **Clinical Phenotype (label)** |
| --- | --- |
| ***Binary*** |  |
| 1210 (imaging visit (2014+)) | Snoring |
| 1920 (imaging visit (2014+)) | Mood swings |
| 1930 (imaging visit (2014+)) | Miserableness |
| 1940 (imaging visit (2014+)) | Irritability |
| 1950 (imaging visit (2014+)) | Sensitivity/hurt feelings |
| 1960 (imaging visit (2014+)) | Fed-up feelings |
| 1970 (imaging visit (2014+)) | Nervous feelings |
| 1980 (imaging visit (2014+)) | Worrier/anxious |
| 1990 (imaging visit (2014+)) | Tense/highly strung |
| 2000 (imaging visit (2014+)) | Worry too long after embarrassment |
| 2010 (imaging visit (2014+)) | Suffer from nerves |
| 2020 (imaging visit (2014+)) | Loneliness/isolation |
| 2030 (imaging visit (2014+)) | Guilty feelings |
| 2040 (imaging visit (2014+)) | Risk taking |
| 2188 (imaging visit (2014+)) | Long-standing illness/disability |
| 2207 (imaging visit (2014+)) | Wears glasses/contacts |
| 2227 (imaging visit (2014+)) | Other eye problems |
| 2316 (imaging visit (2014+)) | Wheeze in chest |
| 2335 (imaging visit (2014+)) | Chest pain/discomfort |
| 2443 (imaging visit (2014+)) | Diabetes diagnosed |
| 2956 (imaging visit (2014+)) | General pain 3+ months |
| 3005 (imaging visit (2014+)) | Fracture from fall |
| 3393 (imaging visit (2014+)) | Hearing aid user |
| 3404 (imaging visit (2014+)) | Neck/shoulder pain for 3+ months |
| 3414 (imaging visit (2014+)) | Hip pain for 3+ months |
| 3571 (imaging visit (2014+)) | Back pain for 3+ months |
| 3606 (imaging visit (2014+)) | Chest pain when walking |
| 3741 (imaging visit (2014+)) | Abdominal pain for 3+ months |
| 3799 (imaging visit (2014+)) | Headaches for 3+ months |
| 4067 (imaging visit (2014+)) | Facial pain for 3+ months |
| 4598 (imaging visit (2014+)) | Ever depressed for a whole week |
| 4631 (imaging visit (2014+)) | Ever disinterested for a whole week |
| 4642 (imaging visit (2014+)) | Ever hyper/manic for 2 days |
| 4653 (imaging visit (2014+)) | Ever highly irritable |
| 4717 (imaging visit (2014+)) | Shortness of breath on level ground |
| 4728 (imaging visit (2014+)) | Leg pain on walking |
| 4792 (imaging visit (2014+)) | Cochlear implant |
| 5663 (imaging visit (2014+)) | Longest manic/irritable episode |
| 6145 (response 3) (imaging visit (2014+)) | Stress: death of relative |
| 6145 (response 6) (imaging visit (2014+)) | Stress: financial |
| 6145 (response 2) (imaging visit (2014+)) | Stress: serious illness/assault to relative |
| 6145 (response 1) (imaging visit (2014+)) | Stress: serious illness/assault to self |
| 6145 (response 4) (imaging visit (2014+)) | Stress: death of spouse |
| 6145 (response 5) (imaging visit (2014+)) | Stress: divorce/separation |
| 6148 (response 5) (imaging visit (2014+)) | Eye problems/disorders: Macular degeneration |
| 6148 (response 4) (imaging visit (2014+)) | Eye problems/disorders: Cataract |
| 6148 (response 1) (imaging visit (2014+)) | Eye problems/disorders: Diabetes-related eye disease |
| 6148 (response 6) (imaging visit (2014+)) | Eye problems/disorders: Other serious eye condition |
| 6148 (response 2) (imaging visit (2014+)) | Eye problems/disorders: Glaucoma |
| 6148 (response 3) (imaging visit (2014+)) | Eye problems/disorders: Vision loss from injury |
| 6149 (response 1) (imaging visit (2014+)) | Mouth/teeth dental problems: Mouth ulcers |
| 6149 (response 3) (imaging visit (2014+)) | Mouth/teeth dental problems: Bleeding gums |
| 6149 (response 6) (imaging visit (2014+)) | Mouth/teeth dental problems: Dentures |
| 6149 (response 4) (imaging visit (2014+)) | Mouth/teeth dental problems: Loose teeth |
| 6149 (response 5) (imaging visit (2014+)) | Mouth/teeth dental problems: Toothache |
| 6149 (response 2) (imaging visit (2014+)) | Mouth/teeth dental problems: Painful gums |
| 6150 (imaging visit (2014+)) | Any vascular/heart problem |
| 6150 (response 4) (imaging visit (2014+)) | Vascular/heart problem: High blood pressure |
| 6150 (response 2) (imaging visit (2014+)) | Vascular/heart problem: Angina |
| 6150 (response 1) (imaging visit (2014+)) | Vascular/heart problem: Heart attack |
| 6150 (response 3) (imaging visit (2014+)) | Vascular/heart problem: Stroke |
| 20116 (imaging visit (2014+)) | Smoking status |
| 20117 (imaging visit (2014+)) | Alcohol drinker status |
| 20122 (initial assessment visit) | Bipolar disorder |
| 20123 (initial assessment visit) | Probable major depression (single episode) |
| 20419 (initial assessment visit) | Difficulty concentrating (anxiety) |
| 20425 (initial assessment visit) | Ever worried more than others |
| 20426 (initial assessment visit) | Restless (anxiety) |
| 20427 (initial assessment visit) | Sleep problems (anxiety) |
| 20437 (initial assessment visit) | Thoughts of death (depression) |
| 20449 (initial assessment visit) | Tiredness (depression) |
| 20450 (initial assessment visit) | Worthlessness (depression) |
| 20532 (initial assessment visit) | Sleep change |
| 20533 (initial assessment visit) | Trouble falling asleep |
| 20534 (initial assessment visit) | Sleeping too much |
| 20535 (initial assessment visit) | Waking too early |
| 20540 (initial assessment visit) | Multiple worries |
| 20541 (initial assessment visit) | Difficulty stopping worrying |
| 21024 (initial assessment visit) | IBS diagnosis |
| 21064 (initial assessment visit) | Sensitive stomach |
| 21065 (initial assessment visit) | Family history of IBS |
| ***Ordered Categorical*** |  |
| 884 (imaging visit (2014+)) | Moderate activity days/weeks |
| 894 (imaging visit (2014+)) | Moderate activity duration |
| 904 (imaging visit (2014+)) | Vigorous activity days/weeks |
| 924 (imaging visit (2014+)) | Walking pace |
| 991 (imaging visit (2014+)) | Strenuous sports frequency |
| 1001 (imaging visit (2014+)) | Strenuous sports duration |
| 1021 (imaging visit (2014+)) | Light DIY duration |
| 1200 (imaging visit (2014+)) | Sleeplessness/insomnia |
| 1220 (imaging visit (2014+)) | Daytime dozing |
| 1239 (imaging visit (2014+)) | Current smoking |
| 1558 (imaging visit (2014+)) | Alcohol intake frequency |
| 2050 (imaging visit (2014+)) | Depressed mood (last 2 weeks) |
| 2060 (imaging visit (2014+)) | Unenthusiasm (last 2 weeks) |
| 2070 (imaging visit (2014+)) | Tenseness/restlessness (last 2 weeks) |
| 2080 (imaging visit (2014+)) | Tiredness/lethargy (last 2 weeks) |
| 2110 (imaging visit (2014+)) | Able to confide |
| 2178 (imaging visit (2014+)) | Overall health rating |
| 2296 (imaging visit (2014+)) | Falls in last year |
| 4526 (imaging visit (2014+)) | Happiness |
| 4620 (imaging visit (2014+)) | Depression episodes |
| 6015 (initial assessment visit) | Chest pain during activity |
| 6016 (initial assessment visit) | Chest pain outside activity |
| 6373 (imaging visit (2014+)) | Number of puzzles correctly solved |
| 20016 (imaging visit (2014+)) | Fluid intelligence |
| 20127 (initial assessment visit) | Neuroticism |
| 20240 (imaging visit (2014+)) | Max digits remembered |
| 20458 (initial assessment visit) | General happiness |
| 20459 (initial assessment visit) | Happiness with health |
| 21048 (initial assessment visit) | Back pain for 3+ months |
| ***Unordered Categorical*** |  |
| 1538 (imaging visit (2014+)) | Major dietary changes (last 5 years) |
| 20126 (imaging visit (2014+)) | Bipolar and major depression status |
| 20536 (imaging visit (2014+)) | Weight change during worst depression episode |
| ***Continuous*** |  |
| 93 (imaging visit (2014+)) | Systolic BP (manual) |
| 94 (imaging visit (2014+)) | Diastolic BP (manual) |
| 95 (imaging visit (2014+)) | Pulse rate |
| 874 (imaging visit (2014+)) | Duration of walks |
| 914 (imaging visit (2014+)) | Duration of vigorous activity |
| 4079 (imaging visit (2014+)) | Diastolic BP (automated) |
| 4080 (imaging visit (2014+)) | Systolic BP (automated) |
| 4609 (imaging visit (2014+)) | Longest depression period |
| 5375 (imaging visit (2014+)) | Longest unenthusiasm/disinterest period |
| 20023 (imaging visit (2014+)) | Reaction time (mean) |
| 20420 (imaging visit (2014+)) | Longest worried/anxious period |
| 21001 (imaging visit (2014+)) | BMI |
| 21002 (imaging visit (2014+)) | Weight |

**eAppendix 2.** PHESANT Phenome Scan Tool.

The UK Biobank data showcase allows researchers to identify variables based on a field type (http://biobank.ctsu.ox.ac.uk/showcase/list.cgi). Field types include integer, continuous, categorical (single) and categorical (multiple) where fields pertain to a diverse range of phenotypes (e.g., blood samples, clinical assessments, health and lifestyle questionnaires). PHESANT processes and analyses each UKB data field separately. Its decision rule starts with the variable field type where a different rule is used to categorize each variable as one of four data types – continuous, ordered categorical, unordered categorical, or binary. Variables of the continuous and integer field type are assigned to the continuous data type. In cases where there are only a few distinct values, continuous variables are assigned as ordered categorical. Variables of the categorical (single) field type are assigned to either the binary, ordered categorical or unordered categorical type (depending on whether the field has two distinct values or has been specified as ordered or unordered in the PHESANT setup files) and variables of the categorical (multiple) field type are converted to a set of binary variables, one for each value in the categorical (multiple) fields. PHESANT applies an inverse normal rank transformation to outcomes of the continuous type prior to testing to ensure they are normally distributed.

**eAppendix 3.** Comparison between UKB imaging cohort with first brain MRI but no repeat imaging and current sample.

| **UK Biobank Variable** | **Imaging Cohort – first brain MRI** | **Current Sample** | ***P*-value** | **Comparison test** |
| --- | --- | --- | --- | --- |
| **Age at recruitment (years)** | 55.2 (7.6) | 52.6 (7.4) | <0.0001 | Welch t-test |
| **Sex, number (%)** | 50,372 | 4329 | <0.0001 | Chi-square test |
| Female  Male | 26,145 (51.9)  24,227 (48.1) | 2362 (54.6)  1967 (45.4) |  |  |
| **Systolic Blood Pressure (mmHg)** | 141.8 (20.1) | 138.3 (18.9) | <0.0001 | Welch t-test |
| **Diastolic Blood Pressure (mmHg)** | 78.9 (10.7) | 78.1 (10.6) | <0.0001 | Welch t-test |
| **Body Mass Index (Kg/m2)** | 26.6 (4.5) | 26.3 (4.3) | <0.0001 | Welch t-test |
| **Current Smoking Status, number (%)** | 50,001 | 4312 | <0.0001 | Chi-square test |
| No  Yes | 48,088 (96.5)  1732 (3.5) | 4165 (96.9)  133 (3.1) |  |  |
| **Brain Imaging, median (range)** | 43,355 | 4329 | <0.0001 | Welch t-test |
| Total volume of WMHs, first imaging visit | 2923 (11,4929) | 2473 (63,313) |  |  |

Descriptives are means (SD) unless otherwise specified. *Abbreviations:* NA, not applicable; MRI, magnetic resonance imaging.

**eAppendix 4.** Baseline WMH (% total brain tissue volume) across quintiles of raw WMH volume change.

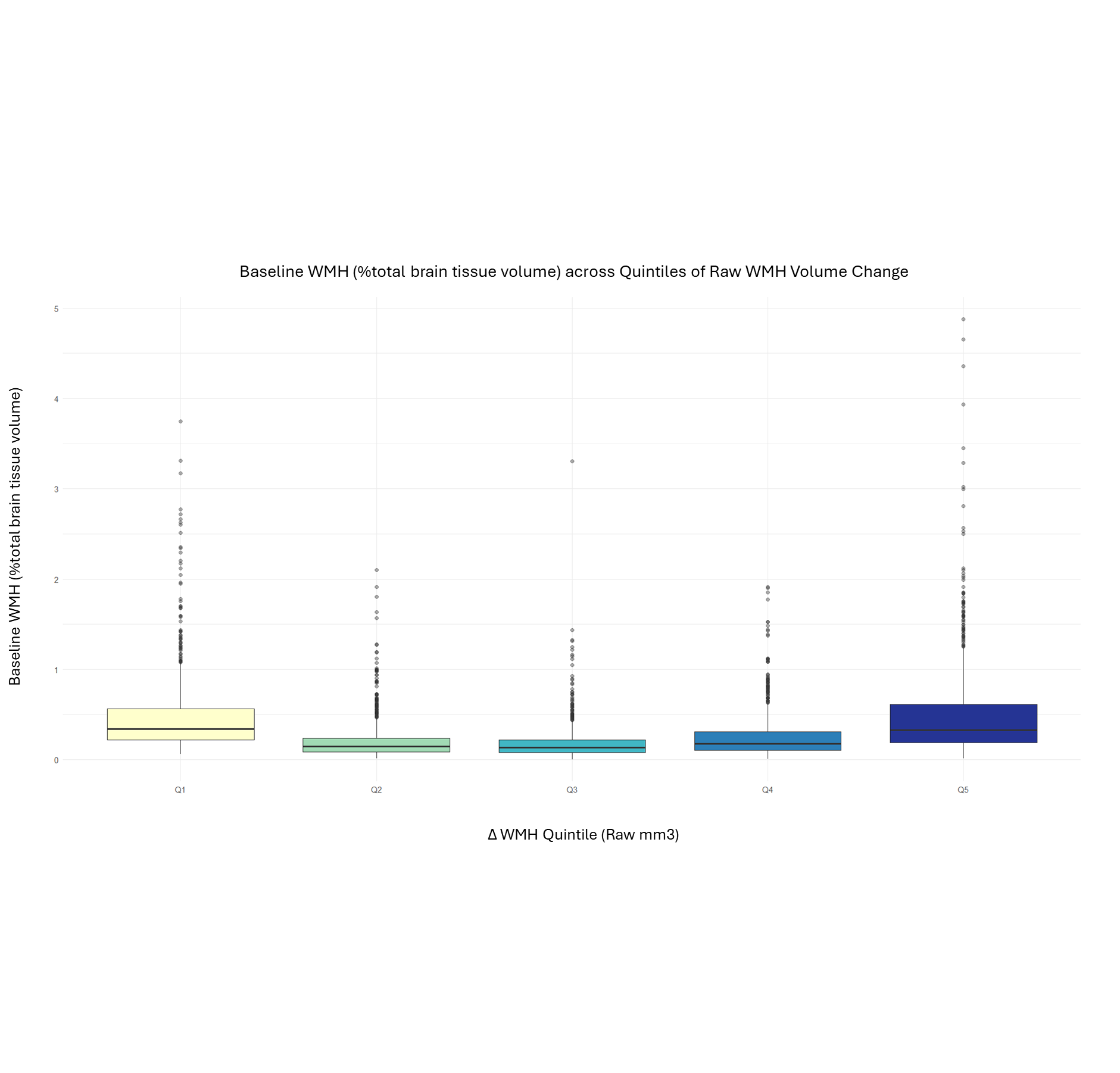

*Notes:* Boxplots depict baseline WMH volumes expressed as a percentage of intracranial volume (%total brain tissue volume) across quintiles of raw WMH change (ΔWMH_raw). Quintiles were derived from the sample distribution of WMH change (Q1 = most regression, Q5 = most progression), each containing roughly 20% of participants (N=4329 in total).

**eAppendix 5.** All linear regression results for baseline clinical phenotypes associated with subsequent WMH volume change.

| **UKB Phenotype** | **Predictor Type** | **Model** | **Estimate** | **SE** | **CI lower** | **CI upper** | ***P*** | **Adjusted *P*** |
| --- | --- | --- | --- | --- | --- | --- | --- | --- |
| Diastolic BP (automated) | Continuous | Partially adjusted | 0.0896 | 0.0182 | 0.0540 | 0.1253 | <0.0001 | 0.0001 |
| Cataract | Binary | Unadjusted | 0.2453 | 0.0539 | 0.1396 | 0.3510 | <0.0001 | 0.0003 |
| Systolic BP (automated) | Continuous | Unadjusted | 0.0832 | 0.0189 | 0.0462 | 0.1202 | <0.0001 | 0.0003 |
| Diastolic BP (automated) | Continuous | Unadjusted | 0.0729 | 0.0181 | 0.0374 | 0.1084 | 0.0001 | 0.0011 |
| Diastolic BP (automated) | Continuous | Fully adjusted | 0.0972 | 0.0243 | 0.0496 | 0.1449 | 0.0001 | 0.0030 |
| Dentures | Binary | Unadjusted | 0.1965 | 0.0547 | 0.0893 | 0.3036 | 0.0003 | 0.0037 |
| Reaction time (mean) | Continuous | Unadjusted | 0.0599 | 0.0165 | 0.0277 | 0.0922 | 0.0003 | 0.0037 |
| Hip pain 3+ months | Binary | Partially adjusted | -0.3854 | 0.1109 | -0.6034 | -0.1674 | 0.0006 | 0.0151 |
| Hip pain 3+ months | Binary | Unadjusted | -0.3536 | 0.1117 | -0.5733 | -0.1339 | 0.0017 | 0.0156 |
| Diabetes-related eye disease | Binary | Unadjusted | 0.5517 | 0.1895 | 0.1801 | 0.9234 | 0.0036 | 0.0290 |
| Unenthusiasm (2 weeks) | Ordered categorical | Unadjusted | -0.0909 | 0.0320 | -0.1536 | -0.0282 | 0.0045 | 0.0317 |
| Diabetes-related eye disease | Binary | Fully adjusted | 0.6448 | 0.2224 | 0.2089 | 1.0808 | 0.0038 | 0.0627 |
| Hip pain 3+ months | Binary | Fully adjusted | -0.3629 | 0.1250 | -0.6090 | -0.1168 | 0.0040 | 0.0627 |
| Systolic BP (automated) | Continuous | Partially adjusted | 0.0578 | 0.0199 | 0.0189 | 0.0968 | 0.0036 | 0.0645 |
| Hearing aid user | Binary | Unadjusted | 0.1686 | 0.0688 | 0.0337 | 0.3036 | 0.0143 | 0.0802 |
| Tiredness/lethargy (2 weeks) | Ordered categorical | Unadjusted | -0.0498 | 0.0202 | -0.0894 | -0.0101 | 0.0139 | 0.0802 |
| Neuroticism | Ordered categorical | Unadjusted | -0.0119 | 0.0052 | -0.0221 | -0.0016 | 0.0233 | 0.1188 |
| Miserableness | Binary | Unadjusted | -0.0702 | 0.0324 | -0.1337 | -0.0067 | 0.0303 | 0.1414 |
| Diabetes-related eye disease | Binary | Partially adjusted | 0.4725 | 0.1861 | 0.1077 | 0.8374 | 0.0111 | 0.1476 |
| Longest depression period | Continuous | Partially adjusted | 0.0591 | 0.0243 | 0.0114 | 0.1068 | 0.0152 | 0.1611 |
| Mood swings | Binary | Unadjusted | -0.0659 | 0.0322 | -0.1291 | -0.0027 | 0.0409 | 0.1613 |
| Fed-up feelings | Binary | Unadjusted | -0.0673 | 0.0333 | -0.1326 | -0.0021 | 0.0432 | 0.1613 |
| Longest depression period | Continuous | Unadjusted | 0.0499 | 0.0246 | 0.0017 | 0.0980 | 0.0424 | 0.1613 |
| Walking pace | Ordered categorical | Unadjusted | -0.0543 | 0.0275 | -0.1082 | -0.0004 | 0.0485 | 0.1699 |
| BMI | Continuous | Unadjusted | 0.0299 | 0.0160 | -0.0014 | 0.0612 | 0.0616 | 0.1938 |
| Bipolar and major depression status: Bipolar I | Unordered categorical | Unadjusted | 0.2466 | 0.1322 | -0.0127 | 0.5059 | 0.0623 | 0.1938 |
| Cataract | Binary | Partially adjusted | 0.1230 | 0.0548 | 0.0155 | 0.2304 | 0.0249 | 0.1976 |
| Longest unenthusiasm/disinterest period | Continuous | Partially adjusted | 0.0689 | 0.0309 | 0.0082 | 0.1295 | 0.0261 | 0.1976 |
| Depressed mood (2 weeks) | Ordered categorical | Unadjusted | -0.0567 | 0.0310 | -0.1176 | 0.0041 | 0.0676 | 0.1992 |
| Bipolar disorder | Binary | Partially adjusted | 1.8043 | 0.5673 | 0.2291 | 3.3795 | 0.0335 | 0.2000 |
| Painful gums | Binary | Partially adjusted | 0.2503 | 0.1180 | 0.0190 | 0.4816 | 0.0340 | 0.2000 |
| Chest pain outside activity | Ordered categorical | Fully adjusted | 0.3567 | 0.1527 | 0.0565 | 0.6569 | 0.0200 | 0.2350 |
| BMI | Continuous | Partially adjusted | 0.0306 | 0.0157 | -0.0003 | 0.0614 | 0.0524 | 0.2776 |
| Longest unenthusiasm/disinterest period | Continuous | Unadjusted | 0.0494 | 0.0313 | -0.0120 | 0.1108 | 0.1146 | 0.3143 |
| Tenseness/restlessness (2 weeks) | Ordered categorical | Unadjusted | -0.0470 | 0.0300 | -0.1059 | 0.0119 | 0.1179 | 0.3143 |
| Systolic BP (manual) | Continuous | Unadjusted | 0.0552 | 0.0370 | -0.0174 | 0.1277 | 0.1360 | 0.3322 |
| Weight | Continuous | Unadjusted | 0.0237 | 0.0161 | -0.0078 | 0.0552 | 0.1403 | 0.3322 |
| Vigorous activity days/weeks | Ordered categorical | Unadjusted | -0.0119 | 0.0081 | -0.0279 | 0.0040 | 0.1424 | 0.3322 |
| Able to confide | Ordered categorical | Unadjusted | -0.0121 | 0.0084 | -0.0287 | 0.0044 | 0.1513 | 0.3355 |
| Strenuous sports duration | Ordered categorical | Unadjusted | -0.0568 | 0.0400 | -0.1354 | 0.0218 | 0.1563 | 0.3355 |
| Moderate activity duration | Ordered categorical | Unadjusted | 0.0276 | 0.0199 | -0.0114 | 0.0666 | 0.1650 | 0.3355 |
| Weight change during worst depression episode: Both gained and lost | Unordered categorical | Unadjusted | -0.1557 | 0.1128 | -0.3769 | 0.0656 | 0.1677 | 0.3355 |
| Puzzle score | Ordered categorical | Unadjusted | -0.0110 | 0.0083 | -0.0272 | 0.0053 | 0.1854 | 0.3581 |
| Reaction time (mean) | Continuous | Partially adjusted | 0.0291 | 0.0170 | -0.0043 | 0.0625 | 0.0882 | 0.4249 |
| Pulse rate | Continuous | Unadjusted | 0.0397 | 0.0356 | -0.0301 | 0.1096 | 0.2645 | 0.4634 |
| Sleeplessness/insomnia | Ordered categorical | Unadjusted | -0.0231 | 0.0208 | -0.0638 | 0.0175 | 0.2648 | 0.4634 |
| Weight change during worst depression episode: Gained | Unordered categorical | Unadjusted | -0.0827 | 0.0740 | -0.2278 | 0.0624 | 0.2636 | 0.4634 |
| Strenuous sports duration | Ordered categorical | Partially adjusted | -0.0655 | 0.0403 | -0.1447 | 0.0137 | 0.1050 | 0.4638 |
| Daytime dozing | Ordered categorical | Unadjusted | 0.0353 | 0.0327 | -0.0289 | 0.0995 | 0.2811 | 0.4770 |
| Longest worried/anxious period | Continuous | Unadjusted | 0.0369 | 0.0371 | -0.0360 | 0.1098 | 0.3210 | 0.5287 |
| Fluid intelligence | Ordered categorical | Partially adjusted | -0.0118 | 0.0080 | -0.0276 | 0.0039 | 0.1408 | 0.5739 |
| General happiness | Ordered categorical | Unadjusted | -0.0207 | 0.0229 | -0.0657 | 0.0242 | 0.3655 | 0.5849 |
| Duration of vigorous activity | Continuous | Unadjusted | -0.0149 | 0.0188 | -0.0518 | 0.0220 | 0.4279 | 0.6624 |
| Back pain (3 months) | Ordered categorical | Unadjusted | -0.0189 | 0.0247 | -0.0672 | 0.0294 | 0.4437 | 0.6624 |
| Fluid intelligence | Ordered categorical | Unadjusted | -0.0061 | 0.0081 | -0.0218 | 0.0097 | 0.4522 | 0.6624 |
| Major dietary changes (5 years): because of other reasons | Unordered categorical | Unadjusted | -0.0248 | 0.0337 | -0.0908 | 0.0412 | 0.4613 | 0.6624 |
| Longest worried/anxious period | Continuous | Partially adjusted | 0.0478 | 0.0365 | -0.0239 | 0.1195 | 0.1908 | 0.6737 |
| Pulse rate | Continuous | Partially adjusted | 0.0382 | 0.0353 | -0.0310 | 0.1075 | 0.2791 | 0.6737 |
| Weight | Continuous | Partially adjusted | 0.0194 | 0.0183 | -0.0165 | 0.0554 | 0.2896 | 0.6737 |
| Vigorous activity days/weeks | Ordered categorical | Partially adjusted | -0.0099 | 0.0080 | -0.0256 | 0.0058 | 0.2173 | 0.6737 |
| Unenthusiasm (2 weeks) | Ordered categorical | Partially adjusted | -0.0387 | 0.0314 | -0.1003 | 0.0230 | 0.2190 | 0.6737 |
| Moderate activity days/weeks | Ordered categorical | Partially adjusted | -0.0085 | 0.0070 | -0.0221 | 0.0052 | 0.2238 | 0.6737 |
| Walking pace | Ordered categorical | Partially adjusted | -0.0313 | 0.0271 | -0.0844 | 0.0217 | 0.2472 | 0.6737 |
| Major dietary changes (5 years): because of illness | Unordered categorical | Partially adjusted | 0.0713 | 0.0677 | -0.0614 | 0.2039 | 0.2924 | 0.6737 |
| Bipolar and major depression status: Bipolar I | Unordered categorical | Partially adjusted | 0.1482 | 0.1391 | -0.1249 | 0.4212 | 0.2872 | 0.6737 |
| Weight change during worst depression episode: Both gained and lost | Unordered categorical | Partially adjusted | -0.1323 | 0.1109 | -0.3498 | 0.0853 | 0.2331 | 0.6737 |
| Reaction time (mean) | Continuous | Fully adjusted | 0.0328 | 0.0200 | -0.0064 | 0.0721 | 0.1013 | 0.6741 |
| Longest depression period | Continuous | Fully adjusted | 0.0451 | 0.0286 | -0.0110 | 0.1013 | 0.1151 | 0.6741 |
| Longest unenthusiasm/disinterest period | Continuous | Fully adjusted | 0.0551 | 0.0365 | -0.0166 | 0.1267 | 0.1318 | 0.6741 |
| Unenthusiasm (2 weeks) | Ordered categorical | Fully adjusted | -0.0652 | 0.0364 | -0.1365 | 0.0061 | 0.0731 | 0.6741 |
| Moderate activity days/weeks | Ordered categorical | Fully adjusted | -0.0132 | 0.0082 | -0.0292 | 0.0029 | 0.1070 | 0.6741 |
| Major dietary changes (5 years): because of illness | Unordered categorical | Fully adjusted | 0.1132 | 0.0773 | -0.0384 | 0.2648 | 0.1434 | 0.6741 |
| Alcohol intake frequency | Ordered categorical | Fully adjusted | 0.0182 | 0.0130 | -0.0074 | 0.0437 | 0.1629 | 0.6962 |
| BMI | Continuous | Fully adjusted | 0.0251 | 0.0192 | -0.0125 | 0.0627 | 0.1905 | 0.7076 |
| Longest worried/anxious period | Continuous | Fully adjusted | 0.0509 | 0.0420 | -0.0316 | 0.1334 | 0.2258 | 0.7076 |
| Fluid intelligence | Ordered categorical | Fully adjusted | -0.0121 | 0.0095 | -0.0307 | 0.0066 | 0.2048 | 0.7076 |
| Sleeplessness/insomnia | Ordered categorical | Fully adjusted | -0.0304 | 0.0243 | -0.0780 | 0.0172 | 0.2108 | 0.7076 |
| Chest pain outside activity | Ordered categorical | Partially adjusted | 0.1194 | 0.1227 | -0.1216 | 0.3604 | 0.3310 | 0.7310 |
| Duration of walks | Continuous | Unadjusted | -0.0088 | 0.0154 | -0.0390 | 0.0213 | 0.5662 | 0.7404 |
| Moderate activity days/weeks | Ordered categorical | Unadjusted | -0.0044 | 0.0071 | -0.0183 | 0.0094 | 0.5309 | 0.7404 |
| Max digits remembered | Ordered categorical | Unadjusted | -0.0085 | 0.0148 | -0.0375 | 0.0206 | 0.5685 | 0.7404 |
| Major dietary changes (5 years): because of illness | Unordered categorical | Unadjusted | 0.0409 | 0.0694 | -0.0951 | 0.1770 | 0.5554 | 0.7404 |
| Strenuous sports frequency | Ordered categorical | Unadjusted | 0.0205 | 0.0378 | -0.0538 | 0.0949 | 0.5874 | 0.7476 |
| Strenuous sports frequency | Ordered categorical | Fully adjusted | 0.0487 | 0.0452 | -0.0401 | 0.1375 | 0.2819 | 0.7967 |
| Neuroticism | Ordered categorical | Fully adjusted | -0.0065 | 0.0061 | -0.0186 | 0.0055 | 0.2882 | 0.7967 |
| Chest pain during activity | Ordered categorical | Unadjusted | -0.0753 | 0.1646 | -0.3986 | 0.2479 | 0.6474 | 0.7999 |
| Falls in last year | Ordered categorical | Unadjusted | -0.0137 | 0.0308 | -0.0741 | 0.0467 | 0.6571 | 0.7999 |
| Happiness with health | Ordered categorical | Unadjusted | -0.0076 | 0.0195 | -0.0459 | 0.0307 | 0.6975 | 0.8251 |
| Depression episodes | Ordered categorical | Unadjusted | 0.0106 | 0.0282 | -0.0447 | 0.0659 | 0.7072 | 0.8251 |
| Alcohol intake frequency | Ordered categorical | Unadjusted | -0.0039 | 0.0112 | -0.0259 | 0.0180 | 0.7250 | 0.8285 |
| Chest pain outside activity | Ordered categorical | Unadjusted | 0.0425 | 0.1298 | -0.2124 | 0.2974 | 0.7433 | 0.8325 |
| Sleeplessness/insomnia | Ordered categorical | Partially adjusted | -0.0174 | 0.0206 | -0.0578 | 0.0230 | 0.3996 | 0.8333 |
| Alcohol intake frequency | Ordered categorical | Partially adjusted | 0.0091 | 0.0111 | -0.0126 | 0.0309 | 0.4088 | 0.8333 |
| Light DIY duration | Ordered categorical | Fully adjusted | -0.0152 | 0.0159 | -0.0464 | 0.0160 | 0.3390 | 0.8437 |
| Chest pain during activity | Ordered categorical | Fully adjusted | -0.1789 | 0.1877 | -0.5478 | 0.1900 | 0.3411 | 0.8437 |
| Able to confide | Ordered categorical | Partially adjusted | -0.0064 | 0.0083 | -0.0227 | 0.0100 | 0.4447 | 0.8548 |
| Neuroticism | Ordered categorical | Partially adjusted | -0.0039 | 0.0052 | -0.0141 | 0.0063 | 0.4516 | 0.8548 |
| General happiness | Ordered categorical | Fully adjusted | -0.0226 | 0.0259 | -0.0733 | 0.0281 | 0.3815 | 0.8571 |
| Strenuous sports duration | Ordered categorical | Fully adjusted | -0.0426 | 0.0488 | -0.1385 | 0.0533 | 0.3830 | 0.8571 |
| Duration of vigorous activity | Continuous | Fully adjusted | 0.0169 | 0.0215 | -0.0252 | 0.0591 | 0.4306 | 0.8749 |
| Weight | Continuous | Fully adjusted | 0.0170 | 0.0224 | -0.0269 | 0.0610 | 0.4468 | 0.8749 |
| Bipolar and major depression status: Bipolar II | Unordered categorical | Fully adjusted | -0.0662 | 0.0812 | -0.2256 | 0.0932 | 0.4154 | 0.8749 |
| Chest pain during activity | Ordered categorical | Partially adjusted | -0.1103 | 0.1557 | -0.4162 | 0.1955 | 0.4789 | 0.8753 |
| Strenuous sports frequency | Ordered categorical | Partially adjusted | 0.0253 | 0.0377 | -0.0486 | 0.0993 | 0.5013 | 0.8856 |
| Diastolic BP (manual) | Continuous | Unadjusted | 0.0083 | 0.0356 | -0.0616 | 0.0782 | 0.8151 | 0.8950 |
| Walking pace | Ordered categorical | Fully adjusted | -0.0201 | 0.0321 | -0.0830 | 0.0429 | 0.5318 | 0.9113 |
| Falls in last year | Ordered categorical | Fully adjusted | 0.0211 | 0.0361 | -0.0498 | 0.0919 | 0.5598 | 0.9113 |
| Depression episodes | Ordered categorical | Fully adjusted | -0.0187 | 0.0326 | -0.0826 | 0.0453 | 0.5674 | 0.9113 |
| Moderate activity duration | Ordered categorical | Fully adjusted | 0.0131 | 0.0232 | -0.0324 | 0.0586 | 0.5733 | 0.9113 |
| Back pain (3 months) | Ordered categorical | Fully adjusted | -0.0157 | 0.0284 | -0.0714 | 0.0401 | 0.5817 | 0.9113 |
| Weight change during worst depression episode: Both gained and lost | Unordered categorical | Fully adjusted | -0.0819 | 0.1312 | -0.3393 | 0.1756 | 0.5326 | 0.9113 |
| Overall health rating | Ordered categorical | Unadjusted | -0.0044 | 0.0239 | -0.0512 | 0.0424 | 0.8537 | 0.9193 |
| Weight change during worst depression episode: Lost | Unordered categorical | Unadjusted | -0.0066 | 0.0569 | -0.1183 | 0.1050 | 0.9072 | 0.9586 |
| Systolic BP (automated) | Continuous | Fully adjusted | -0.0125 | 0.0265 | -0.0645 | 0.0394 | 0.6362 | 0.9596 |
| Daytime dozing | Ordered categorical | Fully adjusted | 0.0169 | 0.0376 | -0.0568 | 0.0905 | 0.6533 | 0.9596 |
| Tiredness/lethargy (2 weeks) | Ordered categorical | Partially adjusted | -0.0110 | 0.0200 | -0.0503 | 0.0283 | 0.5823 | 0.9603 |
| Overall health rating | Ordered categorical | Partially adjusted | 0.0129 | 0.0234 | -0.0330 | 0.0587 | 0.5825 | 0.9603 |
| Moderate activity duration | Ordered categorical | Partially adjusted | 0.0104 | 0.0196 | -0.0281 | 0.0488 | 0.5979 | 0.9603 |
| Bipolar and major depression status: Bipolar II | Unordered categorical | Unadjusted | 0.0060 | 0.0699 | -0.1312 | 0.1432 | 0.9314 | 0.9659 |
| Daytime dozing | Ordered categorical | Partially adjusted | 0.0159 | 0.0321 | -0.0470 | 0.0788 | 0.6207 | 0.9675 |
| Systolic BP (manual) | Continuous | Partially adjusted | 0.0112 | 0.0384 | -0.0643 | 0.0866 | 0.7714 | 0.9687 |
| Diastolic BP (manual) | Continuous | Partially adjusted | 0.0057 | 0.0359 | -0.0647 | 0.0761 | 0.8729 | 0.9687 |
| Duration of vigorous activity | Continuous | Partially adjusted | -0.0029 | 0.0185 | -0.0391 | 0.0333 | 0.8762 | 0.9687 |
| Happiness with health | Ordered categorical | Partially adjusted | 0.0064 | 0.0191 | -0.0311 | 0.0439 | 0.7381 | 0.9687 |
| Light DIY duration | Ordered categorical | Partially adjusted | -0.0037 | 0.0137 | -0.0305 | 0.0231 | 0.7857 | 0.9687 |
| Current smoking | Ordered categorical | Partially adjusted | 0.0128 | 0.0529 | -0.0909 | 0.1165 | 0.8088 | 0.9687 |
| Falls in last year | Ordered categorical | Partially adjusted | -0.0068 | 0.0303 | -0.0661 | 0.0526 | 0.8234 | 0.9687 |
| Depression episodes | Ordered categorical | Partially adjusted | 0.0061 | 0.0278 | -0.0484 | 0.0607 | 0.8257 | 0.9687 |
| General happiness | Ordered categorical | Partially adjusted | 0.0039 | 0.0225 | -0.0403 | 0.0481 | 0.8619 | 0.9687 |
| Tenseness/restlessness (2 weeks) | Ordered categorical | Partially adjusted | -0.0039 | 0.0294 | -0.0616 | 0.0538 | 0.8958 | 0.9687 |
| Max digits remembered | Ordered categorical | Partially adjusted | 0.0017 | 0.0147 | -0.0272 | 0.0305 | 0.9106 | 0.9687 |
| Puzzle score | Ordered categorical | Partially adjusted | 0.0009 | 0.0085 | -0.0157 | 0.0176 | 0.9139 | 0.9687 |
| Major dietary changes (5 years): because of other reasons | Unordered categorical | Partially adjusted | 0.0142 | 0.0330 | -0.0505 | 0.0790 | 0.6666 | 0.9687 |
| Bipolar and major depression status: Bipolar II | Unordered categorical | Partially adjusted | 0.0086 | 0.0678 | -0.1244 | 0.1416 | 0.8991 | 0.9687 |
| Weight change during worst depression episode: Gained | Unordered categorical | Partially adjusted | -0.0136 | 0.0741 | -0.1590 | 0.1317 | 0.8540 | 0.9687 |
| Weight change during worst depression episode: Lost | Unordered categorical | Partially adjusted | 0.0097 | 0.0572 | -0.1026 | 0.1220 | 0.8653 | 0.9687 |
| Overall health rating | Ordered categorical | Fully adjusted | 0.0113 | 0.0277 | -0.0430 | 0.0655 | 0.6842 | 0.9745 |
| Duration of walks | Continuous | Fully adjusted | -0.0022 | 0.0178 | -0.0371 | 0.0327 | 0.9006 | 0.9810 |
| Depressed mood (2 weeks) | Ordered categorical | Fully adjusted | -0.0104 | 0.0359 | -0.0808 | 0.0601 | 0.7730 | 0.9810 |
| Tiredness/lethargy (2 weeks) | Ordered categorical | Fully adjusted | -0.0060 | 0.0233 | -0.0516 | 0.0396 | 0.7960 | 0.9810 |
| Current smoking | Ordered categorical | Fully adjusted | -0.0077 | 0.0628 | -0.1308 | 0.1154 | 0.9024 | 0.9810 |
| Tenseness/restlessness (2 weeks) | Ordered categorical | Fully adjusted | -0.0039 | 0.0341 | -0.0707 | 0.0629 | 0.9090 | 0.9810 |
| Happiness with health | Ordered categorical | Fully adjusted | -0.0025 | 0.0225 | -0.0466 | 0.0417 | 0.9121 | 0.9810 |
| Puzzle score | Ordered categorical | Fully adjusted | 0.0010 | 0.0101 | -0.0189 | 0.0209 | 0.9226 | 0.9810 |
| Vigorous activity days/weeks | Ordered categorical | Fully adjusted | 0.0007 | 0.0093 | -0.0175 | 0.0189 | 0.9399 | 0.9810 |
| Max digits remembered | Ordered categorical | Fully adjusted | -0.0013 | 0.0171 | -0.0349 | 0.0324 | 0.9416 | 0.9810 |
| Major dietary changes (5 years): because of other reasons | Unordered categorical | Fully adjusted | 0.0061 | 0.0389 | -0.0701 | 0.0823 | 0.8751 | 0.9810 |
| Bipolar and major depression status: Bipolar I | Unordered categorical | Fully adjusted | 0.0079 | 0.1589 | -0.3040 | 0.3199 | 0.9602 | 0.9810 |
| Weight change during worst depression episode: Gained | Unordered categorical | Fully adjusted | 0.0247 | 0.0888 | -0.1496 | 0.1990 | 0.7812 | 0.9810 |
| Weight change during worst depression episode: Lost | Unordered categorical | Fully adjusted | 0.0114 | 0.0695 | -0.1249 | 0.1477 | 0.8694 | 0.9810 |
| Light DIY duration | Ordered categorical | Unadjusted | -0.0005 | 0.0139 | -0.0278 | 0.0267 | 0.9690 | 0.9866 |
| Able to confide | Ordered categorical | Fully adjusted | -0.0001 | 0.0098 | -0.0194 | 0.0192 | 0.9892 | 0.9892 |
| Current smoking | Ordered categorical | Unadjusted | -0.0007 | 0.0537 | -0.1059 | 0.1046 | 0.9901 | 0.9901 |
| Duration of walks | Continuous | Partially adjusted | -0.0008 | 0.0150 | -0.0303 | 0.0287 | 0.9567 | 0.9942 |
| Depressed mood (2 weeks) | Ordered categorical | Partially adjusted | -0.0009 | 0.0307 | -0.0610 | 0.0593 | 0.9775 | 0.9957 |
| Back pain (3 months) | Ordered categorical | Partially adjusted | -0.0001 | 0.0241 | -0.0474 | 0.0471 | 0.9957 | 0.9957 |

*Abbreviations:* BP, blood pressure; CI, confidence intervals; FDR, False Discovery Rate-corrected; SE, standard error; UKB, UK Biobank. For binary predictors, estimates represent the standardized difference in WMH change between individuals with a condition and those without. For ordered categorical predictors, estimates reflect the standardized difference in WMH change per one-step increase in category. For unordered categorical predictors, estimates reflect the standardized difference in WMH change compared to the reference category. For continuous predictors, estimates represents the standardized difference in WMH volume change per 1 SD increase in the predictor (inverse rank normal transformed by PHESANT as required). Three levels of adjustment include: (1) unadjusted (predictor only), (2) minimally adjusted for age, sex, total brain tissue volume, and follow-up duration, and (3) fully adjusted, additionally including blood pressure.

**eAppendix 6.** A comparison between WMH progressors, WMH regressors, and WMH stable (N=4329).

| **UK Biobank Variable** | **WMH Progressors (n=2337)** | **WMH Regressors (n=1126)** | **WMH Stable (n=866)** |
| --- | --- | --- | --- |
| **Age at recruitment (years), median (IQR)** | 53 (13) | 53 (11) | 49 (10) |
| **Sex, number (%)** | | |  |
| Female  Male | 1264 (54.1)  1073 (45.9) | 568 (50.4)  558 (49.6) | 530 (61.2)  336 (38.8) |
| **Smoking, number (%)** | | | |
| Yes, on most or all days  Only occasionally  No  Prefer not to answer | 44 (1.9)  38 (1.6)  2243 (96)  12 (0.5) | 18 (1.6)  15 (1.3)  1088 (96.6)  5 (0.4) | 12 (1.4)  9 (1.04)  844 (97.5)  1 (0.1) |
| **Systolic and Diastolic Blood Pressure, number** | 1730 | 777 | 622 |
| **Systolic Blood Pressure (automated; mmHg), mean (SD)** | 139.3 (18.8) | 139.05 (19.3) | 134.7 (18.07) |
| **Diastolic Blood Pressure (automated; mmHg), mean (SD)** | 78.4 (10.7) | 78.08 (10.5) | 77.1 (10.6) |
| **Body Mass Index, number** | 2235 | 1065 | 820 |
| **Body Mass Index (Kg/m2), mean (SD)** | 26.3 (4.4) | 26.4 (4.2) | 25.9 (4.3) |
| **Total Volume of WMHs (mm3)** | | |  |
| First imaging visit, median (IQR) | 2474 (3291) | 3566.5 (3928.8) | 1582.5 (1592.8) |
| Follow-up imaging visit, median (IQR) | 3998 (4949) | 2213 (2747.8) | 1538.5 (1597) |
| Raw difference between time points (first subtracted from follow-up) | 1524 (net increase) | -1353.5 (net decrease) | -44 (net decrease) |

*Abbreviations:* IQR, interquartile range; SD, standard deviation; WMH, white matter hyperintensities.

**eAppendix 7.** All unadjusted multinomial regression results for baseline clinical phenotypes associated with subsequent WMH volume change not surviving FDR-correction at *p*<0.05.

| **UKB Phenotype** | **Comparison (versus Stable)** | **Predictor Type** | **Estimate** | **SE** | **CI lower** | **CI upper** | **OR** | **OR lower** | **OR upper** | ***P*** | **Adjusted *P*** |
| --- | --- | --- | --- | --- | --- | --- | --- | --- | --- | --- | --- |
| Cataract | Regression | binary | 0.3566 | 0.1844 | -0.0048 | 0.7179 | 1.4284 | 0.9952 | 2.0502 | 0.0531 | 0.3988 |
| Bleeding gums | Regression | binary | -0.2736 | 0.1415 | -0.5510 | 0.0038 | 0.7606 | 0.5764 | 1.0038 | 0.0532 | 0.3988 |
| Stress: financial | Regression | binary | -0.5038 | 0.2618 | -1.0170 | 0.0095 | 0.6043 | 0.3617 | 1.0095 | 0.0544 | 0.3988 |
| Hip pain 3+ months | Regression | binary | -0.6852 | 0.3565 | -1.3841 | 0.0136 | 0.5040 | 0.2506 | 1.0137 | 0.0546 | 0.3988 |
| Tiredness (depression) | Regression | binary | -0.3754 | 0.1958 | -0.7593 | 0.0084 | 0.6870 | 0.4680 | 1.0085 | 0.0552 | 0.3988 |
| Painful gums | Regression | binary | -0.7051 | 0.3896 | -1.4688 | 0.0585 | 0.4941 | 0.2302 | 1.0603 | 0.0703 | 0.4779 |
| Abdominal pain 3+ months | Progression | binary | -0.6302 | 0.3812 | -1.3774 | 0.1170 | 0.5325 | 0.2522 | 1.1241 | 0.0983 | 0.5552 |
| Mood swings | Progression | binary | -0.1346 | 0.0833 | -0.2978 | 0.0287 | 0.8741 | 0.7424 | 1.0291 | 0.1063 | 0.5552 |
| Worthlessness (depression) | Regression | binary | -0.2311 | 0.1432 | -0.5117 | 0.0495 | 0.7937 | 0.5995 | 1.0507 | 0.1065 | 0.5552 |
| Worthlessness (depression) | Progression | binary | -0.2036 | 0.1265 | -0.4515 | 0.0443 | 0.8158 | 0.6367 | 1.0453 | 0.1074 | 0.5552 |
| Wears glasses/contacts | Progression | binary | 0.2454 | 0.1569 | -0.0622 | 0.5530 | 1.2781 | 0.9397 | 1.7385 | 0.1179 | 0.5617 |
| Guilty feelings | Regression | binary | -0.1612 | 0.1035 | -0.3640 | 0.0416 | 0.8511 | 0.6949 | 1.0424 | 0.1192 | 0.5617 |
| Wheeze in chest | Regression | binary | -0.2065 | 0.1342 | -0.4695 | 0.0565 | 0.8134 | 0.6253 | 1.0581 | 0.1238 | 0.5719 |
| Multiple worries | Progression | binary | -0.3038 | 0.2000 | -0.6957 | 0.0882 | 0.7380 | 0.4987 | 1.0922 | 0.1287 | 0.5825 |
| Suffer from nerves | Progression | binary | -0.1652 | 0.1102 | -0.3812 | 0.0507 | 0.8477 | 0.6831 | 1.0521 | 0.1338 | 0.5830 |
| Sleeping too much | Regression | binary | -0.2898 | 0.1977 | -0.6773 | 0.0977 | 0.7484 | 0.5080 | 1.1026 | 0.1427 | 0.6104 |
| Multiple worries | Regression | binary | -0.3246 | 0.2264 | -0.7683 | 0.1190 | 0.7228 | 0.4638 | 1.1264 | 0.1515 | 0.6117 |
| Sleep problems (anxiety) | Progression | binary | 0.2998 | 0.2094 | -0.1107 | 0.7103 | 1.3496 | 0.8952 | 2.0346 | 0.1523 | 0.6117 |
| Smoking status | Regression | binary | -0.3676 | 0.2605 | -0.8782 | 0.1429 | 0.6924 | 0.4155 | 1.1536 | 0.1582 | 0.6117 |
| Other serious eye condition | Progression | binary | -0.4300 | 0.3060 | -1.0298 | 0.1698 | 0.6505 | 0.3571 | 1.1851 | 0.1600 | 0.6117 |
| Thoughts of death (depression) | Regression | binary | -0.1998 | 0.1429 | -0.4799 | 0.0803 | 0.8189 | 0.6188 | 1.0836 | 0.1620 | 0.6117 |
| Miserableness | Progression | binary | -0.1159 | 0.0837 | -0.2799 | 0.0481 | 0.8906 | 0.7559 | 1.0493 | 0.1662 | 0.6117 |
| Long-standing illness/disability | Regression | binary | 0.1496 | 0.1083 | -0.0626 | 0.3619 | 1.1614 | 0.9393 | 1.4360 | 0.1670 | 0.6117 |
| Stress: serious illness/assault self | Progression | binary | 0.2697 | 0.1989 | -0.1201 | 0.6595 | 1.3096 | 0.8869 | 1.9338 | 0.1750 | 0.6220 |
| Tiredness (depression) | Progression | binary | -0.2366 | 0.1772 | -0.5839 | 0.1107 | 0.7893 | 0.5577 | 1.1170 | 0.1817 | 0.6336 |
| Dentures | Regression | binary | 0.2296 | 0.1738 | -0.1111 | 0.5703 | 1.2581 | 0.8949 | 1.7687 | 0.1865 | 0.6336 |
| Sleeping too much | Progression | binary | -0.2171 | 0.1717 | -0.5537 | 0.1195 | 0.8048 | 0.5748 | 1.1269 | 0.2062 | 0.6803 |
| Diabetes-related eye disease | Progression | binary | 0.6740 | 0.5470 | -0.3981 | 1.7462 | 1.9621 | 0.6716 | 5.7325 | 0.2179 | 0.6982 |
| Loneliness/isolation | Progression | binary | -0.1425 | 0.1163 | -0.3705 | 0.0855 | 0.8672 | 0.6904 | 1.0893 | 0.2206 | 0.6982 |
| Headaches 3+ months | Regression | binary | 0.3112 | 0.2570 | -0.1925 | 0.8148 | 1.3650 | 0.8249 | 2.2587 | 0.2259 | 0.6983 |
| Fed-up feelings | Progression | binary | -0.1028 | 0.0865 | -0.2724 | 0.0668 | 0.9023 | 0.7615 | 1.0690 | 0.2346 | 0.7132 |
| Shortness of breath on level ground | Progression | binary | -0.1964 | 0.1690 | -0.5276 | 0.1348 | 0.8217 | 0.5900 | 1.1443 | 0.2451 | 0.7345 |
| Abdominal pain 3+ months | Regression | binary | -0.4504 | 0.4137 | -1.2613 | 0.3604 | 0.6374 | 0.2833 | 1.4340 | 0.2763 | 0.7883 |
| Chest pain when walking | Regression | binary | 0.5695 | 0.5242 | -0.4579 | 1.5970 | 1.7674 | 0.6326 | 4.9380 | 0.2773 | 0.7883 |
| Diabetes diagnosed | Regression | binary | 0.2484 | 0.2324 | -0.2071 | 0.7039 | 1.2820 | 0.8130 | 2.0215 | 0.2851 | 0.7883 |
| Neck/shoulder pain 3+ months | Regression | binary | -0.2651 | 0.2488 | -0.7527 | 0.2225 | 0.7672 | 0.4711 | 1.2492 | 0.2867 | 0.7883 |
| Diabetes diagnosed | Progression | binary | 0.2210 | 0.2088 | -0.1884 | 0.6303 | 1.2473 | 0.8283 | 1.8782 | 0.2901 | 0.7883 |
| Facial pain 3+ months: No | Progression | binary | 0.8473 | 0.8112 | -0.7426 | 2.4372 | 2.3333 | 0.4759 | 11.4410 | 0.2962 | 0.7957 |
| Vision loss from injury | Progression | binary | 1.0928 | 1.0613 | -0.9874 | 3.1729 | 2.9825 | 0.3726 | 23.8767 | 0.3032 | 0.8050 |
| Difficulty stopping worrying | Regression | binary | -0.4118 | 0.4235 | -1.2419 | 0.4183 | 0.6625 | 0.2888 | 1.5193 | 0.3309 | 0.8315 |
| Neck/shoulder pain 3+ months | Progression | binary | -0.2100 | 0.2165 | -0.6344 | 0.2143 | 0.8106 | 0.5303 | 1.2390 | 0.3320 | 0.8315 |
| Snoring | Progression | binary | -0.0833 | 0.0865 | -0.2528 | 0.0862 | 0.9201 | 0.7767 | 1.0900 | 0.3356 | 0.8315 |
| Miserableness | Regression | binary | -0.0917 | 0.0953 | -0.2784 | 0.0950 | 0.9124 | 0.7570 | 1.0997 | 0.3358 | 0.8315 |
| High blood pressure | Progression | binary | -0.3947 | 0.4122 | -1.2027 | 0.4133 | 0.6739 | 0.3004 | 1.5118 | 0.3384 | 0.8315 |
| Restless (anxiety) | Regression | binary | -0.1887 | 0.2023 | -0.5851 | 0.2078 | 0.8281 | 0.5570 | 1.2309 | 0.3509 | 0.8331 |
| Ever worried more than others | Progression | binary | -0.1046 | 0.1123 | -0.3247 | 0.1156 | 0.9007 | 0.7227 | 1.1225 | 0.3517 | 0.8331 |
| Shortness of breath on level ground | Regression | binary | -0.1801 | 0.1941 | -0.5604 | 0.2003 | 0.8352 | 0.5710 | 1.2217 | 0.3534 | 0.8331 |
| Longest manic/irritable episode | Regression | binary | 0.1429 | 0.1628 | -0.1763 | 0.4620 | 1.1536 | 0.8384 | 1.5872 | 0.3802 | 0.8595 |
| Guilty feelings | Progression | binary | -0.0782 | 0.0899 | -0.2544 | 0.0979 | 0.9247 | 0.7754 | 1.1029 | 0.3840 | 0.8595 |
| Ever worried more than others | Regression | binary | -0.1092 | 0.1285 | -0.3610 | 0.1426 | 0.8965 | 0.6970 | 1.1533 | 0.3952 | 0.8595 |
| Smoking status | Progression | binary | -0.1813 | 0.2163 | -0.6052 | 0.2427 | 0.8342 | 0.5460 | 1.2747 | 0.4021 | 0.8595 |
| Sleep problems (anxiety) | Regression | binary | 0.2013 | 0.2411 | -0.2712 | 0.6739 | 1.2230 | 0.7624 | 1.9618 | 0.4037 | 0.8595 |
| Ever highly irritable | Regression | binary | -0.1118 | 0.1355 | -0.3774 | 0.1539 | 0.8943 | 0.6856 | 1.1663 | 0.4096 | 0.8595 |
| Worry too long after embarrassment | Progression | binary | -0.0665 | 0.0812 | -0.2257 | 0.0927 | 0.9357 | 0.7979 | 1.0972 | 0.4130 | 0.8595 |
| Probable major depression (single episode) | Progression | binary | -0.2032 | 0.2543 | -0.7017 | 0.2953 | 0.8161 | 0.4958 | 1.3436 | 0.4244 | 0.8676 |
| Risk taking | Regression | binary | -0.0834 | 0.1044 | -0.2880 | 0.1212 | 0.9200 | 0.7497 | 1.1289 | 0.4244 | 0.8676 |
| Other serious eye condition | Regression | binary | -0.2615 | 0.3460 | -0.9398 | 0.4167 | 0.7699 | 0.3907 | 1.5170 | 0.4498 | 0.9031 |
| Fracture from fall | Progression | binary | -0.2084 | 0.2780 | -0.7532 | 0.3365 | 0.8119 | 0.4708 | 1.4000 | 0.4535 | 0.9031 |
| Diabetes-related eye disease | Regression | binary | -0.5426 | 0.7651 | -2.0422 | 0.9570 | 0.5812 | 0.1297 | 2.6039 | 0.4782 | 0.9274 |
| Sensitive stomach | Progression | binary | 0.0805 | 0.1139 | -0.1428 | 0.3038 | 1.0838 | 0.8669 | 1.3550 | 0.4799 | 0.9274 |
| Irritability | Regression | binary | 0.0740 | 0.1049 | -0.1316 | 0.2795 | 1.0768 | 0.8767 | 1.3225 | 0.4807 | 0.9274 |
| Irritability | Progression | binary | -0.0651 | 0.0934 | -0.2481 | 0.1180 | 0.9370 | 0.7802 | 1.1252 | 0.4859 | 0.9274 |
| Waking too early | Regression | binary | 0.1293 | 0.1946 | -0.2521 | 0.5107 | 1.1380 | 0.7771 | 1.6665 | 0.5064 | 0.9274 |
| Suffer from nerves | Regression | binary | -0.0823 | 0.1249 | -0.3272 | 0.1626 | 0.9210 | 0.7209 | 1.1765 | 0.5100 | 0.9274 |
| Loose teeth | Progression | binary | 0.2176 | 0.3316 | -0.4323 | 0.8675 | 1.2431 | 0.6490 | 2.3809 | 0.5117 | 0.9274 |
| Ever disinterested â‰¥1 week | Progression | binary | -0.0549 | 0.0852 | -0.2219 | 0.1120 | 0.9465 | 0.8010 | 1.1185 | 0.5190 | 0.9274 |
| Trouble falling asleep | Progression | binary | -0.1062 | 0.1660 | -0.4315 | 0.2191 | 0.8992 | 0.6495 | 1.2449 | 0.5222 | 0.9274 |
| Wears glasses/contacts | Regression | binary | 0.1098 | 0.1765 | -0.2362 | 0.4558 | 1.1161 | 0.7896 | 1.5775 | 0.5339 | 0.9274 |
| Stress: divorce/separation | Regression | binary | -0.2607 | 0.4288 | -1.1012 | 0.5798 | 0.7705 | 0.3325 | 1.7857 | 0.5432 | 0.9274 |
| Toothache | Progression | binary | -0.1821 | 0.3031 | -0.7763 | 0.4120 | 0.8335 | 0.4601 | 1.5099 | 0.5480 | 0.9274 |
| Stress: death of relative | Regression | binary | 0.0742 | 0.1265 | -0.1738 | 0.3222 | 1.0770 | 0.8405 | 1.3801 | 0.5576 | 0.9274 |
| Family history of IBS | Progression | binary | -0.0707 | 0.1255 | -0.3167 | 0.1753 | 0.9317 | 0.7285 | 1.1916 | 0.5732 | 0.9274 |
| Mood swings | Regression | binary | -0.0527 | 0.0944 | -0.2378 | 0.1324 | 0.9487 | 0.7884 | 1.1415 | 0.5769 | 0.9274 |
| Longest manic/irritable episode | Progression | binary | 0.0799 | 0.1441 | -0.2025 | 0.3623 | 1.0832 | 0.8167 | 1.4367 | 0.5791 | 0.9274 |
| Waking too early | Progression | binary | -0.0892 | 0.1688 | -0.4200 | 0.2416 | 0.9147 | 0.6570 | 1.2733 | 0.5972 | 0.9432 |
| Ever depressed for a whole week | Regression | binary | 0.0444 | 0.0912 | -0.1345 | 0.2232 | 1.0454 | 0.8742 | 1.2501 | 0.6266 | 0.9731 |
| Chest pain/discomfort | Regression | binary | -0.0755 | 0.1567 | -0.3827 | 0.2316 | 0.9272 | 0.6820 | 1.2606 | 0.6298 | 0.9731 |
| Mouth ulcers | Progression | binary | -0.0703 | 0.1482 | -0.3608 | 0.2202 | 0.9321 | 0.6972 | 1.2463 | 0.6354 | 0.9731 |
| Restless (anxiety) | Progression | binary | -0.0811 | 0.1762 | -0.4264 | 0.2642 | 0.9221 | 0.6528 | 1.3024 | 0.6453 | 0.9731 |
| Stress: divorce/separation | Progression | binary | -0.1674 | 0.3640 | -0.8809 | 0.5461 | 0.8459 | 0.4144 | 1.7265 | 0.6456 | 0.9731 |
| Ever disinterested for a whole week | Regression | binary | -0.0432 | 0.0969 | -0.2332 | 0.1467 | 0.9577 | 0.7920 | 1.1580 | 0.6555 | 0.9731 |
| Difficulty concentrating (anxiety) | Progression | binary | -0.0850 | 0.1906 | -0.4585 | 0.2885 | 0.9185 | 0.6323 | 1.3344 | 0.6556 | 0.9731 |
| Any vascular/heart problem | Progression | binary | 0.1724 | 0.4079 | -0.6271 | 0.9720 | 1.1882 | 0.5341 | 2.6432 | 0.6725 | 0.9830 |
| Toothache | Regression | binary | -0.1444 | 0.3466 | -0.8237 | 0.5350 | 0.8656 | 0.4388 | 1.7074 | 0.6770 | 0.9830 |
| Risk taking | Progression | binary | -0.0369 | 0.0914 | -0.2161 | 0.1423 | 0.9638 | 0.8057 | 1.1530 | 0.6867 | 0.9830 |
| Back pain 3+ months | Progression | binary | -0.0734 | 0.1825 | -0.4311 | 0.2842 | 0.9292 | 0.6498 | 1.3287 | 0.6874 | 0.9830 |
| Sleep change | Regression | binary | 0.0771 | 0.1983 | -0.3114 | 0.4657 | 1.0802 | 0.7324 | 1.5932 | 0.6972 | 0.9830 |
| Nervous feelings | Regression | binary | -0.0464 | 0.1208 | -0.2831 | 0.1903 | 0.9547 | 0.7534 | 1.2097 | 0.7009 | 0.9830 |
| Stress: financial | Progression | binary | -0.0730 | 0.2070 | -0.4786 | 0.3326 | 0.9296 | 0.6196 | 1.3946 | 0.7243 | 0.9830 |
| Chest pain when walking | Progression | binary | 0.1615 | 0.4904 | -0.7998 | 1.1228 | 1.1753 | 0.4494 | 3.0733 | 0.7420 | 0.9830 |
| Fracture from fall | Regression | binary | -0.0973 | 0.3063 | -0.6978 | 0.5031 | 0.9072 | 0.4977 | 1.6538 | 0.7507 | 0.9830 |
| High blood pressure | Regression | binary | -0.2101 | 0.6724 | -1.5281 | 1.1079 | 0.8105 | 0.2170 | 3.0280 | 0.7547 | 0.9830 |
| Trouble falling asleep | Regression | binary | -0.0585 | 0.1874 | -0.4259 | 0.3089 | 0.9432 | 0.6532 | 1.3619 | 0.7551 | 0.9830 |
| Long-standing illness/disability | Progression | binary | 0.0301 | 0.0967 | -0.1594 | 0.2197 | 1.0306 | 0.8526 | 1.2457 | 0.7554 | 0.9830 |
| Other eye problems | Progression | binary | 0.0329 | 0.1081 | -0.1790 | 0.2449 | 1.0335 | 0.8361 | 1.2774 | 0.7608 | 0.9830 |
| Chest pain/discomfort | Progression | binary | -0.0410 | 0.1366 | -0.3088 | 0.2268 | 0.9598 | 0.7343 | 1.2545 | 0.7638 | 0.9830 |
| Painful gums | Progression | binary | -0.0871 | 0.2903 | -0.6560 | 0.4818 | 0.9166 | 0.5189 | 1.6190 | 0.7641 | 0.9830 |
| Other eye problems | Regression | binary | -0.0337 | 0.1239 | -0.2765 | 0.2092 | 0.9669 | 0.7585 | 1.2326 | 0.7859 | 0.9830 |
| Hearing aid user | Regression | binary | 0.0610 | 0.2270 | -0.3840 | 0.5059 | 1.0629 | 0.6811 | 1.6585 | 0.7883 | 0.9830 |
| Worrier/anxious | Progression | binary | -0.0216 | 0.0808 | -0.1800 | 0.1369 | 0.9787 | 0.8353 | 1.1467 | 0.7897 | 0.9830 |
| Family history of IBS | Regression | binary | 0.0359 | 0.1401 | -0.2386 | 0.3105 | 1.0366 | 0.7877 | 1.3640 | 0.7975 | 0.9830 |
| Leg pain on walking | Regression | binary | -0.0287 | 0.1209 | -0.2656 | 0.2082 | 0.9717 | 0.7668 | 1.2315 | 0.8124 | 0.9830 |
| Tense/highly strung | Regression | binary | 0.0323 | 0.1412 | -0.2445 | 0.3090 | 1.0328 | 0.7831 | 1.3621 | 0.8192 | 0.9830 |
| Macular degeneration | Progression | binary | 0.0770 | 0.3689 | -0.6461 | 0.8001 | 1.0800 | 0.5241 | 2.2257 | 0.8347 | 0.9830 |
| Difficulty concentrating (anxiety) | Regression | binary | -0.0453 | 0.2194 | -0.4753 | 0.3847 | 0.9557 | 0.6217 | 1.4692 | 0.8364 | 0.9830 |
| Worrier/anxious | Regression | binary | -0.0180 | 0.0918 | -0.1979 | 0.1620 | 0.9822 | 0.8204 | 1.1758 | 0.8448 | 0.9830 |
| Sensitivity/hurt feelings | Regression | binary | -0.0174 | 0.0924 | -0.1984 | 0.1636 | 0.9828 | 0.8200 | 1.1778 | 0.8507 | 0.9830 |
| Mouth ulcers | Regression | binary | -0.0304 | 0.1680 | -0.3596 | 0.2988 | 0.9701 | 0.6979 | 1.3483 | 0.8565 | 0.9830 |
| Glaucoma | Progression | binary | 0.0582 | 0.3249 | -0.5786 | 0.6951 | 1.0599 | 0.5607 | 2.0038 | 0.8578 | 0.9830 |
| Alcohol drinker status | Progression | binary | -0.0308 | 0.1751 | -0.3739 | 0.3124 | 0.9697 | 0.6881 | 1.3666 | 0.8605 | 0.9830 |
| Macular degeneration | Regression | binary | -0.0715 | 0.4306 | -0.9155 | 0.7725 | 0.9310 | 0.4003 | 2.1652 | 0.8682 | 0.9830 |
| Worry too long after embarrassment | Regression | binary | 0.0141 | 0.0923 | -0.1668 | 0.1950 | 1.0142 | 0.8464 | 1.2154 | 0.8784 | 0.9830 |
| Sensitivity/hurt feelings | Progression | binary | 0.0121 | 0.0812 | -0.1470 | 0.1713 | 1.0122 | 0.8633 | 1.1868 | 0.8811 | 0.9830 |
| Leg pain on walking | Progression | binary | -0.0154 | 0.1060 | -0.2232 | 0.1924 | 0.9847 | 0.8000 | 1.2121 | 0.8845 | 0.9830 |
| Sensitive stomach | Regression | binary | 0.0188 | 0.1293 | -0.2347 | 0.2723 | 1.0189 | 0.7908 | 1.3129 | 0.8846 | 0.9830 |
| Loneliness/isolation | Regression | binary | -0.0185 | 0.1308 | -0.2748 | 0.2378 | 0.9816 | 0.7597 | 1.2684 | 0.8873 | 0.9830 |
| Stress: death of relative | Progression | binary | 0.0152 | 0.1124 | -0.2051 | 0.2355 | 1.0153 | 0.8146 | 1.2656 | 0.8924 | 0.9830 |
| Tense/highly strung | Progression | binary | 0.0156 | 0.1246 | -0.2286 | 0.2599 | 1.0158 | 0.7957 | 1.2968 | 0.9001 | 0.9830 |
| Ever hyper/manic for 2 days | Regression | binary | -0.0316 | 0.2538 | -0.5290 | 0.4658 | 0.9689 | 0.5892 | 1.5933 | 0.9009 | 0.9830 |
| Glaucoma | Regression | binary | -0.0460 | 0.3762 | -0.7834 | 0.6913 | 0.9550 | 0.4569 | 1.9963 | 0.9026 | 0.9830 |
| Ever highly irritable | Progression | binary | -0.0136 | 0.1172 | -0.2433 | 0.2161 | 0.9865 | 0.7841 | 1.2412 | 0.9076 | 0.9830 |
| Any vascular/heart problem | Regression | binary | -0.0736 | 0.6371 | -1.3223 | 1.1751 | 0.9290 | 0.2665 | 3.2384 | 0.9080 | 0.9830 |
| Probable major depression (single episode) | Regression | binary | -0.0317 | 0.2828 | -0.5861 | 0.5226 | 0.9688 | 0.5565 | 1.6864 | 0.9107 | 0.9830 |
| Loose teeth | Regression | binary | -0.0372 | 0.3900 | -0.8015 | 0.7272 | 0.9635 | 0.4487 | 2.0693 | 0.9241 | 0.9883 |
| Sleep change | Progression | binary | -0.0135 | 0.1730 | -0.3525 | 0.3256 | 0.9866 | 0.7029 | 1.3848 | 0.9378 | 0.9943 |
| IBS diagnosis | Progression | binary | 0.0094 | 0.1407 | -0.2664 | 0.2852 | 1.0094 | 0.7661 | 1.3300 | 0.9470 | 0.9943 |
| Back pain 3+ months | Regression | binary | -0.0089 | 0.2056 | -0.4118 | 0.3940 | 0.9912 | 0.6625 | 1.4829 | 0.9655 | 0.9961 |
| Fed-up feelings | Regression | binary | 0.0033 | 0.0978 | -0.1883 | 0.1949 | 1.0033 | 0.8284 | 1.2152 | 0.9727 | 0.9961 |
| Ever hyper/manic for 2 days | Progression | binary | 0.0071 | 0.2213 | -0.4266 | 0.4408 | 1.0071 | 0.6527 | 1.5539 | 0.9744 | 0.9961 |
| IBS diagnosis | Regression | binary | 0.0051 | 0.1591 | -0.3068 | 0.3169 | 1.0051 | 0.7358 | 1.3729 | 0.9746 | 0.9961 |
| Ever depressed for a whole week | Progression | binary | 0.0012 | 0.0801 | -0.1558 | 0.1583 | 1.0012 | 0.8557 | 1.1715 | 0.9878 | 0.9965 |
| Difficulty stopping worrying | Progression | binary | 0.0046 | 0.3978 | -0.7750 | 0.7842 | 1.0046 | 0.4607 | 2.1907 | 0.9908 | 0.9965 |
| Alcohol drinker status | Regression | binary | -0.0016 | 0.1996 | -0.3928 | 0.3896 | 0.9984 | 0.6752 | 1.4764 | 0.9938 | 0.9965 |
| Nervous feelings | Progression | binary | -0.0008 | 0.1056 | -0.2079 | 0.2063 | 0.9992 | 0.8123 | 1.2291 | 0.9939 | 0.9965 |
| Wheeze in chest | Progression | binary | -0.0005 | 0.1142 | -0.2243 | 0.2233 | 0.9995 | 0.7990 | 1.2502 | 0.9965 | 0.9965 |
| Weight | Regression | continuous | 0.1855 | 0.0480 | 0.0915 | 0.2796 | 1.2039 | 1.0958 | 1.3226 | 0.0001 | 0.0085 |
| Systolic BP (manual) | Progression | continuous | 0.2748 | 0.1012 | 0.0765 | 0.4730 | 1.3162 | 1.0795 | 1.6049 | 0.0066 | 0.2337 |
| BMI | Regression | continuous | 0.1275 | 0.0476 | 0.0342 | 0.2208 | 1.1360 | 1.0347 | 1.2471 | 0.0074 | 0.2337 |
| Diastolic BP (manual) | Regression | continuous | 0.2508 | 0.1047 | 0.0455 | 0.4560 | 1.2850 | 1.0465 | 1.5778 | 0.0166 | 0.2875 |
| BMI | Progression | continuous | 0.0993 | 0.0418 | 0.0173 | 0.1813 | 1.1044 | 1.0174 | 1.1987 | 0.0176 | 0.2875 |
| Systolic BP (manual) | Regression | continuous | 0.2559 | 0.1088 | 0.0427 | 0.4691 | 1.2916 | 1.0436 | 1.5985 | 0.0187 | 0.2875 |
| Diastolic BP (manual) | Progression | continuous | 0.2085 | 0.0972 | 0.0180 | 0.3990 | 1.2318 | 1.0181 | 1.4904 | 0.0320 | 0.3210 |
| Pulse rate | Progression | continuous | 0.1959 | 0.0965 | 0.0066 | 0.3851 | 1.2164 | 1.0067 | 1.4698 | 0.0425 | 0.3988 |
| Longest unenthusiasm/disinterest period | Regression | continuous | -0.1688 | 0.0912 | -0.3475 | 0.0099 | 0.8447 | 0.7065 | 1.0099 | 0.0640 | 0.4483 |
| Duration of vigorous activity | Progression | continuous | -0.0770 | 0.0479 | -0.1709 | 0.0169 | 0.9259 | 0.8429 | 1.0171 | 0.1082 | 0.5552 |
| Duration of walks | Progression | continuous | -0.0570 | 0.0402 | -0.1357 | 0.0217 | 0.9446 | 0.8731 | 1.0219 | 0.1557 | 0.6117 |
| Reaction time (mean) | Progression | continuous | 0.0513 | 0.0425 | -0.0319 | 0.1346 | 1.0527 | 0.9686 | 1.1440 | 0.2267 | 0.6983 |
| Duration of walks | Regression | continuous | -0.0491 | 0.0458 | -0.1389 | 0.0406 | 0.9520 | 0.8703 | 1.0415 | 0.2833 | 0.7883 |
| Pulse rate | Regression | continuous | 0.0979 | 0.1038 | -0.1057 | 0.3014 | 1.1028 | 0.8997 | 1.3518 | 0.3459 | 0.8331 |
| Longest worried/anxious period | Progression | continuous | 0.0690 | 0.1063 | -0.1394 | 0.2774 | 1.0714 | 0.8699 | 1.3197 | 0.5163 | 0.9274 |
| Longest depression period | Regression | continuous | -0.0389 | 0.0705 | -0.1772 | 0.0993 | 0.9618 | 0.8376 | 1.1044 | 0.5811 | 0.9274 |
| Longest depression period | Progression | continuous | 0.0327 | 0.0624 | -0.0896 | 0.1550 | 1.0332 | 0.9143 | 1.1676 | 0.6002 | 0.9432 |
| Longest unenthusiasm/disinterest period | Progression | continuous | 0.0302 | 0.0800 | -0.1266 | 0.1870 | 1.0307 | 0.8811 | 1.2056 | 0.7058 | 0.9830 |
| Reaction time (mean) | Regression | continuous | -0.0119 | 0.0486 | -0.1073 | 0.0834 | 0.9881 | 0.8983 | 1.0870 | 0.8060 | 0.9830 |
| Duration of vigorous activity | Regression | continuous | -0.0105 | 0.0546 | -0.1175 | 0.0965 | 0.9895 | 0.8891 | 1.1013 | 0.8472 | 0.9830 |
| Longest worried/anxious period | Regression | continuous | -0.0123 | 0.1197 | -0.2469 | 0.2222 | 0.9877 | 0.7812 | 1.2489 | 0.9180 | 0.9863 |
| Walking pace | Progression | ordered | -0.2864 | 0.0734 | -0.4303 | -0.1424 | 0.7510 | 0.6503 | 0.8673 | 0.0001 | 0.0085 |
| Tiredness/lethargy (2 weeks) | Progression | ordered | -0.1728 | 0.0511 | -0.2730 | -0.0726 | 0.8413 | 0.7611 | 0.9300 | 0.0007 | 0.0419 |
| Walking pace | Regression | ordered | -0.2372 | 0.0831 | -0.4001 | -0.0742 | 0.7889 | 0.6702 | 0.9285 | 0.0043 | 0.2005 |
| Puzzle score | Progression | ordered | -0.0539 | 0.0217 | -0.0965 | -0.0114 | 0.9475 | 0.9080 | 0.9887 | 0.0130 | 0.2875 |
| Max digits remembered | Regression | ordered | -0.1054 | 0.0441 | -0.1918 | -0.0190 | 0.9000 | 0.8255 | 0.9811 | 0.0168 | 0.2875 |
| General happiness | Regression | ordered | -0.1486 | 0.0679 | -0.2817 | -0.0156 | 0.8619 | 0.7545 | 0.9846 | 0.0286 | 0.3210 |
| Max digits remembered | Progression | ordered | -0.0833 | 0.0384 | -0.1586 | -0.0080 | 0.9201 | 0.8533 | 0.9921 | 0.0302 | 0.3210 |
| Puzzle score | Regression | ordered | -0.0473 | 0.0243 | -0.0949 | 0.0003 | 0.9538 | 0.9095 | 1.0003 | 0.0515 | 0.3988 |
| Overall health rating | Regression | ordered | 0.1269 | 0.0711 | -0.0126 | 0.2663 | 1.1353 | 0.9875 | 1.3051 | 0.0745 | 0.4918 |
| General happiness | Progression | ordered | -0.1041 | 0.0596 | -0.2208 | 0.0127 | 0.9012 | 0.8018 | 1.0128 | 0.0806 | 0.5173 |
| Tiredness/lethargy (2 weeks) | Regression | ordered | -0.0997 | 0.0578 | -0.2129 | 0.0136 | 0.9051 | 0.8082 | 1.0137 | 0.0846 | 0.5282 |
| Able to confide | Progression | ordered | -0.0385 | 0.0225 | -0.0826 | 0.0056 | 0.9622 | 0.9208 | 1.0056 | 0.0870 | 0.5288 |
| Tenseness/restlessness (2 weeks) | Progression | ordered | -0.1284 | 0.0757 | -0.2767 | 0.0200 | 0.8795 | 0.7583 | 1.0202 | 0.0899 | 0.5324 |
| Able to confide | Regression | ordered | -0.0414 | 0.0254 | -0.0911 | 0.0083 | 0.9595 | 0.9129 | 1.0084 | 0.1029 | 0.5552 |
| Neuroticism | Progression | ordered | -0.0214 | 0.0135 | -0.0479 | 0.0052 | 0.9789 | 0.9532 | 1.0052 | 0.1144 | 0.5617 |
| Strenuous sports duration | Regression | ordered | 0.1650 | 0.1056 | -0.0420 | 0.3721 | 1.1794 | 0.9589 | 1.4507 | 0.1182 | 0.5617 |
| Back pain (3 months) | Progression | ordered | -0.0974 | 0.0645 | -0.2238 | 0.0290 | 0.9072 | 0.7995 | 1.0295 | 0.1311 | 0.5825 |
| Current smoking | Progression | ordered | 0.2129 | 0.1528 | -0.0866 | 0.5123 | 1.2372 | 0.9171 | 1.6691 | 0.1635 | 0.6117 |
| Vigorous activity days/weeks | Progression | ordered | -0.0291 | 0.0211 | -0.0705 | 0.0124 | 0.9714 | 0.9319 | 1.0125 | 0.1695 | 0.6117 |
| Moderate activity days/weeks | Regression | ordered | 0.0278 | 0.0210 | -0.0134 | 0.0690 | 1.0282 | 0.9867 | 1.0714 | 0.1858 | 0.6336 |
| Falls in last year | Progression | ordered | -0.0982 | 0.0787 | -0.2524 | 0.0559 | 0.9064 | 0.7769 | 1.0575 | 0.2117 | 0.6886 |
| Strenuous sports frequency | Progression | ordered | 0.0998 | 0.0864 | -0.0695 | 0.2691 | 1.1049 | 0.9328 | 1.3088 | 0.2480 | 0.7345 |
| Tenseness/restlessness (2 weeks) | Regression | ordered | -0.0987 | 0.0864 | -0.2680 | 0.0706 | 0.9060 | 0.7649 | 1.0732 | 0.2532 | 0.7404 |
| Chest pain during activity | Regression | ordered | -0.6516 | 0.6466 | -1.9189 | 0.6157 | 0.5212 | 0.1468 | 1.8510 | 0.3136 | 0.8139 |
| Unenthusiasm (2 weeks) | Regression | ordered | 0.0839 | 0.0922 | -0.0968 | 0.2647 | 1.0876 | 0.9077 | 1.3030 | 0.3627 | 0.8463 |
| Fluid intelligence | Progression | ordered | -0.0186 | 0.0207 | -0.0592 | 0.0220 | 0.9815 | 0.9425 | 1.0222 | 0.3685 | 0.8511 |
| Back pain (3 months) | Regression | ordered | -0.0629 | 0.0729 | -0.2058 | 0.0800 | 0.9391 | 0.8140 | 1.0833 | 0.3885 | 0.8595 |
| Unenthusiasm (2 weeks) | Progression | ordered | -0.0712 | 0.0843 | -0.2365 | 0.0941 | 0.9313 | 0.7893 | 1.0987 | 0.3984 | 0.8595 |
| Strenuous sports frequency | Regression | ordered | 0.0813 | 0.0991 | -0.1130 | 0.2756 | 1.0847 | 0.8931 | 1.3173 | 0.4122 | 0.8595 |
| Chest pain outside activity | Progression | ordered | 0.3293 | 0.4021 | -0.4588 | 1.1174 | 1.3900 | 0.6321 | 3.0568 | 0.4128 | 0.8595 |
| Alcohol intake frequency | Progression | ordered | -0.0227 | 0.0291 | -0.0798 | 0.0343 | 0.9775 | 0.9233 | 1.0349 | 0.4343 | 0.8800 |
| Depressed mood (2 weeks) | Regression | ordered | 0.0609 | 0.0908 | -0.1171 | 0.2389 | 1.0628 | 0.8895 | 1.2698 | 0.5025 | 0.9274 |
| Moderate activity duration | Progression | ordered | 0.0329 | 0.0516 | -0.0684 | 0.1341 | 1.0334 | 0.9339 | 1.1435 | 0.5247 | 0.9274 |
| Neuroticism | Regression | ordered | -0.0096 | 0.0153 | -0.0395 | 0.0204 | 0.9905 | 0.9612 | 1.0206 | 0.5300 | 0.9274 |
| Falls in last year | Regression | ordered | -0.0556 | 0.0892 | -0.2304 | 0.1192 | 0.9459 | 0.7942 | 1.1266 | 0.5330 | 0.9274 |
| Light DIY duration | Progression | ordered | 0.0222 | 0.0359 | -0.0482 | 0.0926 | 1.0225 | 0.9530 | 1.0970 | 0.5363 | 0.9274 |
| Current smoking | Regression | ordered | 0.1072 | 0.1745 | -0.2347 | 0.4492 | 1.1132 | 0.7908 | 1.5670 | 0.5388 | 0.9274 |
| Overall health rating | Progression | ordered | 0.0372 | 0.0629 | -0.0861 | 0.1605 | 1.0379 | 0.9175 | 1.1741 | 0.5541 | 0.9274 |
| Moderate activity days/weeks | Progression | ordered | 0.0107 | 0.0185 | -0.0255 | 0.0469 | 1.0107 | 0.9748 | 1.0480 | 0.5633 | 0.9274 |
| Alcohol intake frequency | Regression | ordered | -0.0190 | 0.0331 | -0.0838 | 0.0459 | 0.9812 | 0.9196 | 1.0470 | 0.5667 | 0.9274 |
| Fluid intelligence | Regression | ordered | -0.0131 | 0.0237 | -0.0596 | 0.0334 | 0.9870 | 0.9421 | 1.0340 | 0.5806 | 0.9274 |
| Chest pain during activity | Progression | ordered | -0.2122 | 0.3856 | -0.9680 | 0.5436 | 0.8088 | 0.3799 | 1.7222 | 0.5821 | 0.9274 |
| Daytime dozing | Progression | ordered | 0.0389 | 0.0862 | -0.1300 | 0.2078 | 1.0397 | 0.8781 | 1.2310 | 0.6516 | 0.9731 |
| Happiness | Progression | ordered | -0.0236 | 0.0577 | -0.1367 | 0.0895 | 0.9767 | 0.8722 | 1.0936 | 0.6823 | 0.9830 |
| Depressed mood (2 weeks) | Progression | ordered | -0.0301 | 0.0818 | -0.1904 | 0.1302 | 0.9703 | 0.8266 | 1.1390 | 0.7128 | 0.9830 |
| Vigorous activity days/weeks | Regression | ordered | 0.0086 | 0.0238 | -0.0382 | 0.0553 | 1.0086 | 0.9626 | 1.0568 | 0.7195 | 0.9830 |
| Strenuous sports duration | Progression | ordered | 0.0309 | 0.0946 | -0.1545 | 0.2162 | 1.0314 | 0.8569 | 1.2414 | 0.7440 | 0.9830 |
| Sleeplessness/insomnia | Progression | ordered | -0.0173 | 0.0542 | -0.1237 | 0.0890 | 0.9828 | 0.8837 | 1.0930 | 0.7491 | 0.9830 |
| Daytime dozing | Regression | ordered | 0.0250 | 0.0979 | -0.1669 | 0.2168 | 1.0253 | 0.8463 | 1.2422 | 0.7987 | 0.9830 |
| Sleeplessness/insomnia | Regression | ordered | -0.0147 | 0.0616 | -0.1355 | 0.1061 | 0.9854 | 0.8733 | 1.1119 | 0.8111 | 0.9830 |
| Happiness with health | Regression | ordered | 0.0137 | 0.0578 | -0.0997 | 0.1270 | 1.0138 | 0.9051 | 1.1355 | 0.8129 | 0.9830 |
| Happiness | Regression | ordered | -0.0133 | 0.0656 | -0.1418 | 0.1152 | 0.9868 | 0.8678 | 1.1221 | 0.8395 | 0.9830 |
| Moderate activity duration | Regression | ordered | 0.0106 | 0.0589 | -0.1048 | 0.1260 | 1.0107 | 0.9005 | 1.1343 | 0.8569 | 0.9830 |
| Depression episodes | Regression | ordered | 0.0105 | 0.0806 | -0.1475 | 0.1686 | 1.0106 | 0.8628 | 1.1837 | 0.8961 | 0.9830 |
| Depression episodes | Progression | ordered | 0.0055 | 0.0714 | -0.1344 | 0.1454 | 1.0055 | 0.8742 | 1.1565 | 0.9389 | 0.9943 |
| Happiness with health | Progression | ordered | 0.0035 | 0.0510 | -0.0964 | 0.1035 | 1.0035 | 0.9081 | 1.1090 | 0.9451 | 0.9943 |
| Light DIY duration | Regression | ordered | 0.0022 | 0.0414 | -0.0789 | 0.0833 | 1.0022 | 0.9242 | 1.0869 | 0.9567 | 0.9961 |
| Chest pain outside activity | Regression | ordered | 0.0175 | 0.5210 | -1.0037 | 1.0386 | 1.0176 | 0.3665 | 2.8254 | 0.9733 | 0.9961 |
| Weight change during worst depression episode | Progression | unordered | -0.0844 | 0.0660 | -0.2138 | 0.0451 | 0.9191 | 0.8075 | 1.0461 | 0.2015 | 0.6744 |
| Weight change during worst depression episode | Regression | unordered | -0.0766 | 0.0751 | -0.2237 | 0.0705 | 0.9263 | 0.7996 | 1.0731 | 0.3077 | 0.8078 |
| Bipolar and major depression status | Progression | unordered | 0.0393 | 0.0885 | -0.1342 | 0.2128 | 1.0401 | 0.8744 | 1.2371 | 0.6572 | 0.9731 |
| Major dietary changes (5 years) | Regression | unordered | 0.0163 | 0.0497 | -0.0810 | 0.1137 | 1.0165 | 0.9222 | 1.1204 | 0.7423 | 0.9830 |
| Bipolar and major depression status | Regression | unordered | 0.0230 | 0.1030 | -0.1789 | 0.2249 | 1.0233 | 0.8362 | 1.2523 | 0.8232 | 0.9830 |
| Major dietary changes (5 years) | Progression | unordered | -0.0089 | 0.0438 | -0.0948 | 0.0770 | 0.9911 | 0.9095 | 1.0801 | 0.8391 | 0.9830 |

*Abbreviations:* BMI, body mass index; BP, blood pressure; CI, confidence interval; FDR, false-discovery rate corrected; OR, odds ratio; SE, standard error; UKB, UK Biobank.

**eAppendix 8.** All partially and fully adjusted multinomial regression results for baseline clinical phenotypes associated with subsequent WMH volume not surviving FDR-correction at *p*<0.05.

| **UKB Phenotype** | **Comparison (versus Stable)** | **Predictor Type** | **Model** | **Estimate** | **SE** | **CI lower** | **CI upper** | **OR** | **OR lower** | **OR upper** | ***P*** | **Adjusted *P*** |
| --- | --- | --- | --- | --- | --- | --- | --- | --- | --- | --- | --- | --- |
| Hip pain 3+ months | Progression | binary | Fully adjusted | -1.3952 | 0.4323 | -2.2424 | -0.5479 | 0.2478 | 0.1062 | 0.5781 | 0.0012 | 0.5778 |
| Walking pace | Progression | ordered | Partially adjusted | -0.2299 | 0.0764 | -0.3796 | -0.0801 | 0.7947 | 0.6841 | 0.9231 | 0.0026 | 0.6095 |
| Hip pain 3+ months | Progression | binary | Partially adjusted | -0.9298 | 0.3364 | -1.5892 | -0.2705 | 0.3946 | 0.2041 | 0.7630 | 0.0057 | 0.8808 |
| Weight | Regression | continuous | Partially adjusted | 0.1413 | 0.0568 | 0.0299 | 0.2526 | 1.1518 | 1.0304 | 1.2874 | 0.0129 | 0.8942 |
| Diastolic BP (manual) | Regression | continuous | Partially adjusted | 0.2362 | 0.1111 | 0.0184 | 0.4541 | 1.2665 | 1.0186 | 1.5747 | 0.0335 | 0.8942 |
| Walking pace | Regression | ordered | Partially adjusted | -0.1910 | 0.0855 | -0.3585 | -0.0235 | 0.8261 | 0.6987 | 0.9768 | 0.0254 | 0.8942 |
| Stress: serious illness/assault self | Regression | binary | Partially adjusted | 0.4103 | 0.2182 | -0.0173 | 0.8380 | 1.5073 | 0.9828 | 2.3117 | 0.0600 | 0.8942 |
| Hip pain 3+ months | Regression | binary | Partially adjusted | -0.6883 | 0.3714 | -1.4163 | 0.0396 | 0.5024 | 0.2426 | 1.0404 | 0.0638 | 0.8942 |
| Overall health rating | Regression | ordered | Partially adjusted | 0.1483 | 0.0722 | 0.0068 | 0.2899 | 1.1599 | 1.0068 | 1.3363 | 0.0400 | 0.8942 |
| BMI | Regression | continuous | Partially adjusted | 0.1048 | 0.0489 | 0.0090 | 0.2007 | 1.1105 | 1.0091 | 1.2222 | 0.0320 | 0.8942 |
| Unenthusiasm (2 weeks) | Regression | ordered | Partially adjusted | 0.1850 | 0.0958 | -0.0028 | 0.3729 | 1.2033 | 0.9972 | 1.4519 | 0.0535 | 0.8942 |
| Max digits remembered | Regression | ordered | Partially adjusted | -0.0890 | 0.0458 | -0.1788 | 0.0009 | 0.9149 | 0.8363 | 1.0009 | 0.0523 | 0.8942 |
| Reaction time (mean) | Regression | continuous | Partially adjusted | -0.0981 | 0.0528 | -0.2016 | 0.0055 | 0.9066 | 0.8174 | 1.0055 | 0.0634 | 0.8942 |
| BMI | Progression | continuous | Partially adjusted | 0.0907 | 0.0433 | 0.0058 | 0.1756 | 1.0949 | 1.0059 | 1.1919 | 0.0362 | 0.8942 |
| Headaches 3+ months | Progression | binary | Partially adjusted | 0.5323 | 0.2360 | 0.0697 | 0.9949 | 1.7029 | 1.0722 | 2.7045 | 0.0241 | 0.8942 |
| Cataract | Progression | binary | Partially adjusted | 0.3261 | 0.1730 | -0.0131 | 0.6652 | 1.3855 | 0.9870 | 1.9448 | 0.0595 | 0.8942 |
| Diastolic BP (manual) | Progression | continuous | Partially adjusted | 0.1983 | 0.1040 | -0.0054 | 0.4021 | 1.2194 | 0.9946 | 1.4950 | 0.0564 | 0.8942 |
| Pulse rate | Progression | continuous | Partially adjusted | 0.2315 | 0.1028 | 0.0300 | 0.4330 | 1.2605 | 1.0305 | 1.5419 | 0.0243 | 0.8942 |
| Strenuous sports frequency | Regression | ordered | Partially adjusted | 0.1248 | 0.1008 | -0.0727 | 0.3224 | 1.1330 | 0.9299 | 1.3804 | 0.2154 | 0.8942 |
| Pulse rate | Regression | continuous | Partially adjusted | 0.1534 | 0.1097 | -0.0615 | 0.3683 | 1.1658 | 0.9403 | 1.4453 | 0.1618 | 0.8942 |
| Systolic BP (manual) | Regression | continuous | Partially adjusted | 0.1845 | 0.1185 | -0.0477 | 0.4167 | 1.2026 | 0.9534 | 1.5170 | 0.1194 | 0.8942 |
| Puzzle score | Regression | ordered | Partially adjusted | -0.0347 | 0.0256 | -0.0849 | 0.0156 | 0.9659 | 0.9186 | 1.0157 | 0.1761 | 0.8942 |
| Bleeding gums | Regression | binary | Partially adjusted | -0.1748 | 0.1450 | -0.4589 | 0.1093 | 0.8396 | 0.6320 | 1.1155 | 0.2279 | 0.8942 |
| Painful gums | Regression | binary | Partially adjusted | -0.5919 | 0.3926 | -1.3615 | 0.1776 | 0.5533 | 0.2563 | 1.1943 | 0.1316 | 0.8942 |
| Vision loss from injury | Regression | binary | Partially adjusted | 1.2424 | 1.1040 | -0.9214 | 3.4062 | 3.4638 | 0.3979 | 30.1494 | 0.2604 | 0.8942 |
| Diabetes-related eye disease | Regression | binary | Partially adjusted | -0.7440 | 0.7686 | -2.2505 | 0.7625 | 0.4752 | 0.1053 | 2.1437 | 0.3331 | 0.8942 |
| Stress: financial | Regression | binary | Partially adjusted | -0.4120 | 0.2692 | -0.9396 | 0.1157 | 0.6624 | 0.3908 | 1.1227 | 0.1260 | 0.8942 |
| Stress: death of spouse | Regression | binary | Partially adjusted | 1.8609 | 1.0512 | -0.1995 | 3.9212 | 6.4295 | 0.8192 | 50.4633 | 0.0767 | 0.8942 |
| Stress: death of relative | Regression | binary | Partially adjusted | 0.1221 | 0.1312 | -0.1351 | 0.3792 | 1.1298 | 0.8736 | 1.4611 | 0.3522 | 0.8942 |
| Chest pain during activity | Regression | ordered | Partially adjusted | -0.6414 | 0.6717 | -1.9579 | 0.6751 | 0.5266 | 0.1412 | 1.9642 | 0.3396 | 0.8942 |
| Longest unenthusiasm/disinterest period | Regression | continuous | Partially adjusted | -0.1416 | 0.0948 | -0.3274 | 0.0443 | 0.8680 | 0.7208 | 1.0453 | 0.1355 | 0.8942 |
| Ever depressed â‰¥1 week | Regression | binary | Partially adjusted | 0.1326 | 0.0945 | -0.0528 | 0.3179 | 1.1417 | 0.9486 | 1.3742 | 0.1609 | 0.8942 |
| Facial pain 3+ months | Regression | binary | Partially adjusted | 1.4837 | 1.1555 | -0.7810 | 3.7485 | 4.4094 | 0.4579 | 42.4577 | 0.1991 | 0.8942 |
| Headaches 3+ months | Regression | binary | Partially adjusted | 0.3547 | 0.2671 | -0.1687 | 0.8782 | 1.4258 | 0.8448 | 2.4065 | 0.1841 | 0.8942 |
| Abdominal pain 3+ months | Regression | binary | Partially adjusted | -0.4532 | 0.4299 | -1.2958 | 0.3895 | 0.6356 | 0.2737 | 1.4762 | 0.2919 | 0.8942 |
| Chest pain when walking | Regression | binary | Partially adjusted | 0.7446 | 0.5647 | -0.3623 | 1.8514 | 2.1055 | 0.6961 | 6.3689 | 0.1873 | 0.8942 |
| Neck/shoulder pain 3+ months | Regression | binary | Partially adjusted | -0.3082 | 0.2566 | -0.8112 | 0.1948 | 0.7348 | 0.4443 | 1.2151 | 0.2298 | 0.8942 |
| Hearing aid user | Regression | binary | Partially adjusted | -0.2152 | 0.2323 | -0.6705 | 0.2401 | 0.8064 | 0.5114 | 1.2714 | 0.3542 | 0.8942 |
| Wheeze in chest | Regression | binary | Partially adjusted | -0.2301 | 0.1373 | -0.4993 | 0.0390 | 0.7944 | 0.6070 | 1.0398 | 0.0938 | 0.8942 |
| Long-standing illness/disability | Regression | binary | Partially adjusted | 0.1110 | 0.1112 | -0.1070 | 0.3289 | 1.1174 | 0.8985 | 1.3895 | 0.3183 | 0.8942 |
| Family history of IBS | Regression | binary | Partially adjusted | 0.1470 | 0.1442 | -0.1356 | 0.4296 | 1.1583 | 0.8732 | 1.5366 | 0.3080 | 0.8942 |
| IBS diagnosis | Regression | binary | Partially adjusted | 0.1522 | 0.1633 | -0.1678 | 0.4723 | 1.1644 | 0.8455 | 1.6037 | 0.3512 | 0.8942 |
| Waking too early | Regression | binary | Partially adjusted | 0.2059 | 0.2002 | -0.1866 | 0.5983 | 1.2286 | 0.8298 | 1.8191 | 0.3039 | 0.8942 |
| Sleep change | Regression | binary | Partially adjusted | 0.1915 | 0.2032 | -0.2067 | 0.5897 | 1.2111 | 0.8133 | 1.8035 | 0.3458 | 0.8942 |
| Depressed mood (2 weeks) | Regression | ordered | Partially adjusted | 0.1629 | 0.0948 | -0.0229 | 0.3487 | 1.1769 | 0.9774 | 1.4172 | 0.0857 | 0.8942 |
| General happiness | Regression | ordered | Partially adjusted | -0.0958 | 0.0696 | -0.2323 | 0.0406 | 0.9086 | 0.7927 | 1.0415 | 0.1687 | 0.8942 |
| Sleep problems (anxiety) | Regression | binary | Partially adjusted | 0.2812 | 0.2504 | -0.2097 | 0.7720 | 1.3247 | 0.8108 | 2.1641 | 0.2616 | 0.8942 |
| Risk taking | Regression | binary | Partially adjusted | -0.1424 | 0.1088 | -0.3556 | 0.0708 | 0.8673 | 0.7008 | 1.0734 | 0.1906 | 0.8942 |
| Worry too long after embarrassment | Regression | binary | Partially adjusted | 0.1110 | 0.0959 | -0.0769 | 0.2990 | 1.1174 | 0.9260 | 1.3485 | 0.2468 | 0.8942 |
| Tense/highly strung | Regression | binary | Partially adjusted | 0.1738 | 0.1464 | -0.1132 | 0.4608 | 1.1898 | 0.8930 | 1.5854 | 0.2353 | 0.8942 |
| Worrier/anxious | Regression | binary | Partially adjusted | 0.1421 | 0.0960 | -0.0460 | 0.3303 | 1.1527 | 0.9550 | 1.3914 | 0.1388 | 0.8942 |
| Fed-up feelings | Regression | binary | Partially adjusted | 0.1311 | 0.1016 | -0.0680 | 0.3301 | 1.1401 | 0.9343 | 1.3912 | 0.1968 | 0.8942 |
| Sensitivity/hurt feelings | Regression | binary | Partially adjusted | 0.0939 | 0.0965 | -0.0951 | 0.2830 | 1.0985 | 0.9093 | 1.3271 | 0.3301 | 0.8942 |
| Irritability | Regression | binary | Partially adjusted | 0.1345 | 0.1080 | -0.0771 | 0.3461 | 1.1440 | 0.9258 | 1.4135 | 0.2128 | 0.8942 |
| Snoring | Regression | binary | Partially adjusted | -0.1043 | 0.1018 | -0.3038 | 0.0952 | 0.9010 | 0.7380 | 1.0999 | 0.3056 | 0.8942 |
| Major dietary changes (5 years) | Regression | unordered | Partially adjusted | 0.0547 | 0.0511 | -0.0455 | 0.1549 | 1.0562 | 0.9555 | 1.1675 | 0.2846 | 0.8942 |
| Current smoking | Progression | ordered | Partially adjusted | 0.2514 | 0.1581 | -0.0584 | 0.5612 | 1.2858 | 0.9433 | 1.7528 | 0.1117 | 0.8942 |
| Major dietary changes (5 years) | Progression | unordered | Partially adjusted | 0.0481 | 0.0458 | -0.0417 | 0.1379 | 1.0493 | 0.9592 | 1.1478 | 0.2935 | 0.8942 |
| Sensitivity/hurt feelings | Progression | binary | Partially adjusted | 0.0941 | 0.0859 | -0.0742 | 0.2625 | 1.0987 | 0.9285 | 1.3002 | 0.2730 | 0.8942 |
| Nervous feelings | Progression | binary | Partially adjusted | 0.1144 | 0.1104 | -0.1020 | 0.3308 | 1.1212 | 0.9030 | 1.3920 | 0.3002 | 0.8942 |
| Worrier/anxious | Progression | binary | Partially adjusted | 0.1055 | 0.0857 | -0.0625 | 0.2735 | 1.1113 | 0.9394 | 1.3145 | 0.2183 | 0.8942 |
| Tense/highly strung | Progression | binary | Partially adjusted | 0.1525 | 0.1316 | -0.1054 | 0.4105 | 1.1648 | 0.9000 | 1.5075 | 0.2464 | 0.8942 |
| Fluid intelligence | Progression | ordered | Partially adjusted | -0.0336 | 0.0221 | -0.0769 | 0.0096 | 0.9669 | 0.9260 | 1.0097 | 0.1274 | 0.8942 |
| Reaction time (mean) | Progression | continuous | Partially adjusted | -0.0544 | 0.0467 | -0.1459 | 0.0372 | 0.9471 | 0.8642 | 1.0379 | 0.2445 | 0.8942 |
| Probable major depression (single episode) | Progression | binary | Partially adjusted | -0.2690 | 0.2617 | -0.7819 | 0.2439 | 0.7642 | 0.4576 | 1.2762 | 0.3040 | 0.8942 |
| Max digits remembered | Progression | ordered | Partially adjusted | -0.0536 | 0.0406 | -0.1332 | 0.0259 | 0.9478 | 0.8753 | 1.0262 | 0.1862 | 0.8942 |
| Longest worried/anxious period | Progression | continuous | Partially adjusted | 0.1100 | 0.1112 | -0.1080 | 0.3280 | 1.1163 | 0.8976 | 1.3882 | 0.3226 | 0.8942 |
| Sleep problems (anxiety) | Progression | binary | Partially adjusted | 0.3245 | 0.2224 | -0.1115 | 0.7605 | 1.3833 | 0.8945 | 2.1393 | 0.1446 | 0.8942 |
| Thoughts of death (depression) | Progression | binary | Partially adjusted | -0.1766 | 0.1328 | -0.4370 | 0.0837 | 0.8381 | 0.6460 | 1.0873 | 0.1836 | 0.8942 |
| Depressed mood (2 weeks) | Progression | ordered | Partially adjusted | 0.1092 | 0.0864 | -0.0601 | 0.2785 | 1.1154 | 0.9417 | 1.3211 | 0.2060 | 0.8942 |
| Weight change during worst depression episode | Progression | unordered | Partially adjusted | -0.0712 | 0.0707 | -0.2097 | 0.0673 | 0.9313 | 0.8108 | 1.0696 | 0.3137 | 0.8942 |
| Tiredness/lethargy (2 weeks) | Progression | ordered | Partially adjusted | -0.0709 | 0.0539 | -0.1767 | 0.0348 | 0.9315 | 0.8381 | 1.0354 | 0.1885 | 0.8942 |
| Weight | Progression | continuous | Partially adjusted | 0.0671 | 0.0505 | -0.0319 | 0.1661 | 1.0694 | 0.9686 | 1.1807 | 0.1842 | 0.8942 |
| Back pain (3 months) | Progression | ordered | Partially adjusted | -0.0634 | 0.0673 | -0.1953 | 0.0686 | 0.9386 | 0.8226 | 1.0710 | 0.3465 | 0.8942 |
| Overall health rating | Progression | ordered | Partially adjusted | 0.0797 | 0.0649 | -0.0475 | 0.2069 | 1.0830 | 0.9536 | 1.2298 | 0.2194 | 0.8942 |
| Falls in last year | Progression | ordered | Partially adjusted | -0.0890 | 0.0824 | -0.2505 | 0.0725 | 0.9148 | 0.7784 | 1.0752 | 0.2800 | 0.8942 |
| Fracture from fall | Progression | binary | Partially adjusted | -0.2928 | 0.2922 | -0.8655 | 0.2800 | 0.7462 | 0.4208 | 1.3231 | 0.3164 | 0.8942 |
| Neck/shoulder pain 3+ months | Progression | binary | Partially adjusted | -0.2466 | 0.2294 | -0.6963 | 0.2031 | 0.7815 | 0.4984 | 1.2252 | 0.2824 | 0.8942 |
| Abdominal pain 3+ months | Progression | binary | Partially adjusted | -0.6582 | 0.4073 | -1.4565 | 0.1401 | 0.5178 | 0.2331 | 1.1503 | 0.1061 | 0.8942 |
| Happiness | Progression | ordered | Partially adjusted | 0.0602 | 0.0602 | -0.0579 | 0.1782 | 1.0620 | 0.9438 | 1.1951 | 0.3178 | 0.8942 |
| Ever depressed for a whole week | Progression | binary | Partially adjusted | 0.1011 | 0.0842 | -0.0640 | 0.2662 | 1.1064 | 0.9380 | 1.3049 | 0.2300 | 0.8942 |
| Longest depression period | Progression | continuous | Partially adjusted | 0.0715 | 0.0653 | -0.0564 | 0.1995 | 1.0742 | 0.9452 | 1.2207 | 0.2730 | 0.8942 |
| Ever highly irritable | Progression | binary | Partially adjusted | 0.1472 | 0.1225 | -0.0930 | 0.3873 | 1.1585 | 0.9112 | 1.4730 | 0.2297 | 0.8942 |
| Longest unenthusiasm/disinterest period | Progression | continuous | Partially adjusted | 0.0895 | 0.0847 | -0.0765 | 0.2555 | 1.0936 | 0.9264 | 1.2911 | 0.2905 | 0.8942 |
| Chest pain outside activity | Progression | ordered | Partially adjusted | 0.4857 | 0.4137 | -0.3252 | 1.2966 | 1.6253 | 0.7224 | 3.6568 | 0.2404 | 0.8942 |
| Stress: serious illness/assault self | Progression | binary | Partially adjusted | 0.2885 | 0.2028 | -0.1091 | 0.6861 | 1.3344 | 0.8967 | 1.9859 | 0.1550 | 0.8942 |
| Stress: serious illness/assault relative | Regression | binary | Partially adjusted | -0.2058 | 0.1349 | -0.4702 | 0.0586 | 0.8140 | 0.6249 | 1.0603 | 0.1270 | 0.8942 |
| Stress: serious illness/assault relative | Progression | binary | Partially adjusted | -0.2161 | 0.1182 | -0.4478 | 0.0157 | 0.8057 | 0.6390 | 1.0158 | 0.0676 | 0.8942 |
| Stress: death of relative | Progression | binary | Partially adjusted | 0.1191 | 0.1180 | -0.1122 | 0.3503 | 1.1264 | 0.8939 | 1.4196 | 0.3130 | 0.8942 |
| Stress: death of spouse | Progression | binary | Partially adjusted | 1.5389 | 1.0330 | -0.4858 | 3.5637 | 4.6596 | 0.6152 | 35.2925 | 0.1363 | 0.8942 |
| Other serious eye condition | Progression | binary | Partially adjusted | -0.5337 | 0.3162 | -1.1534 | 0.0860 | 0.5864 | 0.3156 | 1.0898 | 0.0914 | 0.8942 |
| Bleeding gums | Progression | binary | Partially adjusted | -0.2183 | 0.1277 | -0.4686 | 0.0320 | 0.8039 | 0.6259 | 1.0325 | 0.0874 | 0.8942 |
| High blood pressure | Progression | binary | Partially adjusted | -0.4371 | 0.4586 | -1.3360 | 0.4617 | 0.6459 | 0.2629 | 1.5868 | 0.3405 | 0.8942 |
| Puzzle score | Progression | ordered | Partially adjusted | -0.0244 | 0.0232 | -0.0699 | 0.0212 | 0.9759 | 0.9325 | 1.0214 | 0.2946 | 0.8942 |
| Duration of walks | Progression | continuous | Partially adjusted | -0.0408 | 0.0417 | -0.1225 | 0.0408 | 0.9600 | 0.8847 | 1.0417 | 0.3269 | 0.8942 |
| Vigorous activity days/weeks | Progression | ordered | Partially adjusted | -0.0295 | 0.0222 | -0.0730 | 0.0139 | 0.9709 | 0.9296 | 1.0140 | 0.1826 | 0.8942 |
| Duration of vigorous activity | Progression | continuous | Partially adjusted | -0.0610 | 0.0501 | -0.1592 | 0.0371 | 0.9408 | 0.8529 | 1.0378 | 0.2229 | 0.8942 |
| Systolic BP (manual) | Progression | continuous | Partially adjusted | 0.1800 | 0.1105 | -0.0367 | 0.3966 | 1.1972 | 0.9640 | 1.4868 | 0.1035 | 0.8942 |
| Strenuous sports frequency | Progression | ordered | Partially adjusted | 0.1339 | 0.0897 | -0.0419 | 0.3097 | 1.1433 | 0.9590 | 1.3630 | 0.1355 | 0.8942 |
| Duration of walks | Regression | continuous | Partially adjusted | -0.0398 | 0.0468 | -0.1316 | 0.0520 | 0.9610 | 0.8767 | 1.0533 | 0.3951 | 0.8942 |
| Other serious eye condition | Regression | binary | Partially adjusted | -0.3048 | 0.3497 | -0.9903 | 0.3807 | 0.7373 | 0.3715 | 1.4633 | 0.3835 | 0.8942 |
| Other eye problems | Regression | binary | Partially adjusted | -0.1103 | 0.1276 | -0.3604 | 0.1398 | 0.8955 | 0.6974 | 1.1500 | 0.3873 | 0.8942 |
| Difficulty stopping worrying | Regression | binary | Partially adjusted | -0.3777 | 0.4471 | -1.2541 | 0.4986 | 0.6854 | 0.2853 | 1.6464 | 0.3982 | 0.8942 |
| Tiredness (depression) | Regression | binary | Partially adjusted | -0.1856 | 0.2016 | -0.5807 | 0.2095 | 0.8306 | 0.5595 | 1.2330 | 0.3571 | 0.8942 |
| Thoughts of death (depression) | Regression | binary | Partially adjusted | -0.1310 | 0.1478 | -0.4206 | 0.1586 | 0.8772 | 0.6567 | 1.1719 | 0.3753 | 0.8942 |
| Neuroticism | Regression | ordered | Partially adjusted | 0.0137 | 0.0159 | -0.0176 | 0.0449 | 1.0138 | 0.9826 | 1.0459 | 0.3908 | 0.8942 |
| Smoking status | Regression | binary | Partially adjusted | -0.2850 | 0.3345 | -0.9407 | 0.3707 | 0.7520 | 0.3904 | 1.4487 | 0.3942 | 0.8942 |
| Mood swings | Regression | binary | Partially adjusted | 0.0849 | 0.0981 | -0.1073 | 0.2771 | 1.0886 | 0.8982 | 1.3193 | 0.3866 | 0.8942 |
| Daytime dozing | Regression | ordered | Partially adjusted | -0.0850 | 0.0996 | -0.2803 | 0.1102 | 0.9185 | 0.7556 | 1.1166 | 0.3935 | 0.8942 |
| Wears glasses/contacts | Progression | binary | Partially adjusted | 0.1435 | 0.1649 | -0.1798 | 0.4668 | 1.1543 | 0.8355 | 1.5948 | 0.3843 | 0.8942 |
| Ever disinterested for a whole week | Progression | binary | Partially adjusted | 0.0759 | 0.0896 | -0.0996 | 0.2515 | 1.0789 | 0.9052 | 1.2859 | 0.3967 | 0.8942 |
| Sleeping too much | Regression | binary | Partially adjusted | -0.1654 | 0.2041 | -0.5654 | 0.2347 | 0.8476 | 0.5681 | 1.2645 | 0.4178 | 0.8942 |
| Fluid intelligence | Regression | ordered | Partially adjusted | -0.0197 | 0.0248 | -0.0682 | 0.0289 | 0.9805 | 0.9341 | 1.0293 | 0.4271 | 0.8942 |
| Miserableness | Regression | binary | Partially adjusted | 0.0811 | 0.0996 | -0.1140 | 0.2763 | 1.0845 | 0.8923 | 1.3182 | 0.4151 | 0.8942 |
| Current smoking | Regression | ordered | Partially adjusted | 0.1434 | 0.1775 | -0.2045 | 0.4914 | 1.1542 | 0.8150 | 1.6347 | 0.4191 | 0.8942 |
| Sensitive stomach | Progression | binary | Partially adjusted | 0.0932 | 0.1188 | -0.1397 | 0.3261 | 1.0977 | 0.8697 | 1.3856 | 0.4327 | 0.8942 |
| Diabetes-related eye disease | Progression | binary | Partially adjusted | 0.4423 | 0.5582 | -0.6519 | 1.5364 | 1.5562 | 0.5211 | 4.6480 | 0.4282 | 0.8942 |
| Toothache | Regression | binary | Partially adjusted | -0.2448 | 0.3550 | -0.9406 | 0.4509 | 0.7828 | 0.3904 | 1.5697 | 0.4904 | 0.8942 |
| Glaucoma | Regression | binary | Partially adjusted | -0.2678 | 0.3807 | -1.0140 | 0.4783 | 0.7650 | 0.3628 | 1.6134 | 0.4817 | 0.8942 |
| Longest manic/irritable episode | Regression | binary | Partially adjusted | 0.1290 | 0.1679 | -0.2000 | 0.4581 | 1.1377 | 0.8187 | 1.5810 | 0.4421 | 0.8942 |
| Happiness | Regression | ordered | Partially adjusted | 0.0486 | 0.0673 | -0.0833 | 0.1805 | 1.0498 | 0.9201 | 1.1979 | 0.4699 | 0.8942 |
| Diabetes diagnosed | Regression | binary | Partially adjusted | 0.1684 | 0.2375 | -0.2972 | 0.6340 | 1.1834 | 0.7429 | 1.8851 | 0.4784 | 0.8942 |
| Worthlessness (depression) | Regression | binary | Partially adjusted | -0.1106 | 0.1486 | -0.4020 | 0.1807 | 0.8953 | 0.6690 | 1.1980 | 0.4566 | 0.8942 |
| Daytime dozing | Progression | ordered | Partially adjusted | -0.0606 | 0.0887 | -0.2344 | 0.1132 | 0.9412 | 0.7910 | 1.1199 | 0.4944 | 0.8942 |
| Miserableness | Progression | binary | Partially adjusted | 0.0670 | 0.0887 | -0.1070 | 0.2409 | 1.0693 | 0.8986 | 1.2724 | 0.4505 | 0.8942 |
| Suffer from nerves | Progression | binary | Partially adjusted | -0.0838 | 0.1145 | -0.3082 | 0.1407 | 0.9196 | 0.7347 | 1.1511 | 0.4645 | 0.8942 |
| Risk taking | Progression | binary | Partially adjusted | -0.0677 | 0.0967 | -0.2572 | 0.1219 | 0.9345 | 0.7732 | 1.1296 | 0.4839 | 0.8942 |
| Happiness with health | Progression | ordered | Partially adjusted | 0.0384 | 0.0527 | -0.0649 | 0.1417 | 1.0391 | 0.9371 | 1.1522 | 0.4667 | 0.8942 |
| Unenthusiasm (2 weeks) | Progression | ordered | Partially adjusted | 0.0658 | 0.0884 | -0.1076 | 0.2391 | 1.0680 | 0.8980 | 1.2701 | 0.4571 | 0.8942 |
| Shortness of breath on level ground | Progression | binary | Partially adjusted | -0.1253 | 0.1759 | -0.4700 | 0.2194 | 0.8823 | 0.6250 | 1.2453 | 0.4762 | 0.8942 |
| Glaucoma | Progression | binary | Partially adjusted | -0.2481 | 0.3342 | -0.9030 | 0.4069 | 0.7803 | 0.4053 | 1.5022 | 0.4579 | 0.8942 |
| Toothache | Progression | binary | Partially adjusted | -0.2140 | 0.3135 | -0.8285 | 0.4005 | 0.8073 | 0.4367 | 1.4925 | 0.4949 | 0.8942 |
| Hip pain 3+ months | Regression | binary | Fully adjusted | -1.1618 | 0.4753 | -2.0934 | -0.2303 | 0.3129 | 0.1233 | 0.7943 | 0.0145 | 0.8942 |
| Walking pace | Progression | ordered | Fully adjusted | -0.2353 | 0.0913 | -0.4142 | -0.0564 | 0.7903 | 0.6609 | 0.9451 | 0.0099 | 0.8942 |
| Strenuous sports frequency | Regression | ordered | Fully adjusted | 0.0894 | 0.1240 | -0.1536 | 0.3324 | 1.0935 | 0.8576 | 1.3943 | 0.4710 | 0.8942 |
| Walking pace | Regression | ordered | Fully adjusted | -0.2348 | 0.1029 | -0.4365 | -0.0331 | 0.7907 | 0.6463 | 0.9674 | 0.0225 | 0.8942 |
| Vigorous activity days/weeks | Regression | ordered | Fully adjusted | -0.0312 | 0.0291 | -0.0883 | 0.0259 | 0.9693 | 0.9155 | 1.0262 | 0.2839 | 0.8942 |
| Moderate activity duration | Regression | ordered | Fully adjusted | -0.0497 | 0.0727 | -0.1923 | 0.0929 | 0.9515 | 0.8251 | 1.0973 | 0.4945 | 0.8942 |
| Duration of walks | Regression | continuous | Fully adjusted | -0.0678 | 0.0564 | -0.1784 | 0.0427 | 0.9344 | 0.8366 | 1.0436 | 0.2290 | 0.8942 |
| Dentures | Regression | binary | Fully adjusted | -0.2398 | 0.2160 | -0.6631 | 0.1835 | 0.7868 | 0.5152 | 1.2014 | 0.2669 | 0.8942 |
| Toothache | Regression | binary | Fully adjusted | -0.3850 | 0.4090 | -1.1867 | 0.4166 | 0.6804 | 0.3052 | 1.5168 | 0.3465 | 0.8942 |
| Bleeding gums | Regression | binary | Fully adjusted | -0.2307 | 0.1768 | -0.5773 | 0.1159 | 0.7940 | 0.5614 | 1.1229 | 0.1921 | 0.8942 |
| Painful gums | Regression | binary | Fully adjusted | -0.5467 | 0.4360 | -1.4013 | 0.3079 | 0.5789 | 0.2463 | 1.3606 | 0.2099 | 0.8942 |
| Mouth ulcers | Regression | binary | Fully adjusted | 0.2056 | 0.1980 | -0.1825 | 0.5937 | 1.2283 | 0.8332 | 1.8106 | 0.2990 | 0.8942 |
| Other serious eye condition | Regression | binary | Fully adjusted | -0.3053 | 0.4170 | -1.1227 | 0.5120 | 0.7369 | 0.3254 | 1.6686 | 0.4641 | 0.8942 |
| Macular degeneration | Regression | binary | Fully adjusted | -0.3592 | 0.4808 | -1.3015 | 0.5832 | 0.6983 | 0.2721 | 1.7918 | 0.4551 | 0.8942 |
| Glaucoma | Regression | binary | Fully adjusted | -0.5952 | 0.4689 | -1.5143 | 0.3238 | 0.5514 | 0.2200 | 1.3824 | 0.2043 | 0.8942 |
| Stress: financial | Regression | binary | Fully adjusted | -0.3839 | 0.3233 | -1.0176 | 0.2498 | 0.6812 | 0.3615 | 1.2838 | 0.2351 | 0.8942 |
| Stress: death of spouse | Regression | binary | Fully adjusted | 1.5640 | 1.0763 | -0.5454 | 3.6735 | 4.7779 | 0.5796 | 39.3882 | 0.1462 | 0.8942 |
| Stress: death of relative | Regression | binary | Fully adjusted | 0.1298 | 0.1574 | -0.1786 | 0.4382 | 1.1386 | 0.8364 | 1.5500 | 0.4094 | 0.8942 |
| Stress: serious illness/assault self | Regression | binary | Fully adjusted | 0.4594 | 0.2553 | -0.0410 | 0.9597 | 1.5831 | 0.9598 | 2.6110 | 0.0719 | 0.8942 |
| Chest pain during activity | Regression | ordered | Fully adjusted | -0.5461 | 0.7144 | -1.9463 | 0.8542 | 0.5792 | 0.1428 | 2.3494 | 0.4446 | 0.8942 |
| Longest manic/irritable episode | Regression | binary | Fully adjusted | 0.2874 | 0.1974 | -0.0994 | 0.6743 | 1.3330 | 0.9054 | 1.9627 | 0.1453 | 0.8942 |
| Longest unenthusiasm/disinterest period | Regression | continuous | Fully adjusted | -0.1153 | 0.1121 | -0.3350 | 0.1044 | 0.8911 | 0.7154 | 1.1100 | 0.3036 | 0.8942 |
| Leg pain on walking | Regression | binary | Fully adjusted | -0.1624 | 0.1518 | -0.4600 | 0.1352 | 0.8501 | 0.6313 | 1.1448 | 0.2848 | 0.8942 |
| Ever highly irritable | Regression | binary | Fully adjusted | -0.1285 | 0.1635 | -0.4490 | 0.1920 | 0.8794 | 0.6383 | 1.2116 | 0.4319 | 0.8942 |
| Ever depressed â‰¥1 week | Regression | binary | Fully adjusted | 0.1245 | 0.1132 | -0.0974 | 0.3464 | 1.1326 | 0.9072 | 1.4140 | 0.2713 | 0.8942 |
| Happiness | Regression | ordered | Fully adjusted | 0.0823 | 0.0797 | -0.0739 | 0.2385 | 1.0858 | 0.9288 | 1.2694 | 0.3016 | 0.8942 |
| Facial pain 3+ months | Regression | binary | Fully adjusted | 2.7335 | 2.0012 | -1.1888 | 6.6558 | 15.3867 | 0.3046 | 777.2759 | 0.1720 | 0.8942 |
| Abdominal pain 3+ months | Regression | binary | Fully adjusted | -0.4539 | 0.5093 | -1.4520 | 0.5443 | 0.6352 | 0.2341 | 1.7234 | 0.3728 | 0.8942 |
| Chest pain when walking | Regression | binary | Fully adjusted | 0.9774 | 0.8540 | -0.6965 | 2.6513 | 2.6574 | 0.4983 | 14.1717 | 0.2525 | 0.8942 |
| Chest pain/discomfort | Regression | binary | Fully adjusted | 0.1624 | 0.1917 | -0.2134 | 0.5382 | 1.1763 | 0.8078 | 1.7128 | 0.3970 | 0.8942 |
| Wheeze in chest | Regression | binary | Fully adjusted | -0.2987 | 0.1606 | -0.6135 | 0.0162 | 0.7418 | 0.5414 | 1.0163 | 0.0630 | 0.8942 |
| Overall health rating | Regression | ordered | Fully adjusted | 0.1545 | 0.0864 | -0.0148 | 0.3238 | 1.1671 | 0.9853 | 1.3824 | 0.0737 | 0.8942 |
| Back pain (3 months) | Regression | ordered | Fully adjusted | -0.0777 | 0.0896 | -0.2533 | 0.0979 | 0.9252 | 0.7763 | 1.1028 | 0.3858 | 0.8942 |
| IBS diagnosis | Regression | binary | Fully adjusted | 0.1360 | 0.1919 | -0.2400 | 0.5120 | 1.1457 | 0.7866 | 1.6686 | 0.4785 | 0.8942 |
| Weight | Regression | continuous | Fully adjusted | 0.1381 | 0.0709 | -0.0009 | 0.2771 | 1.1481 | 0.9991 | 1.3193 | 0.0515 | 0.8942 |
| BMI | Regression | continuous | Fully adjusted | 0.1074 | 0.0608 | -0.0118 | 0.2265 | 1.1133 | 0.9883 | 1.2542 | 0.0774 | 0.8942 |
| Unenthusiasm (2 weeks) | Regression | ordered | Fully adjusted | 0.1668 | 0.1110 | -0.0507 | 0.3843 | 1.1815 | 0.9506 | 1.4686 | 0.1327 | 0.8942 |
| Difficulty stopping worrying | Regression | binary | Fully adjusted | -0.5135 | 0.5098 | -1.5128 | 0.4858 | 0.5984 | 0.2203 | 1.6255 | 0.3139 | 0.8942 |
| Multiple worries | Regression | binary | Fully adjusted | -0.2300 | 0.2811 | -0.7809 | 0.3208 | 0.7945 | 0.4580 | 1.3783 | 0.4131 | 0.8942 |
| Waking too early | Regression | binary | Fully adjusted | 0.2057 | 0.2424 | -0.2693 | 0.6808 | 1.2284 | 0.7639 | 1.9754 | 0.3960 | 0.8942 |
| Sleeping too much | Regression | binary | Fully adjusted | -0.2913 | 0.2538 | -0.7888 | 0.2062 | 0.7473 | 0.4544 | 1.2290 | 0.2511 | 0.8942 |
| Sleep change | Regression | binary | Fully adjusted | 0.5021 | 0.2405 | 0.0307 | 0.9734 | 1.6521 | 1.0312 | 2.6470 | 0.0368 | 0.8942 |
| Depressed mood (2 weeks) | Regression | ordered | Fully adjusted | 0.2308 | 0.1153 | 0.0048 | 0.4567 | 1.2596 | 1.0048 | 1.5789 | 0.0453 | 0.8942 |
| Tiredness (depression) | Regression | binary | Fully adjusted | -0.3864 | 0.2436 | -0.8638 | 0.0910 | 0.6795 | 0.4216 | 1.0953 | 0.1127 | 0.8942 |
| Thoughts of death (depression) | Regression | binary | Fully adjusted | -0.1348 | 0.1779 | -0.4836 | 0.2139 | 0.8739 | 0.6166 | 1.2385 | 0.4486 | 0.8942 |
| Sleep problems (anxiety) | Regression | binary | Fully adjusted | 0.2470 | 0.3064 | -0.3534 | 0.8475 | 1.2802 | 0.7023 | 2.3338 | 0.4201 | 0.8942 |
| Restless (anxiety) | Regression | binary | Fully adjusted | -0.2138 | 0.2483 | -0.7004 | 0.2728 | 0.8075 | 0.4964 | 1.3137 | 0.3892 | 0.8942 |
| Difficulty concentrating (anxiety) | Regression | binary | Fully adjusted | 0.2597 | 0.2727 | -0.2747 | 0.7941 | 1.2966 | 0.7598 | 2.2125 | 0.3408 | 0.8942 |
| Risk taking | Regression | binary | Fully adjusted | -0.2184 | 0.1279 | -0.4691 | 0.0323 | 0.8038 | 0.6255 | 1.0328 | 0.0877 | 0.8942 |
| Guilty feelings | Regression | binary | Fully adjusted | -0.1345 | 0.1280 | -0.3854 | 0.1164 | 0.8742 | 0.6802 | 1.1235 | 0.2935 | 0.8942 |
| Max digits remembered | Regression | ordered | Fully adjusted | -0.1131 | 0.0536 | -0.2182 | -0.0080 | 0.8931 | 0.8040 | 0.9921 | 0.0350 | 0.8942 |
| Neuroticism | Regression | ordered | Fully adjusted | 0.0226 | 0.0190 | -0.0146 | 0.0598 | 1.0228 | 0.9855 | 1.0616 | 0.2336 | 0.8942 |
| Bipolar and major depression status | Regression | unordered | Fully adjusted | 0.2039 | 0.1239 | -0.0389 | 0.4467 | 1.2262 | 0.9619 | 1.5632 | 0.0998 | 0.8942 |
| Probable major depression (single episode) | Regression | binary | Fully adjusted | 0.2379 | 0.3411 | -0.4307 | 0.9065 | 1.2686 | 0.6501 | 2.4758 | 0.4855 | 0.8942 |
| Alcohol drinker status | Regression | binary | Fully adjusted | 0.2747 | 0.2437 | -0.2030 | 0.7524 | 1.3162 | 0.8163 | 2.1221 | 0.2596 | 0.8942 |
| Smoking status | Regression | binary | Fully adjusted | -0.3800 | 0.4246 | -1.2123 | 0.4522 | 0.6838 | 0.2975 | 1.5718 | 0.3708 | 0.8942 |
| Reaction time (mean) | Regression | continuous | Fully adjusted | -0.0913 | 0.0631 | -0.2150 | 0.0324 | 0.9128 | 0.8066 | 1.0329 | 0.1481 | 0.8942 |
| Worry too long after embarrassment | Regression | binary | Fully adjusted | 0.1744 | 0.1144 | -0.0498 | 0.3986 | 1.1905 | 0.9514 | 1.4897 | 0.1274 | 0.8942 |
| Tense/highly strung | Regression | binary | Fully adjusted | 0.1856 | 0.1711 | -0.1497 | 0.5209 | 1.2040 | 0.8610 | 1.6836 | 0.2778 | 0.8942 |
| Worrier/anxious | Regression | binary | Fully adjusted | 0.1983 | 0.1146 | -0.0264 | 0.4230 | 1.2193 | 0.9740 | 1.5265 | 0.0836 | 0.8942 |
| Fed-up feelings | Regression | binary | Fully adjusted | 0.1829 | 0.1213 | -0.0548 | 0.4207 | 1.2007 | 0.9466 | 1.5230 | 0.1316 | 0.8942 |
| Sensitivity/hurt feelings | Regression | binary | Fully adjusted | 0.1239 | 0.1150 | -0.1014 | 0.3493 | 1.1319 | 0.9036 | 1.4181 | 0.2811 | 0.8942 |
| Irritability | Regression | binary | Fully adjusted | 0.1072 | 0.1279 | -0.1435 | 0.3580 | 1.1132 | 0.8663 | 1.4304 | 0.4019 | 0.8942 |
| Miserableness | Regression | binary | Fully adjusted | 0.1631 | 0.1188 | -0.0698 | 0.3960 | 1.1772 | 0.9326 | 1.4859 | 0.1699 | 0.8942 |
| Mood swings | Regression | binary | Fully adjusted | 0.1170 | 0.1171 | -0.1126 | 0.3467 | 1.1242 | 0.8935 | 1.4143 | 0.3177 | 0.8942 |
| Light DIY duration | Regression | ordered | Fully adjusted | -0.0546 | 0.0502 | -0.1530 | 0.0438 | 0.9468 | 0.8581 | 1.0448 | 0.2767 | 0.8942 |
| Sleeplessness/insomnia | Regression | ordered | Fully adjusted | 0.0604 | 0.0765 | -0.0896 | 0.2103 | 1.0622 | 0.9143 | 1.2341 | 0.4300 | 0.8942 |
| Strenuous sports duration | Progression | ordered | Fully adjusted | -0.0979 | 0.1215 | -0.3361 | 0.1402 | 0.9067 | 0.7146 | 1.1505 | 0.4202 | 0.8942 |
| Light DIY duration | Progression | ordered | Fully adjusted | -0.0569 | 0.0440 | -0.1431 | 0.0293 | 0.9447 | 0.8667 | 1.0297 | 0.1958 | 0.8942 |
| Current smoking | Progression | ordered | Fully adjusted | 0.1697 | 0.1805 | -0.1840 | 0.5234 | 1.1849 | 0.8319 | 1.6877 | 0.3471 | 0.8942 |
| Alcohol intake frequency | Progression | ordered | Fully adjusted | 0.0430 | 0.0366 | -0.0287 | 0.1147 | 1.0439 | 0.9717 | 1.1215 | 0.2396 | 0.8942 |
| Miserableness | Progression | binary | Fully adjusted | 0.0802 | 0.1049 | -0.1253 | 0.2857 | 1.0835 | 0.8822 | 1.3307 | 0.4443 | 0.8942 |
| Sensitivity/hurt feelings | Progression | binary | Fully adjusted | 0.1744 | 0.1011 | -0.0239 | 0.3726 | 1.1905 | 0.9764 | 1.4516 | 0.0847 | 0.8942 |
| Nervous feelings | Progression | binary | Fully adjusted | 0.1060 | 0.1293 | -0.1474 | 0.3593 | 1.1118 | 0.8629 | 1.4324 | 0.4124 | 0.8942 |
| Worrier/anxious | Progression | binary | Fully adjusted | 0.0925 | 0.1011 | -0.1057 | 0.2907 | 1.0969 | 0.8997 | 1.3374 | 0.3603 | 0.8942 |
| Tense/highly strung | Progression | binary | Fully adjusted | 0.2079 | 0.1514 | -0.0889 | 0.5046 | 1.2310 | 0.9150 | 1.6563 | 0.1698 | 0.8942 |
| Bipolar and major depression status | Progression | unordered | Fully adjusted | 0.0843 | 0.1090 | -0.1293 | 0.2978 | 1.0879 | 0.8787 | 1.3469 | 0.4393 | 0.8942 |
| Loneliness/isolation | Progression | binary | Fully adjusted | -0.1550 | 0.1420 | -0.4333 | 0.1233 | 0.8564 | 0.6483 | 1.1312 | 0.2749 | 0.8942 |
| Max digits remembered | Progression | ordered | Fully adjusted | -0.0322 | 0.0468 | -0.1238 | 0.0595 | 0.9684 | 0.8835 | 1.0613 | 0.4918 | 0.8942 |
| Risk taking | Progression | binary | Fully adjusted | -0.1537 | 0.1125 | -0.3742 | 0.0667 | 0.8575 | 0.6878 | 1.0690 | 0.1717 | 0.8942 |
| Thoughts of death (depression) | Progression | binary | Fully adjusted | -0.1246 | 0.1579 | -0.4341 | 0.1850 | 0.8829 | 0.6479 | 1.2032 | 0.4303 | 0.8942 |
| Tiredness (depression) | Progression | binary | Fully adjusted | -0.2249 | 0.2237 | -0.6633 | 0.2134 | 0.7986 | 0.5151 | 1.2379 | 0.3145 | 0.8942 |
| Worthlessness (depression) | Progression | binary | Fully adjusted | -0.1281 | 0.1591 | -0.4399 | 0.1838 | 0.8798 | 0.6441 | 1.2018 | 0.4209 | 0.8942 |
| Depressed mood (2 weeks) | Progression | ordered | Fully adjusted | 0.1767 | 0.1042 | -0.0275 | 0.3809 | 1.1932 | 0.9728 | 1.4636 | 0.0899 | 0.8942 |
| Sleep change | Progression | binary | Fully adjusted | 0.2272 | 0.2047 | -0.1741 | 0.6285 | 1.2551 | 0.8402 | 1.8748 | 0.2671 | 0.8942 |
| Weight change during worst depression episode | Progression | unordered | Fully adjusted | -0.0887 | 0.0843 | -0.2539 | 0.0766 | 0.9152 | 0.7758 | 1.0796 | 0.2929 | 0.8942 |
| Multiple worries | Progression | binary | Fully adjusted | -0.1929 | 0.2530 | -0.6888 | 0.3031 | 0.8246 | 0.5022 | 1.3540 | 0.4459 | 0.8942 |
| Tiredness/lethargy (2 weeks) | Progression | ordered | Fully adjusted | -0.0457 | 0.0628 | -0.1688 | 0.0774 | 0.9553 | 0.8447 | 1.0804 | 0.4665 | 0.8942 |
| BMI | Progression | continuous | Fully adjusted | 0.0940 | 0.0530 | -0.0098 | 0.1978 | 1.0985 | 0.9902 | 1.2187 | 0.0760 | 0.8942 |
| Weight | Progression | continuous | Fully adjusted | 0.0589 | 0.0620 | -0.0626 | 0.1805 | 1.0607 | 0.9393 | 1.1978 | 0.3420 | 0.8942 |
| Back pain (3 months) | Progression | ordered | Fully adjusted | -0.0873 | 0.0791 | -0.2424 | 0.0678 | 0.9164 | 0.7847 | 1.0701 | 0.2699 | 0.8942 |
| Sensitive stomach | Progression | binary | Fully adjusted | 0.1064 | 0.1405 | -0.1689 | 0.3817 | 1.1122 | 0.8446 | 1.4647 | 0.4489 | 0.8942 |
| Family history of IBS | Progression | binary | Fully adjusted | 0.1074 | 0.1519 | -0.1904 | 0.4051 | 1.1133 | 0.8267 | 1.4994 | 0.4797 | 0.8942 |
| Able to confide | Regression | ordered | Fully adjusted | -0.0229 | 0.0311 | -0.0839 | 0.0381 | 0.9774 | 0.9196 | 1.0389 | 0.4628 | 0.8942 |
| Overall health rating | Progression | ordered | Fully adjusted | 0.0839 | 0.0769 | -0.0667 | 0.2346 | 1.0876 | 0.9354 | 1.2644 | 0.2749 | 0.8942 |
| Wears glasses/contacts | Progression | binary | Fully adjusted | 0.3622 | 0.1891 | -0.0084 | 0.7329 | 1.4366 | 0.9916 | 2.0811 | 0.0554 | 0.8942 |
| Wheeze in chest | Progression | binary | Fully adjusted | -0.1148 | 0.1365 | -0.3823 | 0.1527 | 0.8916 | 0.6823 | 1.1650 | 0.4003 | 0.8942 |
| Diabetes diagnosed | Progression | binary | Fully adjusted | 0.3807 | 0.2727 | -0.1538 | 0.9153 | 1.4633 | 0.8574 | 2.4975 | 0.1627 | 0.8942 |
| Chest pain when walking | Progression | binary | Fully adjusted | 0.5922 | 0.8128 | -1.0009 | 2.1852 | 1.8079 | 0.3676 | 8.8926 | 0.4663 | 0.8942 |
| Abdominal pain 3+ months | Progression | binary | Fully adjusted | -0.5903 | 0.4546 | -1.4813 | 0.3006 | 0.5542 | 0.2274 | 1.3507 | 0.1941 | 0.8942 |
| Headaches 3+ months | Progression | binary | Fully adjusted | 0.4895 | 0.2790 | -0.0573 | 1.0364 | 1.6316 | 0.9443 | 2.8191 | 0.0793 | 0.8942 |
| Happiness | Progression | ordered | Fully adjusted | 0.0689 | 0.0705 | -0.0692 | 0.2070 | 1.0713 | 0.9331 | 1.2300 | 0.3282 | 0.8942 |
| Ever depressed for a whole week | Progression | binary | Fully adjusted | 0.0784 | 0.0996 | -0.1168 | 0.2736 | 1.0815 | 0.8897 | 1.3146 | 0.4313 | 0.8942 |
| Longest manic/irritable episode | Progression | binary | Fully adjusted | 0.2345 | 0.1790 | -0.1163 | 0.5853 | 1.2642 | 0.8902 | 1.7954 | 0.1901 | 0.8942 |
| Chest pain outside activity | Progression | ordered | Fully adjusted | 0.5076 | 0.4282 | -0.3316 | 1.3467 | 1.6612 | 0.7178 | 3.8449 | 0.2358 | 0.8942 |
| Stress: serious illness/assault self | Progression | binary | Fully adjusted | 0.1867 | 0.2383 | -0.2804 | 0.6538 | 1.2053 | 0.7555 | 1.9229 | 0.4333 | 0.8942 |
| Stress: serious illness/assault relative | Regression | binary | Fully adjusted | -0.1881 | 0.1598 | -0.5013 | 0.1251 | 0.8285 | 0.6058 | 1.1333 | 0.2392 | 0.8942 |
| Stress: serious illness/assault relative | Progression | binary | Fully adjusted | -0.1868 | 0.1378 | -0.4569 | 0.0834 | 0.8296 | 0.6332 | 1.0870 | 0.1754 | 0.8942 |
| Stress: death of relative | Progression | binary | Fully adjusted | 0.1940 | 0.1394 | -0.0792 | 0.4671 | 1.2141 | 0.9239 | 1.5954 | 0.1640 | 0.8942 |
| Stress: death of spouse | Progression | binary | Fully adjusted | 0.7494 | 1.0686 | -1.3450 | 2.8438 | 2.1156 | 0.2605 | 17.1802 | 0.4831 | 0.8942 |
| Diabetes-related eye disease | Progression | binary | Fully adjusted | 1.5356 | 1.0365 | -0.4960 | 3.5673 | 4.6443 | 0.6090 | 35.4192 | 0.1385 | 0.8942 |
| Macular degeneration | Progression | binary | Fully adjusted | -0.3083 | 0.4130 | -1.1177 | 0.5011 | 0.7347 | 0.3270 | 1.6506 | 0.4554 | 0.8942 |
| Other serious eye condition | Progression | binary | Fully adjusted | -0.6688 | 0.3824 | -1.4183 | 0.0808 | 0.5123 | 0.2421 | 1.0841 | 0.0803 | 0.8942 |
| Bleeding gums | Progression | binary | Fully adjusted | -0.1662 | 0.1515 | -0.4632 | 0.1307 | 0.8469 | 0.6293 | 1.1397 | 0.2726 | 0.8942 |
| Loose teeth | Progression | binary | Fully adjusted | 0.3184 | 0.4306 | -0.5256 | 1.1624 | 1.3749 | 0.5912 | 3.1974 | 0.4597 | 0.8942 |
| Toothache | Progression | binary | Fully adjusted | -0.3678 | 0.3557 | -1.0651 | 0.3294 | 0.6922 | 0.3447 | 1.3902 | 0.3012 | 0.8942 |
| Dentures | Progression | binary | Fully adjusted | -0.1354 | 0.1888 | -0.5054 | 0.2346 | 0.8734 | 0.6033 | 1.2644 | 0.4732 | 0.8942 |
| High blood pressure | Progression | binary | Fully adjusted | -0.5277 | 0.4727 | -1.4541 | 0.3988 | 0.5900 | 0.2336 | 1.4900 | 0.2643 | 0.8942 |
| Duration of walks | Progression | continuous | Fully adjusted | -0.0631 | 0.0495 | -0.1600 | 0.0339 | 0.9389 | 0.8522 | 1.0344 | 0.2022 | 0.8942 |
| Moderate activity days/weeks | Progression | ordered | Fully adjusted | -0.0357 | 0.0228 | -0.0803 | 0.0089 | 0.9649 | 0.9228 | 1.0089 | 0.1162 | 0.8942 |
| Moderate activity duration | Progression | ordered | Fully adjusted | -0.0457 | 0.0639 | -0.1710 | 0.0796 | 0.9553 | 0.8428 | 1.0829 | 0.4748 | 0.8942 |
| Vigorous activity days/weeks | Progression | ordered | Fully adjusted | -0.0290 | 0.0257 | -0.0794 | 0.0214 | 0.9714 | 0.9237 | 1.0216 | 0.2590 | 0.8942 |
| Duration of vigorous activity | Progression | continuous | Fully adjusted | -0.0411 | 0.0587 | -0.1561 | 0.0739 | 0.9597 | 0.8555 | 1.0767 | 0.4837 | 0.8942 |
| Strenuous sports frequency | Progression | ordered | Fully adjusted | 0.1730 | 0.1091 | -0.0409 | 0.3869 | 1.1889 | 0.9600 | 1.4724 | 0.1128 | 0.8942 |
| Longest worried/anxious period | Progression | continuous | Fully adjusted | 0.0898 | 0.1320 | -0.1689 | 0.3485 | 1.0939 | 0.8446 | 1.4169 | 0.4963 | 0.8942 |
| Sleep problems (anxiety) | Progression | binary | Fully adjusted | 0.1803 | 0.2693 | -0.3475 | 0.7082 | 1.1976 | 0.7065 | 2.0303 | 0.5031 | 0.9028 |
| Ever disinterested for a week | Regression | binary | Partially adjusted | 0.0661 | 0.1005 | -0.1309 | 0.2632 | 1.0684 | 0.8773 | 1.3011 | 0.5106 | 0.9065 |
| General happiness | Progression | ordered | Partially adjusted | -0.0407 | 0.0622 | -0.1626 | 0.0812 | 0.9601 | 0.8500 | 1.0845 | 0.5127 | 0.9065 |
| Happiness with health | Regression | ordered | Fully adjusted | 0.0465 | 0.0710 | -0.0927 | 0.1856 | 1.0475 | 0.9115 | 1.2040 | 0.5130 | 0.9065 |
| Worthlessness (depression) | Regression | binary | Fully adjusted | -0.1178 | 0.1798 | -0.4703 | 0.2347 | 0.8889 | 0.6248 | 1.2645 | 0.5125 | 0.9065 |
| Multiple worries | Progression | binary | Partially adjusted | -0.1359 | 0.2088 | -0.5451 | 0.2733 | 0.8729 | 0.5798 | 1.3143 | 0.5151 | 0.9068 |
| Cataract | Regression | binary | Partially adjusted | 0.1208 | 0.1943 | -0.2600 | 0.5015 | 1.1284 | 0.7711 | 1.6512 | 0.5341 | 0.9076 |
| Nervous feelings | Regression | binary | Partially adjusted | 0.0784 | 0.1239 | -0.1644 | 0.3213 | 1.0816 | 0.8484 | 1.3789 | 0.5267 | 0.9076 |
| Strenuous sports duration | Regression | ordered | Partially adjusted | 0.0690 | 0.1098 | -0.1462 | 0.2842 | 1.0715 | 0.8640 | 1.3287 | 0.5296 | 0.9076 |
| Able to confide | Regression | ordered | Partially adjusted | -0.0162 | 0.0261 | -0.0673 | 0.0350 | 0.9840 | 0.9349 | 1.0356 | 0.5352 | 0.9076 |
| Facial pain 3+ months: No | Progression | binary | Partially adjusted | 0.5562 | 0.8858 | -1.1800 | 2.2925 | 1.7441 | 0.3073 | 9.8997 | 0.5301 | 0.9076 |
| Vision loss from injury | Progression | binary | Partially adjusted | 0.6968 | 1.0908 | -1.4412 | 2.8349 | 2.0074 | 0.2366 | 17.0284 | 0.5229 | 0.9076 |
| Suffer from nerves | Regression | binary | Fully adjusted | -0.0981 | 0.1542 | -0.4002 | 0.2041 | 0.9066 | 0.6702 | 1.2264 | 0.5247 | 0.9076 |
| Fluid intelligence | Progression | ordered | Fully adjusted | -0.0165 | 0.0262 | -0.0678 | 0.0348 | 0.9836 | 0.9344 | 1.0354 | 0.5277 | 0.9076 |
| Suffer from nerves | Progression | binary | Fully adjusted | -0.0854 | 0.1349 | -0.3499 | 0.1791 | 0.9182 | 0.7048 | 1.1962 | 0.5270 | 0.9076 |
| Depression episodes | Progression | ordered | Fully adjusted | -0.0546 | 0.0873 | -0.2256 | 0.1165 | 0.9469 | 0.7980 | 1.1235 | 0.5317 | 0.9076 |
| Longest unenthusiasm/disinterest period | Progression | continuous | Fully adjusted | 0.0605 | 0.0983 | -0.1322 | 0.2532 | 1.0623 | 0.8762 | 1.2881 | 0.5384 | 0.9098 |
| Back pain (3 months) | Regression | ordered | Partially adjusted | -0.0437 | 0.0749 | -0.1906 | 0.1032 | 0.9572 | 0.8265 | 1.1087 | 0.5597 | 0.9122 |
| Restless (anxiety) | Regression | binary | Partially adjusted | -0.1275 | 0.2100 | -0.5390 | 0.2840 | 0.8803 | 0.5833 | 1.3285 | 0.5437 | 0.9122 |
| Snoring | Progression | binary | Partially adjusted | -0.0544 | 0.0910 | -0.2327 | 0.1239 | 0.9471 | 0.7924 | 1.1319 | 0.5500 | 0.9122 |
| Diabetes diagnosed | Progression | binary | Partially adjusted | 0.1272 | 0.2176 | -0.2993 | 0.5538 | 1.1357 | 0.7413 | 1.7399 | 0.5588 | 0.9122 |
| Hearing aid user | Progression | binary | Partially adjusted | 0.1155 | 0.2000 | -0.2764 | 0.5074 | 1.1224 | 0.7585 | 1.6610 | 0.5635 | 0.9122 |
| Shortness of breath on level ground | Regression | binary | Partially adjusted | -0.1190 | 0.1993 | -0.5097 | 0.2716 | 0.8878 | 0.6007 | 1.3120 | 0.5503 | 0.9122 |
| Loose teeth | Progression | binary | Partially adjusted | 0.2182 | 0.3619 | -0.4910 | 0.9274 | 1.2438 | 0.6120 | 2.5280 | 0.5465 | 0.9122 |
| Hearing aid user | Regression | binary | Fully adjusted | -0.1583 | 0.2734 | -0.6942 | 0.3775 | 0.8536 | 0.4995 | 1.4586 | 0.5625 | 0.9122 |
| Other eye problems | Regression | binary | Fully adjusted | -0.0904 | 0.1549 | -0.3940 | 0.2132 | 0.9135 | 0.6743 | 1.2376 | 0.5593 | 0.9122 |
| Weight change during worst depression episode | Regression | unordered | Fully adjusted | -0.0575 | 0.0950 | -0.2437 | 0.1287 | 0.9442 | 0.7837 | 1.1374 | 0.5452 | 0.9122 |
| Trouble falling asleep | Progression | binary | Fully adjusted | -0.1262 | 0.2156 | -0.5487 | 0.2963 | 0.8814 | 0.5777 | 1.3449 | 0.5582 | 0.9122 |
| Chest pain during activity | Progression | ordered | Fully adjusted | -0.2803 | 0.4800 | -1.2211 | 0.6605 | 0.7556 | 0.2949 | 1.9358 | 0.5593 | 0.9122 |
| Longest manic/irritable episode | Progression | binary | Partially adjusted | 0.0866 | 0.1508 | -0.2090 | 0.3821 | 1.0904 | 0.8114 | 1.4654 | 0.5660 | 0.9131 |
| Stress: divorce/separation | Regression | binary | Fully adjusted | -0.2846 | 0.5021 | -1.2686 | 0.6995 | 0.7523 | 0.2812 | 2.0127 | 0.5709 | 0.9177 |
| Dentures | Regression | binary | Partially adjusted | -0.0998 | 0.1809 | -0.4542 | 0.2547 | 0.9050 | 0.6349 | 1.2901 | 0.5812 | 0.9197 |
| Weight change during worst depression episode | Regression | unordered | Partially adjusted | -0.0437 | 0.0788 | -0.1981 | 0.1108 | 0.9573 | 0.8203 | 1.1171 | 0.5794 | 0.9197 |
| Fed-up feelings | Progression | binary | Partially adjusted | 0.0511 | 0.0911 | -0.1275 | 0.2297 | 1.0525 | 0.8803 | 1.2582 | 0.5747 | 0.9197 |
| Able to confide | Progression | ordered | Partially adjusted | -0.0129 | 0.0235 | -0.0589 | 0.0331 | 0.9872 | 0.9428 | 1.0337 | 0.5829 | 0.9197 |
| Trouble falling asleep | Regression | binary | Fully adjusted | -0.1325 | 0.2419 | -0.6066 | 0.3416 | 0.8759 | 0.5452 | 1.4073 | 0.5840 | 0.9197 |
| Chest pain/discomfort | Progression | binary | Fully adjusted | 0.0950 | 0.1714 | -0.2409 | 0.4308 | 1.0996 | 0.7859 | 1.5385 | 0.5794 | 0.9197 |
| Multiple worries | Regression | binary | Partially adjusted | -0.1227 | 0.2334 | -0.5802 | 0.3349 | 0.8846 | 0.5598 | 1.3978 | 0.5993 | 0.9218 |
| Difficulty concentrating (anxiety) | Regression | binary | Partially adjusted | 0.1192 | 0.2261 | -0.3239 | 0.5623 | 1.1266 | 0.7233 | 1.7547 | 0.5981 | 0.9218 |
| Fed-up feelings | Progression | binary | Fully adjusted | 0.0575 | 0.1077 | -0.1536 | 0.2686 | 1.0592 | 0.8576 | 1.3081 | 0.5934 | 0.9218 |
| Difficulty concentrating (anxiety) | Progression | binary | Fully adjusted | 0.1287 | 0.2378 | -0.3374 | 0.5947 | 1.1373 | 0.7136 | 1.8125 | 0.5884 | 0.9218 |
| Any vascular/heart problem | Progression | binary | Fully adjusted | 0.2471 | 0.4624 | -0.6593 | 1.1534 | 1.2802 | 0.5172 | 3.1690 | 0.5932 | 0.9218 |
| Puzzle score | Regression | ordered | Fully adjusted | -0.0161 | 0.0303 | -0.0755 | 0.0433 | 0.9840 | 0.9273 | 1.0443 | 0.5955 | 0.9218 |
| Fracture from fall | Progression | binary | Fully adjusted | -0.1829 | 0.3464 | -0.8619 | 0.4961 | 0.8329 | 0.4224 | 1.6423 | 0.5976 | 0.9218 |
| Probable major depression (single episode) | Progression | binary | Fully adjusted | -0.1659 | 0.3211 | -0.7953 | 0.4634 | 0.8471 | 0.4515 | 1.5895 | 0.6053 | 0.9281 |
| Other eye problems | Progression | binary | Partially adjusted | -0.0577 | 0.1127 | -0.2786 | 0.1633 | 0.9440 | 0.7568 | 1.1774 | 0.6091 | 0.9307 |
| Happiness with health | Regression | ordered | Partially adjusted | 0.0298 | 0.0591 | -0.0860 | 0.1457 | 1.0303 | 0.9176 | 1.1568 | 0.6135 | 0.9320 |
| Probable major depression (single episode) | Regression | binary | Partially adjusted | -0.1475 | 0.2926 | -0.7210 | 0.4259 | 0.8628 | 0.4863 | 1.5309 | 0.6140 | 0.9320 |
| Happiness with health | Progression | ordered | Fully adjusted | 0.0315 | 0.0628 | -0.0916 | 0.1547 | 1.0320 | 0.9124 | 1.1673 | 0.6160 | 0.9320 |
| Guilty feelings | Progression | binary | Partially adjusted | 0.0465 | 0.0947 | -0.1391 | 0.2320 | 1.0476 | 0.8702 | 1.2612 | 0.6235 | 0.9344 |
| IBS diagnosis | Progression | binary | Partially adjusted | 0.0722 | 0.1475 | -0.2169 | 0.3613 | 1.0748 | 0.8050 | 1.4351 | 0.6246 | 0.9344 |
| Duration of vigorous activity | Regression | continuous | Fully adjusted | -0.0330 | 0.0669 | -0.1640 | 0.0981 | 0.9676 | 0.8487 | 1.1030 | 0.6219 | 0.9344 |
| Hearing aid user | Progression | binary | Fully adjusted | 0.1155 | 0.2367 | -0.3485 | 0.5795 | 1.1224 | 0.7057 | 1.7852 | 0.6256 | 0.9344 |
| Guilty feelings | Regression | binary | Partially adjusted | -0.0521 | 0.1075 | -0.2629 | 0.1586 | 0.9492 | 0.7688 | 1.1719 | 0.6278 | 0.9346 |
| Headaches 3+ months | Regression | binary | Fully adjusted | 0.1547 | 0.3226 | -0.4777 | 0.7870 | 1.1673 | 0.6202 | 2.1969 | 0.6316 | 0.9373 |
| Chest pain during activity | Progression | ordered | Partially adjusted | -0.1953 | 0.4113 | -1.0015 | 0.6109 | 0.8226 | 0.3673 | 1.8421 | 0.6350 | 0.9381 |
| Sleeplessness/insomnia | Regression | ordered | Partially adjusted | 0.0287 | 0.0640 | -0.0968 | 0.1541 | 1.0291 | 0.9077 | 1.1666 | 0.6542 | 0.9381 |
| Strenuous sports duration | Progression | ordered | Partially adjusted | -0.0456 | 0.1001 | -0.2418 | 0.1507 | 0.9555 | 0.7852 | 1.1627 | 0.6492 | 0.9381 |
| Loneliness/isolation | Progression | binary | Partially adjusted | -0.0560 | 0.1213 | -0.2936 | 0.1817 | 0.9456 | 0.7455 | 1.1993 | 0.6445 | 0.9381 |
| Difficulty concentrating (anxiety) | Progression | binary | Partially adjusted | 0.0922 | 0.2001 | -0.3001 | 0.4845 | 1.0966 | 0.7408 | 1.6233 | 0.6450 | 0.9381 |
| Depression episodes | Regression | ordered | Partially adjusted | 0.0366 | 0.0831 | -0.1263 | 0.1995 | 1.0373 | 0.8813 | 1.2208 | 0.6597 | 0.9381 |
| Cataract | Regression | binary | Fully adjusted | -0.0962 | 0.2186 | -0.5247 | 0.3323 | 0.9083 | 0.5917 | 1.3941 | 0.6599 | 0.9381 |
| Diabetes diagnosed | Regression | binary | Fully adjusted | 0.1379 | 0.3073 | -0.4645 | 0.7402 | 1.1478 | 0.6285 | 2.0964 | 0.6537 | 0.9381 |
| Long-standing illness/disability | Regression | binary | Fully adjusted | 0.0626 | 0.1326 | -0.1974 | 0.3226 | 1.0646 | 0.8209 | 1.3808 | 0.6368 | 0.9381 |
| Sensitive stomach | Regression | binary | Fully adjusted | 0.0737 | 0.1590 | -0.2379 | 0.3852 | 1.0764 | 0.7883 | 1.4700 | 0.6431 | 0.9381 |
| Nervous feelings | Regression | binary | Fully adjusted | 0.0668 | 0.1466 | -0.2206 | 0.3542 | 1.0691 | 0.8020 | 1.4250 | 0.6488 | 0.9381 |
| Neuroticism | Progression | ordered | Fully adjusted | 0.0075 | 0.0169 | -0.0256 | 0.0407 | 1.0076 | 0.9747 | 1.0415 | 0.6552 | 0.9381 |
| Able to confide | Progression | ordered | Fully adjusted | -0.0130 | 0.0279 | -0.0676 | 0.0416 | 0.9871 | 0.9346 | 1.0424 | 0.6402 | 0.9381 |
| Stress: financial | Progression | binary | Fully adjusted | 0.1129 | 0.2570 | -0.3909 | 0.6166 | 1.1195 | 0.6765 | 1.8527 | 0.6605 | 0.9381 |
| Loneliness/isolation | Regression | binary | Partially adjusted | 0.0572 | 0.1345 | -0.2064 | 0.3208 | 1.0589 | 0.8135 | 1.3782 | 0.6707 | 0.9435 |
| Alcohol intake frequency | Regression | ordered | Partially adjusted | 0.0148 | 0.0345 | -0.0529 | 0.0825 | 1.0149 | 0.9484 | 1.0859 | 0.6693 | 0.9435 |
| Mood swings | Progression | binary | Partially adjusted | 0.0371 | 0.0877 | -0.1349 | 0.2091 | 1.0378 | 0.8738 | 1.2325 | 0.6724 | 0.9435 |
| Any vascular/heart problem | Progression | binary | Partially adjusted | 0.1953 | 0.4552 | -0.6969 | 1.0876 | 1.2157 | 0.4981 | 2.9670 | 0.6679 | 0.9435 |
| Moderate activity days/weeks | Progression | ordered | Partially adjusted | -0.0079 | 0.0192 | -0.0456 | 0.0298 | 0.9921 | 0.9554 | 1.0303 | 0.6810 | 0.9497 |
| Cataract | Progression | binary | Fully adjusted | 0.0792 | 0.1917 | -0.2965 | 0.4549 | 1.0824 | 0.7434 | 1.5760 | 0.6794 | 0.9497 |
| Macular degeneration | Regression | binary | Partially adjusted | -0.1738 | 0.4341 | -1.0248 | 0.6771 | 0.8404 | 0.3589 | 1.9681 | 0.6889 | 0.9521 |
| Back pain 3+ months | Progression | binary | Partially adjusted | -0.0781 | 0.1937 | -0.4578 | 0.3017 | 0.9249 | 0.6327 | 1.3521 | 0.6870 | 0.9521 |
| Moderate activity duration | Progression | ordered | Partially adjusted | -0.0219 | 0.0541 | -0.1280 | 0.0842 | 0.9783 | 0.8798 | 1.0879 | 0.6858 | 0.9521 |
| Moderate activity days/weeks | Regression | ordered | Partially adjusted | 0.0079 | 0.0215 | -0.0344 | 0.0501 | 1.0079 | 0.9662 | 1.0514 | 0.7156 | 0.9567 |
| High blood pressure | Regression | binary | Partially adjusted | -0.2087 | 0.7192 | -1.6183 | 1.2009 | 0.8117 | 0.1982 | 3.3232 | 0.7717 | 0.9567 |
| Chest pain/discomfort | Regression | binary | Partially adjusted | -0.0447 | 0.1606 | -0.3596 | 0.2701 | 0.9562 | 0.6980 | 1.3101 | 0.7806 | 0.9567 |
| Falls in last year | Regression | ordered | Partially adjusted | -0.0346 | 0.0920 | -0.2149 | 0.1458 | 0.9660 | 0.8066 | 1.1570 | 0.7072 | 0.9567 |
| Sensitive stomach | Regression | binary | Partially adjusted | 0.0487 | 0.1329 | -0.2118 | 0.3092 | 1.0499 | 0.8092 | 1.3624 | 0.7139 | 0.9567 |
| Tenseness/restlessness (2 weeks) | Regression | ordered | Partially adjusted | -0.0295 | 0.0887 | -0.2033 | 0.1443 | 0.9709 | 0.8160 | 1.1553 | 0.7394 | 0.9567 |
| Ever worried more than others | Regression | binary | Partially adjusted | 0.0377 | 0.1331 | -0.2232 | 0.2987 | 1.0384 | 0.7999 | 1.3481 | 0.7770 | 0.9567 |
| Bipolar and major depression status | Regression | unordered | Partially adjusted | 0.0349 | 0.1060 | -0.1728 | 0.2427 | 1.0356 | 0.8413 | 1.2746 | 0.7416 | 0.9567 |
| Alcohol intake frequency | Progression | ordered | Partially adjusted | 0.0085 | 0.0309 | -0.0521 | 0.0691 | 1.0085 | 0.9493 | 1.0715 | 0.7832 | 0.9567 |
| Bipolar and major depression status | Progression | unordered | Partially adjusted | 0.0278 | 0.0921 | -0.1528 | 0.2083 | 1.0282 | 0.8583 | 1.2316 | 0.7631 | 0.9567 |
| Ever worried more than others | Progression | binary | Partially adjusted | 0.0382 | 0.1184 | -0.1939 | 0.2702 | 1.0389 | 0.8238 | 1.3103 | 0.7471 | 0.9567 |
| Tiredness (depression) | Progression | binary | Partially adjusted | -0.0631 | 0.1844 | -0.4246 | 0.2984 | 0.9388 | 0.6540 | 1.3477 | 0.7322 | 0.9567 |
| Worthlessness (depression) | Progression | binary | Partially adjusted | -0.0465 | 0.1333 | -0.3078 | 0.2147 | 0.9545 | 0.7351 | 1.2395 | 0.7270 | 0.9567 |
| Sleep change | Progression | binary | Partially adjusted | 0.0637 | 0.1793 | -0.2878 | 0.4151 | 1.0658 | 0.7499 | 1.5146 | 0.7224 | 0.9567 |
| Trouble falling asleep | Progression | binary | Partially adjusted | -0.0550 | 0.1729 | -0.3938 | 0.2838 | 0.9465 | 0.6745 | 1.3282 | 0.7505 | 0.9567 |
| Sleeping too much | Progression | binary | Partially adjusted | -0.0648 | 0.1801 | -0.4177 | 0.2881 | 0.9372 | 0.6585 | 1.3339 | 0.7189 | 0.9567 |
| Waking too early | Progression | binary | Partially adjusted | -0.0538 | 0.1755 | -0.3978 | 0.2902 | 0.9476 | 0.6718 | 1.3367 | 0.7592 | 0.9567 |
| Tenseness/restlessness (2 weeks) | Progression | ordered | Partially adjusted | -0.0293 | 0.0788 | -0.1838 | 0.1252 | 0.9711 | 0.8321 | 1.1334 | 0.7103 | 0.9567 |
| Family history of IBS | Progression | binary | Partially adjusted | 0.0366 | 0.1308 | -0.2198 | 0.2929 | 1.0372 | 0.8027 | 1.3403 | 0.7798 | 0.9567 |
| Chest pain when walking | Progression | binary | Partially adjusted | 0.1738 | 0.5409 | -0.8863 | 1.2340 | 1.1898 | 0.4122 | 3.4348 | 0.7479 | 0.9567 |
| Stress: divorce/separation | Progression | binary | Partially adjusted | 0.1135 | 0.3748 | -0.6210 | 0.8481 | 1.1202 | 0.5374 | 2.3351 | 0.7620 | 0.9567 |
| Dentures | Progression | binary | Partially adjusted | -0.0460 | 0.1608 | -0.3613 | 0.2692 | 0.9550 | 0.6968 | 1.3089 | 0.7747 | 0.9567 |
| Moderate activity duration | Regression | ordered | Partially adjusted | -0.0165 | 0.0609 | -0.1358 | 0.1029 | 0.9837 | 0.8730 | 1.1084 | 0.7867 | 0.9567 |
| Fracture from fall | Regression | binary | Partially adjusted | -0.0866 | 0.3247 | -0.7231 | 0.5498 | 0.9170 | 0.4853 | 1.7329 | 0.7896 | 0.9567 |
| Moderate activity days/weeks | Regression | ordered | Fully adjusted | -0.0095 | 0.0257 | -0.0600 | 0.0410 | 0.9906 | 0.9418 | 1.0418 | 0.7125 | 0.9567 |
| High blood pressure | Regression | binary | Fully adjusted | -0.2846 | 0.7357 | -1.7265 | 1.1573 | 0.7523 | 0.1779 | 3.1813 | 0.6988 | 0.9567 |
| Ever hyper/manic for 2 days | Regression | binary | Fully adjusted | -0.0767 | 0.2900 | -0.6452 | 0.4918 | 0.9262 | 0.5246 | 1.6352 | 0.7914 | 0.9567 |
| Longest depression period | Regression | continuous | Fully adjusted | -0.0305 | 0.0870 | -0.2010 | 0.1401 | 0.9700 | 0.8179 | 1.1503 | 0.7261 | 0.9567 |
| Neck/shoulder pain 3+ months | Regression | binary | Fully adjusted | -0.1176 | 0.3158 | -0.7365 | 0.5013 | 0.8891 | 0.4788 | 1.6509 | 0.7096 | 0.9567 |
| Falls in last year | Regression | ordered | Fully adjusted | 0.0351 | 0.1136 | -0.1875 | 0.2577 | 1.0357 | 0.8290 | 1.2940 | 0.7572 | 0.9567 |
| Wears glasses/contacts | Regression | binary | Fully adjusted | 0.0580 | 0.2061 | -0.3460 | 0.4620 | 1.0597 | 0.7075 | 1.5872 | 0.7785 | 0.9567 |
| Family history of IBS | Regression | binary | Fully adjusted | 0.0532 | 0.1730 | -0.2859 | 0.3924 | 1.0547 | 0.7513 | 1.4805 | 0.7584 | 0.9567 |
| Strenuous sports duration | Regression | ordered | Fully adjusted | -0.0405 | 0.1379 | -0.3107 | 0.2298 | 0.9604 | 0.7329 | 1.2583 | 0.7692 | 0.9567 |
| Daytime dozing | Regression | ordered | Fully adjusted | 0.0392 | 0.1191 | -0.1942 | 0.2725 | 1.0400 | 0.8235 | 1.3133 | 0.7420 | 0.9567 |
| Reaction time (mean) | Progression | continuous | Fully adjusted | -0.0147 | 0.0551 | -0.1226 | 0.0933 | 0.9855 | 0.8847 | 1.0977 | 0.7901 | 0.9567 |
| Smoking status | Progression | binary | Fully adjusted | 0.1281 | 0.3381 | -0.5346 | 0.7907 | 1.1366 | 0.5859 | 2.2050 | 0.7048 | 0.9567 |
| Alcohol drinker status | Progression | binary | Fully adjusted | 0.0675 | 0.2063 | -0.3369 | 0.4720 | 1.0699 | 0.7140 | 1.6031 | 0.7435 | 0.9567 |
| Guilty feelings | Progression | binary | Fully adjusted | 0.0320 | 0.1105 | -0.1846 | 0.2486 | 1.0325 | 0.8314 | 1.2823 | 0.7721 | 0.9567 |
| General happiness | Progression | ordered | Fully adjusted | -0.0252 | 0.0722 | -0.1667 | 0.1162 | 0.9751 | 0.8465 | 1.1232 | 0.7266 | 0.9567 |
| Difficulty stopping worrying | Progression | binary | Fully adjusted | 0.1354 | 0.5014 | -0.8473 | 1.1181 | 1.1450 | 0.4286 | 3.0592 | 0.7871 | 0.9567 |
| IBS diagnosis | Progression | binary | Fully adjusted | 0.0661 | 0.1702 | -0.2674 | 0.3996 | 1.0684 | 0.7654 | 1.4913 | 0.6976 | 0.9567 |
| Neck/shoulder pain 3+ months | Progression | binary | Fully adjusted | -0.0865 | 0.2769 | -0.6292 | 0.4563 | 0.9172 | 0.5330 | 1.5782 | 0.7549 | 0.9567 |
| Back pain 3+ months | Progression | binary | Fully adjusted | -0.0719 | 0.2311 | -0.5249 | 0.3810 | 0.9306 | 0.5916 | 1.4638 | 0.7556 | 0.9567 |
| Longest depression period | Progression | continuous | Fully adjusted | 0.0267 | 0.0768 | -0.1238 | 0.1771 | 1.0270 | 0.8836 | 1.1938 | 0.7282 | 0.9567 |
| Ever hyper/manic for 2 days | Progression | binary | Fully adjusted | -0.0871 | 0.2553 | -0.5875 | 0.4133 | 0.9166 | 0.5557 | 1.5118 | 0.7330 | 0.9567 |
| Ever highly irritable | Progression | binary | Fully adjusted | 0.0389 | 0.1400 | -0.2355 | 0.3134 | 1.0397 | 0.7902 | 1.3680 | 0.7810 | 0.9567 |
| Glaucoma | Progression | binary | Fully adjusted | -0.1130 | 0.3749 | -0.8478 | 0.6219 | 0.8932 | 0.4283 | 1.8625 | 0.7632 | 0.9567 |
| Painful gums | Progression | binary | Fully adjusted | -0.1308 | 0.3384 | -0.7941 | 0.5325 | 0.8774 | 0.4520 | 1.7032 | 0.6992 | 0.9567 |
| Suffer from nerves | Regression | binary | Partially adjusted | -0.0319 | 0.1280 | -0.2829 | 0.2190 | 0.9686 | 0.7536 | 1.2448 | 0.8031 | 0.9587 |
| Light DIY duration | Progression | ordered | Partially adjusted | 0.0096 | 0.0377 | -0.0643 | 0.0835 | 1.0096 | 0.9377 | 1.0871 | 0.7992 | 0.9587 |
| Trouble falling asleep | Regression | binary | Partially adjusted | -0.0475 | 0.1924 | -0.4246 | 0.3296 | 0.9536 | 0.6540 | 1.3905 | 0.8050 | 0.9587 |
| Chest pain/discomfort | Progression | binary | Partially adjusted | -0.0350 | 0.1426 | -0.3145 | 0.2446 | 0.9657 | 0.7302 | 1.2770 | 0.8064 | 0.9587 |
| Light DIY duration | Regression | ordered | Partially adjusted | -0.0096 | 0.0427 | -0.0934 | 0.0742 | 0.9904 | 0.9108 | 1.0770 | 0.8217 | 0.9587 |
| Sleeplessness/insomnia | Progression | ordered | Partially adjusted | 0.0132 | 0.0572 | -0.0989 | 0.1254 | 1.0133 | 0.9058 | 1.1336 | 0.8170 | 0.9587 |
| Loose teeth | Regression | binary | Fully adjusted | 0.1195 | 0.4917 | -0.8442 | 1.0832 | 1.1269 | 0.4299 | 2.9541 | 0.8080 | 0.9587 |
| Ever disinterested for a week | Regression | binary | Fully adjusted | 0.0272 | 0.1194 | -0.2069 | 0.2613 | 1.0275 | 0.8131 | 1.2986 | 0.8201 | 0.9587 |
| Depression episodes | Regression | ordered | Fully adjusted | 0.0232 | 0.0992 | -0.1713 | 0.2176 | 1.0234 | 0.8425 | 1.2431 | 0.8155 | 0.9587 |
| Daytime dozing | Progression | ordered | Fully adjusted | 0.0241 | 0.1065 | -0.1846 | 0.2327 | 1.0243 | 0.8314 | 1.2621 | 0.8213 | 0.9587 |
| Sleeping too much | Progression | binary | Fully adjusted | -0.0495 | 0.2166 | -0.4741 | 0.3751 | 0.9517 | 0.6225 | 1.4552 | 0.8194 | 0.9587 |
| Unenthusiasm (2 weeks) | Progression | ordered | Fully adjusted | 0.0232 | 0.1012 | -0.1751 | 0.2215 | 1.0235 | 0.8394 | 1.2480 | 0.8186 | 0.9587 |
| Ever disinterested for a whole week | Progression | binary | Fully adjusted | 0.0248 | 0.1050 | -0.1810 | 0.2307 | 1.0251 | 0.8344 | 1.2594 | 0.8133 | 0.9587 |
| Shortness of breath on level ground | Progression | binary | Fully adjusted | 0.0484 | 0.2150 | -0.3731 | 0.4698 | 1.0495 | 0.6886 | 1.5996 | 0.8221 | 0.9587 |
| Chest pain outside activity | Regression | ordered | Partially adjusted | 0.1130 | 0.5329 | -0.9314 | 1.1574 | 1.1197 | 0.3940 | 3.1817 | 0.8320 | 0.9595 |
| Stress: financial | Progression | binary | Partially adjusted | 0.0440 | 0.2171 | -0.3816 | 0.4697 | 1.0450 | 0.6828 | 1.5994 | 0.8393 | 0.9595 |
| Mouth ulcers | Progression | binary | Partially adjusted | 0.0330 | 0.1527 | -0.2662 | 0.3323 | 1.0336 | 0.7663 | 1.3941 | 0.8288 | 0.9595 |
| Longest worried/anxious period | Regression | continuous | Fully adjusted | 0.0303 | 0.1446 | -0.2531 | 0.3137 | 1.0308 | 0.7764 | 1.3685 | 0.8340 | 0.9595 |
| Current smoking | Regression | ordered | Fully adjusted | 0.0444 | 0.2104 | -0.3681 | 0.4569 | 1.0454 | 0.6920 | 1.5791 | 0.8330 | 0.9595 |
| Irritability | Progression | binary | Fully adjusted | -0.0245 | 0.1143 | -0.2484 | 0.1995 | 0.9758 | 0.7800 | 1.2208 | 0.8304 | 0.9595 |
| Fracture from fall | Regression | binary | Fully adjusted | 0.0813 | 0.3925 | -0.6880 | 0.8505 | 1.0847 | 0.5026 | 2.3408 | 0.8359 | 0.9595 |
| Leg pain on walking | Progression | binary | Fully adjusted | -0.0268 | 0.1313 | -0.2842 | 0.2306 | 0.9736 | 0.7526 | 1.2594 | 0.8384 | 0.9595 |
| Wheeze in chest | Progression | binary | Partially adjusted | -0.0235 | 0.1183 | -0.2554 | 0.2085 | 0.9768 | 0.7746 | 1.2318 | 0.8428 | 0.9611 |
| Loose teeth | Regression | binary | Partially adjusted | -0.0698 | 0.4201 | -0.8933 | 0.7537 | 0.9326 | 0.4093 | 2.1248 | 0.8681 | 0.9616 |
| Mouth ulcers | Regression | binary | Partially adjusted | 0.0279 | 0.1708 | -0.3068 | 0.3626 | 1.0283 | 0.7358 | 1.4371 | 0.8702 | 0.9616 |
| Back pain 3+ months | Regression | binary | Partially adjusted | -0.0413 | 0.2139 | -0.4607 | 0.3780 | 0.9595 | 0.6309 | 1.4593 | 0.8467 | 0.9616 |
| Wears glasses/contacts | Regression | binary | Partially adjusted | 0.0306 | 0.1822 | -0.3264 | 0.3877 | 1.0311 | 0.7215 | 1.4736 | 0.8665 | 0.9616 |
| Irritability | Progression | binary | Partially adjusted | 0.0166 | 0.0975 | -0.1745 | 0.2077 | 1.0167 | 0.8398 | 1.2308 | 0.8651 | 0.9616 |
| Worry too long after embarrassment | Progression | binary | Partially adjusted | 0.0157 | 0.0855 | -0.1519 | 0.1832 | 1.0158 | 0.8591 | 1.2011 | 0.8545 | 0.9616 |
| Depression episodes | Progression | ordered | Partially adjusted | 0.0134 | 0.0748 | -0.1332 | 0.1601 | 1.0135 | 0.8753 | 1.1736 | 0.8573 | 0.9616 |
| Loneliness/isolation | Regression | binary | Fully adjusted | 0.0302 | 0.1587 | -0.2808 | 0.3413 | 1.0307 | 0.7552 | 1.4067 | 0.8488 | 0.9616 |
| Snoring | Progression | binary | Fully adjusted | -0.0199 | 0.1079 | -0.2314 | 0.1916 | 0.9803 | 0.7935 | 1.2112 | 0.8537 | 0.9616 |
| Diabetes-related eye disease | Regression | binary | Fully adjusted | 0.2102 | 1.2293 | -2.1992 | 2.6195 | 1.2339 | 0.1109 | 13.7293 | 0.8642 | 0.9616 |
| Tenseness/restlessness (2 weeks) | Regression | ordered | Fully adjusted | 0.0179 | 0.1041 | -0.1860 | 0.2219 | 1.0181 | 0.8302 | 1.2485 | 0.8631 | 0.9616 |
| Other eye problems | Progression | binary | Fully adjusted | -0.0229 | 0.1352 | -0.2880 | 0.2422 | 0.9774 | 0.7498 | 1.2740 | 0.8656 | 0.9616 |
| Facial pain 3+ months: No | Progression | binary | Fully adjusted | 0.1892 | 1.1550 | -2.0745 | 2.4530 | 1.2083 | 0.1256 | 11.6226 | 0.8699 | 0.9616 |
| Stress: divorce/separation | Regression | binary | Partially adjusted | -0.0691 | 0.4324 | -0.9167 | 0.7784 | 0.9332 | 0.3998 | 2.1781 | 0.8730 | 0.9624 |
| Duration of vigorous activity | Regression | continuous | Partially adjusted | -0.0088 | 0.0564 | -0.1194 | 0.1019 | 0.9913 | 0.8875 | 1.1072 | 0.8765 | 0.9640 |
| Sleeplessness/insomnia | Progression | ordered | Fully adjusted | -0.0098 | 0.0676 | -0.1423 | 0.1226 | 0.9902 | 0.8674 | 1.1305 | 0.8844 | 0.9704 |
| Smoking status | Progression | binary | Partially adjusted | -0.0371 | 0.2818 | -0.5895 | 0.5152 | 0.9635 | 0.5546 | 1.6740 | 0.8951 | 0.9729 |
| General happiness | Regression | ordered | Fully adjusted | -0.0109 | 0.0814 | -0.1703 | 0.1486 | 0.9892 | 0.8434 | 1.1602 | 0.8939 | 0.9729 |
| Ever worried more than others | Regression | binary | Fully adjusted | 0.0218 | 0.1590 | -0.2897 | 0.3334 | 1.0221 | 0.7485 | 1.3957 | 0.8907 | 0.9729 |
| Tenseness/restlessness (2 weeks) | Progression | ordered | Fully adjusted | -0.0123 | 0.0924 | -0.1933 | 0.1688 | 0.9878 | 0.8242 | 1.1838 | 0.8943 | 0.9729 |
| Difficulty stopping worrying | Progression | binary | Partially adjusted | 0.0526 | 0.4303 | -0.7908 | 0.8959 | 1.0540 | 0.4535 | 2.4497 | 0.9027 | 0.9788 |
| Ever hyper/manic for 2 days | Progression | binary | Partially adjusted | 0.0275 | 0.2314 | -0.4261 | 0.4812 | 1.0279 | 0.6531 | 1.6179 | 0.9053 | 0.9793 |
| Mood swings | Progression | binary | Fully adjusted | 0.0114 | 0.1038 | -0.1920 | 0.2148 | 1.0115 | 0.8253 | 1.2396 | 0.9125 | 0.9848 |
| Tiredness/lethargy (2 weeks) | Regression | ordered | Partially adjusted | -0.0064 | 0.0599 | -0.1239 | 0.1111 | 0.9936 | 0.8835 | 1.1175 | 0.9152 | 0.9854 |
| Ever highly irritable | Regression | binary | Partially adjusted | 0.0106 | 0.1396 | -0.2631 | 0.2843 | 1.0106 | 0.7686 | 1.3288 | 0.9397 | 0.9878 |
| Longest depression period | Regression | continuous | Partially adjusted | -0.0059 | 0.0727 | -0.1484 | 0.1366 | 0.9941 | 0.8621 | 1.1463 | 0.9354 | 0.9878 |
| Restless (anxiety) | Progression | binary | Partially adjusted | -0.0148 | 0.1866 | -0.3804 | 0.3509 | 0.9853 | 0.6836 | 1.4203 | 0.9368 | 0.9878 |
| Leg pain on walking | Progression | binary | Partially adjusted | 0.0102 | 0.1112 | -0.2077 | 0.2281 | 1.0102 | 0.8125 | 1.2562 | 0.9270 | 0.9878 |
| Fluid intelligence | Regression | ordered | Fully adjusted | -0.0024 | 0.0297 | -0.0606 | 0.0558 | 0.9976 | 0.9412 | 1.0574 | 0.9350 | 0.9878 |
| Snoring | Regression | binary | Fully adjusted | -0.0122 | 0.1222 | -0.2517 | 0.2273 | 0.9879 | 0.7775 | 1.2552 | 0.9207 | 0.9878 |
| Ever worried more than others | Progression | binary | Fully adjusted | 0.0114 | 0.1403 | -0.2636 | 0.2863 | 1.0114 | 0.7683 | 1.3316 | 0.9355 | 0.9878 |
| Waking too early | Progression | binary | Fully adjusted | 0.0157 | 0.2111 | -0.3980 | 0.4293 | 1.0158 | 0.6717 | 1.5362 | 0.9409 | 0.9878 |
| Tiredness/lethargy (2 weeks) | Regression | ordered | Fully adjusted | -0.0065 | 0.0710 | -0.1456 | 0.1326 | 0.9935 | 0.8645 | 1.1418 | 0.9270 | 0.9878 |
| Falls in last year | Progression | ordered | Fully adjusted | -0.0082 | 0.1007 | -0.2055 | 0.1891 | 0.9918 | 0.8142 | 1.2081 | 0.9348 | 0.9878 |
| Shortness of breath on level ground | Regression | binary | Fully adjusted | 0.0184 | 0.2452 | -0.4623 | 0.4991 | 1.0185 | 0.6298 | 1.6472 | 0.9403 | 0.9878 |
| Painful gums | Progression | binary | Partially adjusted | -0.0208 | 0.2996 | -0.6081 | 0.5666 | 0.9795 | 0.5444 | 1.7622 | 0.9448 | 0.9897 |
| Vigorous activity days/weeks | Regression | ordered | Partially adjusted | -0.0012 | 0.0246 | -0.0495 | 0.0471 | 0.9988 | 0.9517 | 1.0482 | 0.9614 | 0.9929 |
| Any vascular/heart problem | Regression | binary | Partially adjusted | -0.0248 | 0.6834 | -1.3641 | 1.3146 | 0.9755 | 0.2556 | 3.7233 | 0.9711 | 0.9929 |
| Leg pain on walking | Regression | binary | Partially adjusted | -0.0027 | 0.1245 | -0.2467 | 0.2414 | 0.9973 | 0.7813 | 1.2730 | 0.9829 | 0.9929 |
| Ever hyper/manic for 2 days | Regression | binary | Partially adjusted | -0.0076 | 0.2608 | -0.5187 | 0.5036 | 0.9924 | 0.5953 | 1.6546 | 0.9768 | 0.9929 |
| Longest worried/anxious period | Regression | continuous | Partially adjusted | 0.0060 | 0.1228 | -0.2348 | 0.2468 | 1.0060 | 0.7908 | 1.2799 | 0.9609 | 0.9929 |
| Alcohol drinker status | Regression | binary | Partially adjusted | -0.0079 | 0.2015 | -0.4029 | 0.3872 | 0.9922 | 0.6684 | 1.4728 | 0.9689 | 0.9929 |
| Alcohol drinker status | Progression | binary | Partially adjusted | 0.0047 | 0.1805 | -0.3491 | 0.3584 | 1.0047 | 0.7053 | 1.4310 | 0.9794 | 0.9929 |
| Long-standing illness/disability | Progression | binary | Partially adjusted | -0.0054 | 0.1008 | -0.2030 | 0.1923 | 0.9946 | 0.8163 | 1.2120 | 0.9576 | 0.9929 |
| Any vascular/heart problem | Regression | binary | Fully adjusted | 0.0099 | 0.6899 | -1.3423 | 1.3621 | 1.0099 | 0.2612 | 3.9044 | 0.9886 | 0.9929 |
| Back pain 3+ months | Regression | binary | Fully adjusted | 0.0128 | 0.2607 | -0.4982 | 0.5239 | 1.0129 | 0.6076 | 1.6886 | 0.9608 | 0.9929 |
| Major dietary changes (5 years) | Regression | unordered | Fully adjusted | 0.0015 | 0.0606 | -0.1172 | 0.1203 | 1.0015 | 0.8894 | 1.1278 | 0.9800 | 0.9929 |
| Alcohol intake frequency | Regression | ordered | Fully adjusted | -0.0013 | 0.0416 | -0.0828 | 0.0801 | 0.9987 | 0.9205 | 1.0834 | 0.9742 | 0.9929 |
| Major dietary changes (5 years) | Progression | unordered | Fully adjusted | -0.0020 | 0.0535 | -0.1069 | 0.1029 | 0.9980 | 0.8986 | 1.1084 | 0.9698 | 0.9929 |
| Worry too long after embarrassment | Progression | binary | Fully adjusted | -0.0016 | 0.1007 | -0.1990 | 0.1957 | 0.9984 | 0.8195 | 1.2162 | 0.9869 | 0.9929 |
| Restless (anxiety) | Progression | binary | Fully adjusted | -0.0082 | 0.2206 | -0.4407 | 0.4242 | 0.9918 | 0.6436 | 1.5284 | 0.9702 | 0.9929 |
| Long-standing illness/disability | Progression | binary | Fully adjusted | -0.0067 | 0.1186 | -0.2392 | 0.2257 | 0.9933 | 0.7873 | 1.2532 | 0.9547 | 0.9929 |
| Stress: divorce/separation | Progression | binary | Fully adjusted | -0.0102 | 0.4117 | -0.8171 | 0.7967 | 0.9898 | 0.4417 | 2.2182 | 0.9802 | 0.9929 |
| Mouth ulcers | Progression | binary | Fully adjusted | 0.0097 | 0.1810 | -0.3451 | 0.3645 | 1.0098 | 0.7082 | 1.4399 | 0.9571 | 0.9929 |
| Puzzle score | Progression | ordered | Fully adjusted | -0.0004 | 0.0273 | -0.0539 | 0.0531 | 0.9996 | 0.9475 | 1.0546 | 0.9886 | 0.9929 |
| Neuroticism | Progression | ordered | Partially adjusted | 0.0000 | 0.0144 | -0.0281 | 0.0282 | 1.0000 | 0.9723 | 1.0286 | 0.9982 | 0.9990 |
| Macular degeneration | Progression | binary | Partially adjusted | -0.0005 | 0.3770 | -0.7394 | 0.7385 | 0.9995 | 0.4774 | 2.0928 | 0.9990 | 0.9990 |

*Abbreviations:* BMI, body mass index; BP, blood pressure; CI, confidence interval; FDR, false-discovery rate corrected; OR, odds ratio; SE, standard error; UKB, UK Biobank. *Notes:* Comparison is versus Stable as the reference group.
